# Pain across the lifespan: global and regional reference curves from 6.1 million individuals in 118 countries

**DOI:** 10.64898/2026.04.21.26351327

**Authors:** Matt Fillingim, Christophe Tanguay-Sabourin, Lindsay Neuert, Azin Zare, Jax Norman, Gianluca V. Guglietti, Lise Hobeika, Abimbola Ayorinde, Ahmad Salimzadeh, Ahmad-reza Jamshidi, Arash Tehrani-Banihashemi, Beatriz Olaya, Benjamin Longo Mbenza, Bibilola Oladeji, Brenda Penninx, Buddhi Paudyal, Caique de Melo, Chantal Berna-Renella, Clarissa Humberg, Claude-Pierre Pascal, Elisa Smith, Elizabeth VanDenKerkhof, Erik J. Giltay, Esther Garcia-Esquinas, Faisal Parlindungan, Fatima Qayyum Abbasi, Fernando Rodríguez Artalejo, Gareth T Jones, Gary Slade, Hanifa Bouziri, Herta Flor, Hiroyasu Iso, Ian Gilron, Imad Uthman, Ingris Peláez-Ballestas, Jean-Paul Devengi Nzambi, Jose Luis Ayuso-Mateos, Josep Maria Haro, Julia Wager, Juliana Barcellos de Souza, Kazutoshi Nakamura, Keiko Yamada, Lisa Marie Rau, Makram Talih, Marcus J Beasley, María Dueñas, Marie Zins, Marta Miret, Melissa Wake, Mirza Zaka Pratama, Mohammad Almalki, Monique Chaaya, Muhammad Anshory, Naoki Kondo, Navid Moghadam, Norma Mansor, Omar Alsaed, Orawan Keeratisiroj, Oye Gureje, Pande Ketut Kurniari, Pardis Noormohammadpour, Pierrot Lebughe, Raga A Elzahaf, Ramin Kordi, Raquel Lucas, Rashmi Parekh Bhandari, Rudy Hidayat, Saba Samreen, Samar Al-Emadi, Shima RoknSharifi, Shinsuke Inoue, Sudeep Adhikari, Suryo Anggoro Kusumo Wibowo, Sweekriti Sharma, Sylvia Villeneuve, Takahiro Tabuchi, Tayyeba Khursheed, Tie Yamato, Todd Jackson, Veronica Souza Santos, Wantana Siritaratiwat, Xianchen Liu, Zhen-Zhen Liu, Tobias Banaschewski, Gareth J. Barker, Arun L.W. Bokde, Rudiger Bruhl, Sylvane Desrivieres, Penny Gowland, Antoine Grigis, Andreas Heinz, Herve Lemaitre, Jean-Luc Martinot, Marie-Laure Paillere Martinot, Eric Artiges, Frauke Nees, Dimitri Papadopoulos Orfanos, Tomas Paus, Luise Poustka, Michael N. Smolka, Sarah Hohmann, Nilakshi Vaidya, Henrik Walter, Robert Whelan, Paul Wirsching, Gunter Schumann, Susan M Sawyer, Saman Khalatbari-Soltani, Olof Anna Steingrimsdottir, Gary J Macfarlane, Fiona Blyth, Etienne Vachon-Presseau

**Author notes:** Corresponding author: Matt Fillingim; Etienne Vachon-Presseau. **Ethics approval** The pooled analysis was approved by the McGill University Faculty of Medicine and Health Sciences Research Ethics Board (IRB Study Number A12-M69-23B; 23-10-068). The study involved secondary analysis of de-identified individual-level data obtained from public repositories and through data transfer or data use agreements with collaborating institutions. **Ethics and inclusion statement** This study synthesised individual-level data from population-based surveys, cohort studies, and national health studies conducted across multiple world regions. Data were obtained through publicly available repositories or through data transfer and data use agreements with collaborating researchers and institutions. The collaboration included investigators from diverse geographic regions who contributed expertise in epidemiology, clinical pain research, population health, and lived-experiences of pain. **Contributors** MF and EVP conceived the study. MF conducted data harmonisation, statistical analyses, and figure generation. MF and EVP drafted the manuscript. CTS, AZ, JN, GVG, LN, and LH contributed to manuscript refinement. SMS, SKS, OAS, GM, and FMB provided senior input on manuscript development. All other authors contributed data or cohort expertise and reviewed the final manuscript. EVP supervised the study. **Data sharing** Detailed and annotated analysis code is publicly available via GitHub (https://github.com/EVPlab). An interactive benchmarking tool and downloadable summary statistics from this study are publicly available at https://evppainlab.shinyapps.io/global-pain-benchmark/. Researchers seeking access to the original individual-level data should apply directly to the respective data custodians according to their access policies.

## Abstract

Pain is the leading cause of disability worldwide, yet no harmonised self-reported reference framework exists to characterise how its burden is distributed across the lifespan and world regions. Here, we harmonised individual-level self-reported pain data from 6,075,021 participants across 894 population-based data sources in 118 countries to establish global reference trajectories of pain. We implemented these trajectories in an open-access benchmarking platform for positioning external datasets against global pain norms. Pain prevalence ranged from 2.5% for facial pain to 45.0% for back pain, was consistently higher in women across all eleven anatomical sites (risk ratio range 1.09 to 1.83), and increased most steeply before age 55 years. Contrary to existing estimates that generally project higher prevalence of pain conditions in higher Human Development Index (HDI) regions, we found that individuals in the lowest HDI countries experienced nearly twice the late-life prevalence of any bodily pain compared with those in the highest (risk difference 31.8 percentage points [95% CI 30.1–33.6]). Globally, 18.3% of pain burden across anatomical sites was attributable to three modifiable risk factors (smoking, obesity, and low income) but this varied from 12.6% in sub-Saharan Africa to 27.1% in eastern Europe, indicating that the drivers of pain in lower-HDI settings remain poorly characterised.

## Main

Pain is a leading cause of years lived with disability worldwide^1^. Unlike most major health conditions, it lacks an objective biomarker and cannot be assessed independently of self-report^2^. As a result, its population burden depends on how pain is captured across studies and health systems. Existing frameworks, including the Global Burden of Disease (GBD) study, rely on clinically defined conditions and modelled estimates of heterogeneous data sources that may not fully capture pain that does not enter diagnostic pathways, particularly in settings with limited access to care.

Although self-reported pain is routinely collected in population-based studies, these data are fragmented across heterogeneous instruments, recall periods, and study designs that have not been systematically harmonised at global scale. Reported prevalence in the general population therefore varies widely, ranging from 8% to 64% across studies^3–5^, with much of the evidence being derived from high-income settings, often focusing on single phenotypes such as low back pain. Dimensions more closely related to disability and care seeking, such as pain intensity and widespread pain, are also rarely captured in comparable ways across populations ^6,7^. Consequently, the absence of a consistent framework leaves fundamental questions unresolved: when does pain emerge across the lifespan and how do these trajectories differ by phenotypes, sex, and world region. Establishing reference trajectories for pain would provide a common ground for comparing cohorts, clinical populations, and countries, and for assessing whether patterns derived from self-report align with those from condition-based surveillance, particularly in settings underrepresented in global estimates.

In this study, we applied generalized linear mixed-effects model on individual-level self-reported pain data from a global network of population-based studies to construct reference trajectories of pain across the lifespan, implemented in an open-access benchmarking platform. We present distinct lifespan trajectories that differ by anatomical site and population context, with most burden accumulating most rapidly before age 55 years. Moreover, we report that self-reported pain is substantially higher in lower-human development settings than condition-based estimates suggest, with the drivers of this excess poorly understood and distinct from the modifiable risk factors that dominate pain burden in high-income settings.

## Results

### Study population and data coverage

We compiled individual-participant data from population-based national, regional, and international health studies and surveys conducted between 1990 and 2025 that assessed self-reported bodily pain (Supplementary Table 1). We analysed a total of 6,075,021 participants across 894 data sources from 134 population-based study programmes, spanning 118 countries and territories (figure 1A). Participants were aged 5 to 100+ years, and 55% were female. Studies represented all inhabited continents, and data density ranged from 1.8 data sources per country in sub-Saharan Africa to 19.8 in western Europe (figure 1B). The age distribution of contributing study populations is shown in figure 1C.

**Figure 1.**
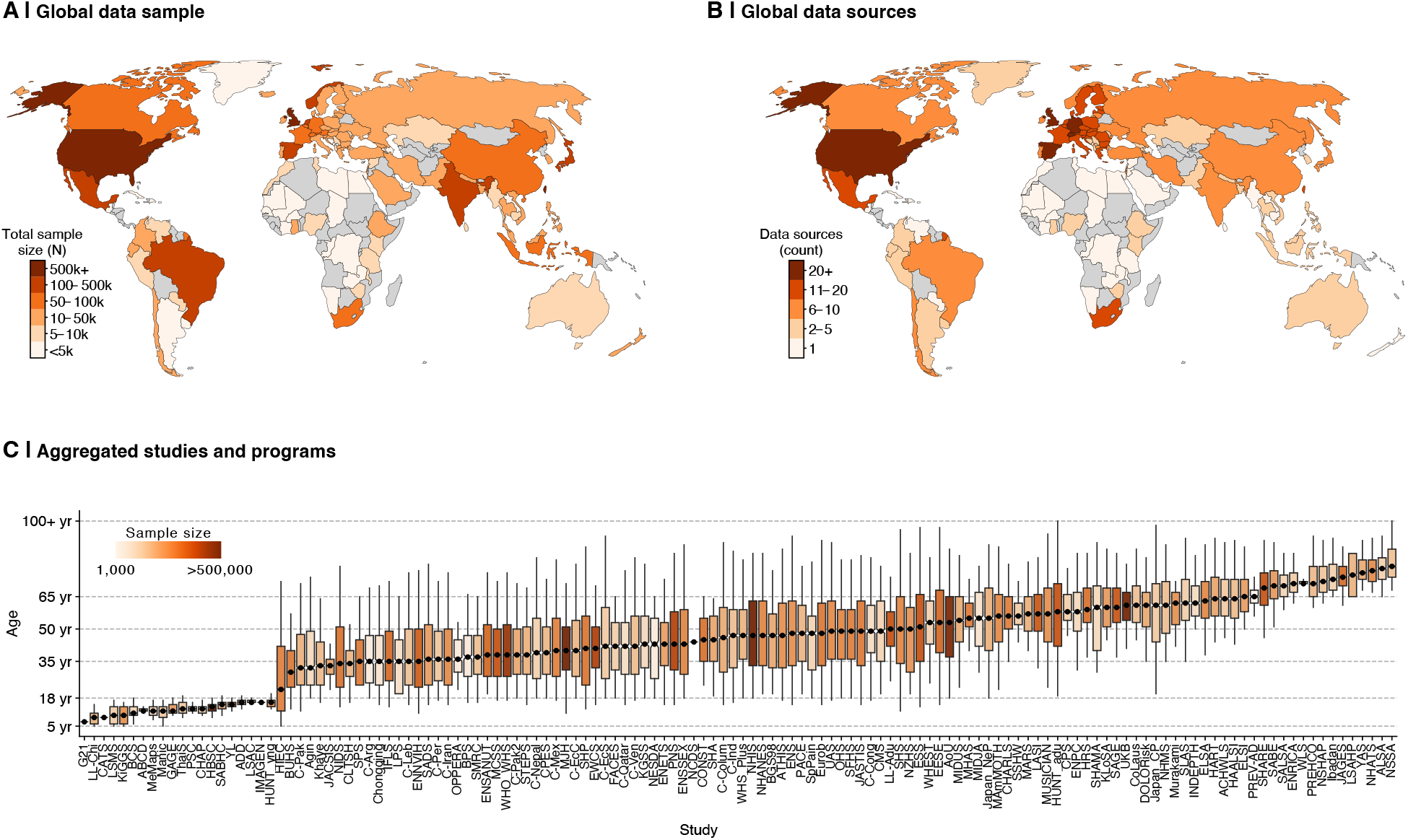
Global coverage and age structure of the pooled dataset. (A) Country-level total sample size (participants with valid data for at least one pain outcome), shown in categories from <5,000 to ≥500,000. (B) Number of contributing data sources per country (distinct surveys and cohorts), shown in categories from 1 to ≥20. Countries without eligible data are shown in grey. (C) Age distribution for each contributing study program or data source, plotted as boxplots (interquartile range with whiskers indicating the age range) and ordered by median age (black points). Box colour indicates study sample size (1,000 to >500,000 participants).

### Lifespan pain trajectories

Pain prevalence estimates are known to vary with methodological factors. To generate global reference curves, our harmonisation procedure mapped study-specific wording and body-map coding schemes to standardised categories (Supplementary Section 3) after evaluating seven key methodological features (see Methods and Supplementary Section 6.2) through a series of sensitivity analyses (Supplementary Section 6, Supplementary Figs. 1–12). Binary indicators of self-reported pain were harmonised at 11 major anatomical sites: head, face, neck or shoulder, foot or ankle, hand or wrist, elbow, chest, back, stomach or abdomen, hip, and knee. Age-specific trajectories of pain prevalence were estimated across the lifespan (ages 5 to 100+; Supplementary Section 6.5) using a generalized linear mixed-effects model (GLMM) with a logit link, incorporating natural cubic splines to flexibly capture nonlinear age trajectories. Random effects accounted for the hierarchical correlation structure of the data, with individuals nested within cohorts and world regions.

Drawing on individual-level data from 1.2 to 4.9 million distinct participants per pain site, pain prevalence varied markedly across anatomical sites and followed distinct age trajectories by site and sex (figure 2). Overall prevalence ranged from 2.5% for facial pain to 45.0% for back pain. Women consistently reported higher prevalence than men across all 11 sites (RR 1.09 to 1.83), with the largest sex differences observed for facial (RR 1.83 [95% CI 1.80–1.87]), headache (RR 1.78 [1.76–1.79]), and stomach/abdominal pain (RR 1.62 [1.60–1.64]). Musculoskeletal sites including back, hip, and knee showed progressive non-linear increases through adulthood, with the most rapid gains between ages 20 and 55 years and peaking at or beyond age 75. Strikingly, the majority of sites (7 of 11) exhibited an inverted U trajectory, with prevalence peaking in mid-to-late adulthood and declining thereafter. This pattern was particularly evident in women for headache (peak age 34 years) and abdominal pain (22 years), and across upper-body sites including neck or shoulder (58 to 62 years), elbow (58 to 59 years), and facial pain (42 to 45 years). Our results refine the prevailing assumption that pain accumulates monotonically with age and instead suggests that several common pain phenotypes peak by mid-to-late adulthood and decline thereafter. Across all sites, prevalence rose most steeply before age 55, indicating that the majority of pain burden accumulates before midlife. These trajectories are publicly accessible through the Global Pain Benchmarking Tool (https://evppainlab.shinyapps.io/global-pain-benchmark/; Extended Data Fig. 7; Supplementary Section 10).

**Figure 2.**
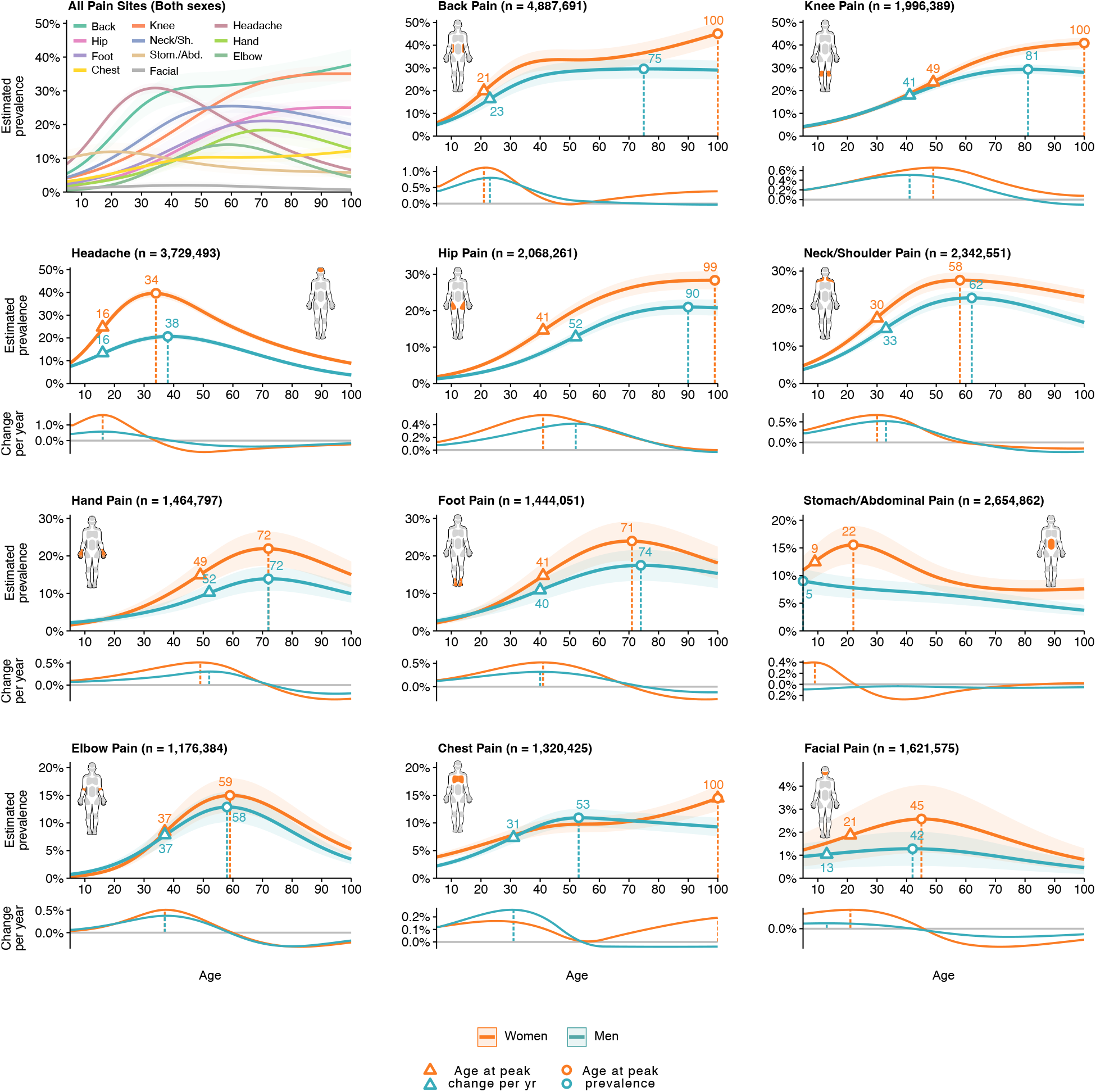
Lifespan trajectories of pain prevalence across for eleven anatomical sites. Each site-specific trajectory was estimated using a mixed-effects logistic regression model with natural cubic splines (3 degrees of freedom, selected via study-clustered 5-fold cross-validation) to capture nonlinear age effects, fitted to individual-level data from 894 population-based sources nested within cohorts and world regions. Trajectories are marginal predictions standardised to a past-month recall period and general case definition. For each site, the upper panel shows model-estimated prevalence (%) across the lifespan (ages 5 to 100+), and the lower panel shows the corresponding age-specific rate of change (percentage-point change per year). Shaded bands indicate 95% confidence intervals. Open triangles mark the age of peak rate of increase in prevalence; open circles mark the age of peak prevalence. Colours denote sex (women: orange; men: blue). Annotated ages are shown in the prevalence panels, with corresponding vertical dashed lines in the rate-of-change panels. Sample sizes reflect the number of distinct individual-level records per site and are shown in panel titles.

### Any bodily pain, high-intensity pain, and generalised pain across the lifespan

A composite any bodily pain outcome was coded as present if pain was reported at any site (Supplementary Section 3). Secondary outcomes included high-intensity pain (average intensity of 7 or higher on a 0–10 scale) and generalised pain (pain in at least four of five body regions following the 2016 fibromyalgia spatial criteria^8^), capturing dimensions of burden more closely related to disability and care seeking^6,7,9^. Here, the prevalence of any bodily pain increased progressively across the lifespan, from approximately 30% at age 5 years to 70% at age 100 years or older (figure 3). All three aggregate pain measures were consistently higher in women than in men: any bodily pain (RR 1.16 [CI 1.15–1.17]), high-intensity pain (RR 1.64 [1.60–1.66]), and generalised pain (RR 1.49 [1.46–1.53]). Whereas any bodily pain increased with age, the severity-based measures showed mid-to late-life peaks followed by declines, with high-intensity pain peaking around 50 years and generalised pain peaking later, around age 70 years. Average pain intensity among individuals reporting pain followed a similar non-linear trajectory, peaking at approximately 5.0 in women and 4.6 in men around age 48 before declining through later life (Extended Data Fig. 6). Overall, our results suggest that while the likelihood of experiencing any pain rises steadily with age, the most disabling forms of pain followed an inverted-U trajectory, peaking in mid-to-late adulthood before declining. The distinction, between pain accumulation and pain that intensifies then resolves, has direct implications for where prevention and care should be concentrated across the lifespan.

**Figure 3.**
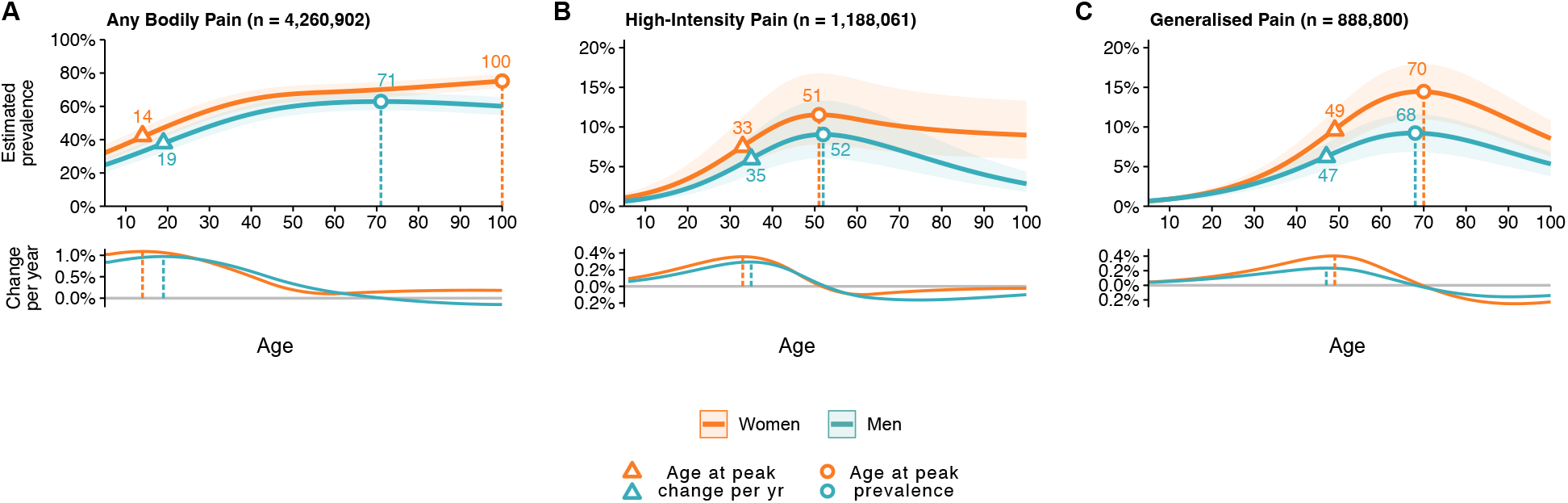
Lifespan trajectories of any bodily pain, high-intensity pain, and generalised pain. Each trajectory was estimated using a mixed-effects logistic regression model with natural cubic splines (3 degrees of freedom, selected via study-clustered 5-fold cross-validation), fitted to individual-level data from 894 population-based sources nested within cohorts, countries, and world regions. Trajectories are marginal predictions standardised to a past-month recall period and general case definition. (A) Any bodily pain prevalence. (B) High-intensity pain, defined as NRS ≥7. (C) Generalised pain, defined as pain in ≥4 of 5 body regions per the 2016 ACR fibromyalgia spatial criteria. In each panel, the upper plot shows model-estimated prevalence (%) across the lifespan (ages 5 to 100+), and the lower plot shows the corresponding age-specific rate of change (percentage-point change per year). Shaded bands indicate 95% confidence intervals. Open triangles mark the age of peak rate of increase in prevalence; open circles mark the age of peak prevalence. Colours denote sex (women: orange; men: blue). Vertical dashed lines correspond to the marked ages. Sample sizes reflect the number of distinct individual-level records and are shown in panel titles.

### Regional variation and national development

Regional trajectories revealed a consistent pattern of early-life convergence and late-life divergence. For any bodily pain and joint pain, prevalence was broadly similar across regions in early adulthood but diverged substantially after age 65 years (figure 4A). Eastern Europe showed the greatest late-life excess, reaching approximately 21 percentage points above the global reference at age 80 or older, while western Europe remained below it throughout late adulthood. Sub-Saharan Africa, central and southern Asia, and north Africa and west Asia showed comparatively low prevalence in early adulthood followed by steeper increases after midlife, ultimately converging with or exceeding the global reference in older age. Back pain followed a similar pattern, diverging across regions after approximately age 50 years, with continued increases into late life in eastern Europe, sub-Saharan Africa, and central and southern Asia, and plateauing or declining trajectories in western Europe and northern America.

**Figure 4.**
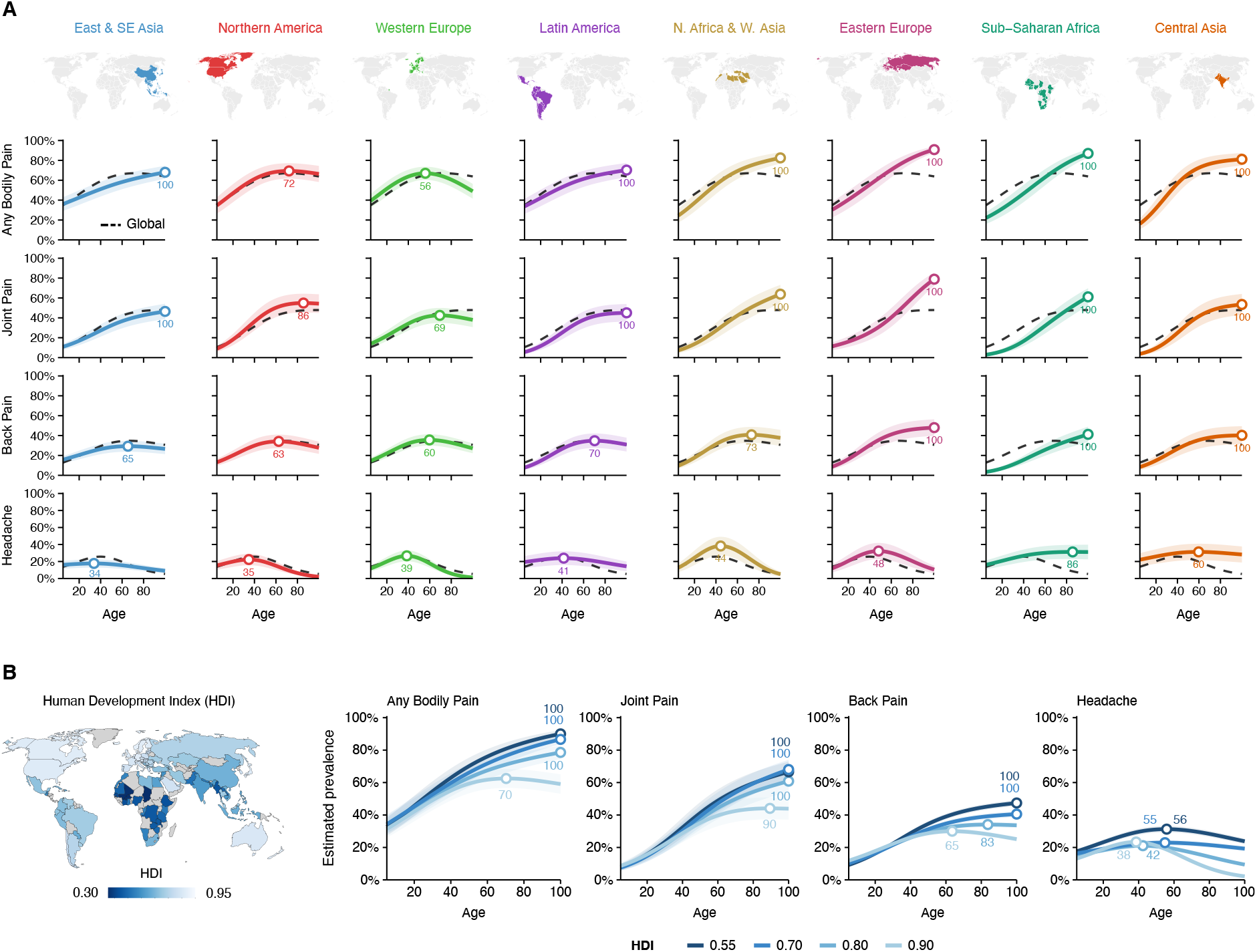
Regional variation and national development in lifespan pain trajectories. (A) Model-estimated prevalence (%) by age for any bodily pain, joint pain, back pain, and headache across eight world regions (Oceania excluded due to insufficient data), defined by the United Nations Sustainable Development Goals (SDG) regional classification. Each column corresponds to one world region, with contributing countries highlighted on the inset world map above the column, and each row to a pain phenotype. Solid coloured lines show region-specific trajectories, open circles mark the age of peak prevalence (annotated with the corresponding age), and shaded bands denote 95% confidence intervals. The dashed black line shows the global reference trajectory estimated from the full pooled dataset. (B) Model-estimated prevalence trajectories for the same four pain outcomes as a continuous function of country-level Human Development Index (HDI), plotted at four representative values (0.55, 0.70, 0.80, 0.90) spanning low to very high development. The inset map shows country-level HDI values used in the analysis; countries without eligible data are shown in grey. Open circles mark the age of peak prevalence; shaded bands denote 95% confidence intervals.

Headache showed the greatest regional heterogeneity, differing not only in magnitude but in trajectory shape. Northern America and western Europe followed an early-adulthood peak and decline consistent with the global reference, whereas east and southeast Asia showed consistently lower and flatter prevalence across the lifespan. Eastern Europe and north Africa and west Asia showed later peaks with more gradual declines, and sub-Saharan Africa showed a modest monotonic increase from adolescence into later life. These contrasting patterns highlight how regional trajectories can differ substantially from the global reference when data from historically underrepresented regions are examined directly.

To examine whether regional differences partly reflected variation in national development, we estimated trajectories as a continuous function of country-level Human Development Index (HDI; figure 4B). From age 40 years onward, the prevalence of any bodily pain, joint pain, and back pain increased more steeply in countries with low HDI values, such that by age 80 years or older, people in low-HDI settings experienced nearly twice the prevalence observed in high-HDI settings (any bodily pain: risk difference 31.8 percentage points [95% CI 30.1–33.6]; joint pain: 26.6 percentage points [24.4–28.9]; back pain: 20.8 percentage points [19.2–22.3]). Headache prevalence was higher in lower-HDI settings throughout the lifespan. In high-HDI settings, headache prevalence declined after age 35 years, whereas this decline was attenuated in lower-HDI settings.

### Individual risk factors and socioeconomic gradients

Smoking, obesity, and low household income each showed distinct age-specific and anatomically patterned associations with pain prevalence (figure 5A; Extended Data Fig. 1). Current smoking was associated with a modest but consistent excess across pain sites from adolescence to age 65 years, with the largest effect observed for facial pain (RR 1.66 [1.62–1.69] versus never smokers; Extended Data Fig. 2). Obesity showed a progressively diverging association from early adulthood onward, concentrated at weight-bearing joints: foot or ankle (RR 1.89 [1.81–1.96] versus normal weight), knee (1.85 [1.81–1.89]), and hip (1.61 [1.53–1.67]), peaking in midlife (Extended Data Fig. 3). Household income level showed a pronounced dose–response pattern across all pain sites from early adulthood onward (Extended Data Fig. 4). This gradient was steepest for chest pain, with individuals in the lowest income level experiencing more than twice the risk compared with those in the highest (RR 2.18 [2.11–2.26]), a disparity that peaked in midlife before converging in older age.

**Figure 5.**
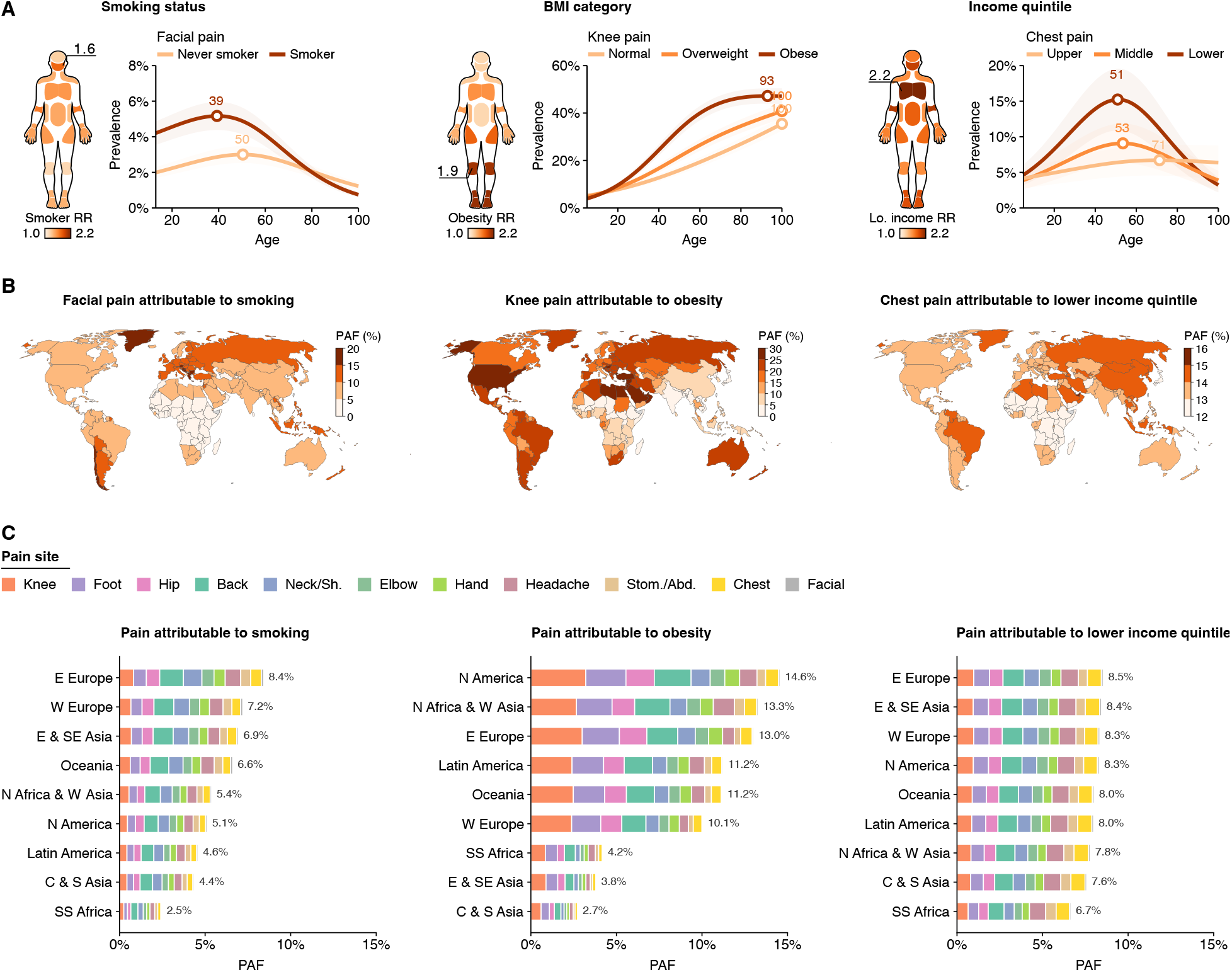
Risk factors, site-specific associations, and population attributable fractions for pain across the lifespan. (A) Body maps display lifespan risk ratios (RR) for each anatomical site: current versus never smokers (left), people with obesity versus normal weight (centre), and lowest versus highest household income quintile (right); colour intensity indicates RR magnitude. Accompanying line plots show estimated prevalence (%) across age with 95% confidence intervals for a representative site per exposure: facial pain for smoking, knee pain for people with obesity (with overweight as an intermediate category), and chest pain for income (with the middle quintile for reference). Open circles and annotated ages mark the age of peak prevalence. (B) Country-level population attributable fractions (PAFs) for the same sentinel site–exposure pairs. PAF colour scales differ across exposures to reflect variation in range. (C) Total PAF across all pain sites by world region for each exposure. Stacked bars show the contribution of each anatomical site to the regional total.

To quantify how much of the population-level pain burden these exposures collectively explain, we estimated population attributable fractions across regions (figure 5B,C). Obesity showed the widest regional variation, ranging from 2.7% in central and southern Asia to 14.6% in northern America, with high burden also concentrated in north Africa and west Asia and eastern Europe. Smoking-attributable fractions ranged from 2.5% in sub-Saharan Africa to 8.4% in eastern Europe, and income-attributable fractions were more consistent across regions (ranging between 6.7%-8.5%). Globally, the combined attributable burden reached 27.1% in eastern Europe compared with 12.6% in sub-Saharan Africa (Extended Data Fig. 5, Supplementary Table 6). In sub-Saharan Africa and central and southern Asia, obesity and smoking PAFs were each below 5%, indicating that the exposures responsible for the largest share of pain burden in high HDI regions account for little of the burden in the lower HDI regions, where the drivers of that burden remain poorly understood.

## Discussion

This study integrates individual-level self-reported pain data from over six million participants, spanning ages 5 to over 100 years, representing one of the largest harmonisation efforts in pain epidemiology to date. Using raw participant-level observations rather than aggregated statistics or clinically defined conditions, we constructed global reference trajectories of pain across the lifespan, including age groups that remain sparsely represented in current surveillance. These curves provide a common framework against which individual cohorts, clinical populations, and countries can be compared. Drawing on 894 population-based sources across 118 countries, these curves provide sufficient geographic coverage to directly test whether self-reported pain burden aligns with or diverges from condition-based global estimates.

The global lifespan trajectories show that pain is not a single epidemiological phenomenon but a set of distinct nonlinear patterns that differ in their timing and anatomical distribution. Across most phenotypes, pain did not increase monotonically with age; instead, the steepest rises occurred before age 55 years, indicating that a substantial share of lifetime pain burden develops during working-age adulthood. Trajectories varied markedly by phenotype: headache, abdominal, and facial pain peaked earlier in life before declining, whereas musculoskeletal pain generally rose progressively with age. Within musculoskeletal sites, upper body pain (including neck, shoulder, and elbow) peaked around midlife and declined thereafter, while lower body pain (including back, hip, and knee) continued to increase across the lifespan, consistent with the greater role of cumulative mechanical and degenerative load at weight-bearing sites. A similar pattern was observed for severity-based measures, with both high-intensity and generalised pain peaking in midlife before declining. Overall, across the majority of anatomical sites (7 of 11) and across severity-based measures, pain followed an inverted U trajectory rather than a uniform age-related accumulation. This concentration of burden accumulation before midlife across most phenotypes suggests that the window for effective prevention may occur earlier than current clinical and policy frameworks typically assume.

Our study differs from the GBD framework in two key respects. First, GBD primarily estimates the prevalence of clinically defined conditions, such as osteoarthritis (OA) defined by clinical or radiographic criteria^10^ and migraine defined according to International Headache Society diagnostic criteria^11^. For neck pain and low back pain, GBD draws on heterogeneous sources including population surveys, administrative health records using International Classification of Diseases codes, and studies of activity-limiting pain^1,12^. The difference between self-reported and diagnosis-based estimates is particularly relevant as self-reported pain can be captured and compared in settings with limited access to care. A second difference is that GBD estimates are derived from aggregated summary statistics and modelled using Bayesian meta-regression to project estimates even where empirical data are sparse or absent^13,14^. Our analysis instead draws directly on individual-level observations from population-based surveys. As a result, GBD estimates for low back pain and knee OA show a near-linear increase across adulthood until approximately 80 years of age followed by a decline. We instead showed non-linear trajectories across the life course, with the steepest increases occurring between puberty and midlife. Differences were also apparent across regions and levels of national development. Whereas GBD generally reports higher prevalence of low back pain, OA, and migraine in high-income regions, we instead observed higher prevalence of self-reported bodily, joint, back, and headache pain in lower-HDI settings, particularly among older adults. Our findings for neck pain, however, were broadly consistent with GBD estimates, with prevalence peaking around age 50 years and lower levels in high-HDI settings. Overall, by estimating pain irrespective of clinical evaluations, using self-reported data, we provide a different view of burden that is informative in populations with limited health-system access or data monitoring.

Our global reference trajectories provide a benchmark against which historically underrepresented regions can be compared. The consistent pattern of early-life convergence and late-life divergence across regions indicates that regional disparities in pain burden emerge primarily after midlife. The greatest late-life excess was concentrated in eastern Europe and in the regions least represented in current surveillance (i.e., sub-Saharan Africa, central and southern Asia, and north Africa and west Asia) where trajectories rose steeply through midlife to exceed the global reference in older age. This regional patterning was not arbitrary but tracked the gradient of national development: the regions with the steepest late-life trajectories were those at the lower end of the HDI distribution, suggesting that the macro-level conditions HDI captures, including access to care, occupational exposures, and comorbidity burden, shape how pain accumulates across the lifespan. Meanwhile, headache showed the greatest regional heterogeneity, characterised by divergence not only in magnitude but in trajectory shape: the early-adulthood peak and decline characteristic of high-income regions was flatter in east and southeast Asia and absent in sub-Saharan Africa and central and southern Asia, where prevalence increased modestly across the lifespan, pointing to regionally distinct determinants that current data cannot fully resolve. Pain surveillance in children and adolescents remains sparse even in higher-development settings and is largely absent in lower-HDI regions, which may contribute to this heterogeneity and limits the ability to characterise early-life trajectories where they may diverge most. These are among the first population-based lifespan self-reported pain estimates available for these settings, and their divergence from the global reference underscores how heavily current global estimates are shaped by data from wealthier regions.

Modifiable risk factors accounted for a substantial but geographically uneven share of pain burden. In eastern Europe, northern America, western Europe, and Latin America, the combined population attributable fraction of smoking, obesity, and low income reached 22% to 27% of site-specific pain burden, identifying a meaningful preventable component concentrated in settings where these exposures are most prevalent. In sub-Saharan Africa and central and southern Asia, however, these same exposures accounted for less than 13% of burden, indicating that the drivers of pain in the world’s most affected populations are fundamentally different from those that dominate in high-income settings. The likely candidates include occupational physical load^15^, limited access to pain management and rehabilitation^16^, and a greater burden of infectious conditions and nutritional deficiencies ^17,18^. Interventions effective in high-income settings are therefore unlikely to translate directly to lower-HDI contexts, where the drivers of pain burden appear fundamentally distinct and remain poorly characterised.

This work has important limitations. Most contributing datasets were cross-sectional, so the trajectories we present describe population patterns rather than within-person change. Our harmonisation procedure mitigated but could not eliminate misclassification due to heterogeneous recall periods, question wording, and other methodological features. Because responses capture current symptoms, estimates should be interpreted as point or period prevalence rather than persistence or chronicity. Coverage is uneven across regions and across the youngest and oldest ages, which widens uncertainty where data are sparse. Socioeconomic and system indicators such as income and HDI serve as proxies and may not fully capture underlying contextual determinants. Cultural differences in symptom appraisal, survivorship at older ages, and unmeasured clinical factors could still influence absolute levels and slopes despite adjustment. Data coverage for non-musculoskeletal pain in mid-to low-HDI strata was insufficient to derive reliable trajectories, particularly for chest, facial and abdominal pain, which are major contributors to pain burden in younger populations. Improved lifecourse pain surveillance in lower-HDI settings, with standardised measures of pain and early-life adversity, is needed to clarify when and why developmental indices most strongly shape pain trajectories. Taken together, these constraints affect interpretation but do not alter the central life-course patterns seen across body sites.

Our findings establish that self-reported pain follows distinct lifespan trajectories that diverge markedly from condition-based global estimates, with the sharpest divergence in the populations most underrepresented in current surveillance. The reference curves and benchmarking infrastructure introduced here provide a foundation for more equitable global pain monitoring on which future surveillance and prevention efforts can build.

## Methods

### Data sources

We compiled individual-participant data from population-based national, regional, and international health studies and surveys conducted between 1990 and 2025 that assessed self-reported bodily pain (Supplementary Table 1). Studies were eligible if they sampled community-dwelling participants without restricting participation based on health status, assessed pain by direct participant self-report, and included either (i) a pain-presence item at one or more target anatomical sites or (ii) an overall or average pain-intensity rating on a 0–10 numerical rating scale (NRS) or visual analogue scale (VAS), with 0 indicating no pain. Studies were excluded if they relied on clinician, proxy, or observational reports or did not measure any target pain outcome. The final analytic dataset included 6,075,021 individuals with valid data for at least one pain outcome, drawn from 134 study programmes, across 118 countries and territories (figure 1). Study programmes comprised 894 distinct data sources, reflecting multiple survey waves or country-specific samples within multi-country studies.

### Pain outcomes

The primary outcomes were binary indicators of self-reported pain at 11 anatomical sites: head, face, neck or shoulder, foot or ankle, hand or wrist, elbow, chest, back, stomach or abdomen, hip, and knee. Reported pain at these anatomical sites was harmonised by mapping study-specific wording and body-map coding schemes to standardised categories (Supplementary Section 3). Where laterality was assessed, pain was coded as present if reported on either side. A composite ‘any bodily pain’ outcome was derived for studies with sufficient site coverage (Supplementary Section 3) and coded as present if pain was reported at any bodily site.

Secondary outcomes were: (1) high-intensity pain, defined as average pain intensity of 7 or higher on a 0–10 NRS or VAS; and (2) generalised pain, defined as pain present in at least four of five body regions (left upper, right upper, left lower, right lower, and axial), following the spatial criteria of the 2016 fibromyalgia revisions^8^. These secondary outcomes captured complementary dimensions of pain burden beyond site-specific prevalence^6,7,9^.

### Geographic and sociodemographic features

Countries were grouped into nine world regions using a modified United Nations Sustainable Development Goals (SDG) regional classification (Supplementary Table 3) ^19^. National development was indexed using the Human Development Index (HDI), obtained from United Nations Development Programme reports and matched to each participant’s country and survey year^20^. Sex was self-reported within each contributing study.

### Individual risk factors

We evaluated three exposures with established associations with pain outcomes across the literature: body mass index, smoking status, and household income. These exposures were selected because they are modifiable, are consistently associated with musculoskeletal and other pain conditions in population-based research ^21–23^, align with risk factors used in global burden analyses including the Global Burden of Disease (GBD) study^1^, and were sufficiently harmonisable across contributing datasets to enable pooled estimation.

Smoking status was categorised as current (regular or occasional), former, or never. BMI was obtained directly from study-provided values when available or otherwise derived from measured or self-reported height and weight. Adults were classified as having normal weight (18.5–24.9 kg/m^2^), overweight (25.0–29.9 kg/m^2^), or obesity (30.0 kg/m^2^ or higher). For participants aged 19 years or younger, BMI categories were assigned using age- and sex-specific cutoffs based on paediatric growth standards from the World Health Organization^24^. Household income was harmonised within each study by ranking participants into study-specific income quintiles (equal-frequency groups), reflecting relative socioeconomic position within studies, consistent with established inequality-monitoring practice^25–27^.

### Harmonisation procedure

Pain prevalence estimates vary with methodological factors such as interview setting, question phrasing, recall period, and study design characteristics^28–30^. To address this, our harmonisation procedure systematically evaluated seven key methodological features for their association with reported pain: recall period, case definition (general versus chronic-explicit), survey format, survey setting, question valence, question structure, and sampling strategy (see Methods and Supplementary Section 6.2). Recall period showed the most consistent and substantial association with reported pain and was included in all models (short: past week; medium: past month; long: past 3–12 months). We additionally coded case definition as general versus chronic-explicit, where chronic-explicit indicated that the question specified pain lasting longer than 3 months; this covariate was included to adjust for the small subset of studies using chronic-explicit wording. The remaining five features (*i*.*e*., survey format, survey setting, question valence, question format, and sampling strategy), had small or site-specific effects (±2 percentage points) that did not alter age-by-site patterns and were not retained to preserve parsimony. Sensitivity analyses on the stability of global trajectories across studies (Supplementary Section 6.1), methodological covariates (Supplementary Section 6.2) and recall periods (Supplementary Section 6.3), random-effects structures (Supplementary Section 6.4), functional form (Supplementary Section 6.5), and data missing (Supplementary Section 6.6) show the robustness of our harmonisation procedure.

### Statistical analysis

We estimated age-specific prevalence trajectories for each pain outcome across the lifespan (ages 5 to 100 or older; ages below 5 and above 100 were bottom- and top-coded) using generalized linear mixed-effects models (GLMMs) with a logit link. Age was modelled using natural cubic splines to flexibly capture nonlinear trajectories. The spline degrees of freedom were selected using study-clustered 5-fold cross-validation; 3 degrees of freedom were retained based on out-of-sample fit (Supplementary Section 6.5). We report 95% confidence intervals throughout.

Primary models included an interaction between age and the exposure of interest, namely sex (figures 2,3), world regions and HDI (figure 4), BMI category, smoking status, or income quintile (figure 5, Extended Data Figs. 1–4), and adjusted for methodological differences using fixed effects for recall period and case definition. All models additionally adjusted for sex, except those in which sex was the primary exposure. Random intercepts for study programme and world region were included to account for clustering and enable partial pooling across heterogeneous data sources. In regional analyses, region was instead treated as a fixed effect with a study-nested-within-region random intercept, preventing any single multi-country study from dominating region-specific estimates. Sensitivity analyses for missing risk-factor data are reported in Supplementary Section 6.6.

Adjusted prevalence trajectories were obtained as marginal predictions and standardised to a past-month recall period and the general case definition (Supplementary Section 5). HDI trajectories were plotted at four representative values (0.55, 0.70, 0.80, 0.90) spanning low to very high development. We also quantified the rate of age-related change as the year-to-year difference in predicted prevalence (percentage-point change per year). All analyses were performed in R (version 4.4.1).

## Supporting information

Supplemental methods and analyses

## Data Availability

An interactive benchmarking tool and downloadable summary statistics from this study are publicly available at https://evppainlab.shinyapps.io/global-pain-benchmark/. Researchers seeking access to the original individual-level data should apply directly to the respective data custodians according to their access policies.

## Acknowledgements

This work was supported by the Canadian Institutes of Health Research (RN441786–453096), the Fonds de recherche du Québec en Santé (283687), the Réseau québécois de recherche sur la douleur, and the Louise and Alan Edwards Grants in Pain Research (to E.V.-P.); by the Louise and Alan Edwards Foundation (to M.F.). We are deeply grateful to all study participants, investigators, data managers, and field teams whose efforts made this work possible. We thank our collaborators and data providers for sharing data and expertise and for their critical contributions to the Lifespan Pain Project. We also thank the McGill Office of Sponsored Research, particularly Ji Eun Lee, for her assistance in coordinating data transfer agreements and institutional contracts that enabled this international collaboration.

