## Extended data figures 1-7 for "Pain across the lifespan: global and regional reference curves from 6.1 million individuals in 118 countries"

Matt Fillingim (McGill University, Canada)*; Christophe Tanguay-Sabourin (University of Montreal, Canada); Lindsay Neuert (McGill University, Canada); Azin Zare (McGill University, Canada); Jax Norman (McGill University, Canada); Gianluca V. Guglietti (McGill University, Canada); Lise Hobeika (McGill University, Canada); Abimbola Ayorinde (University of Warwick, UK); Ahmad Salimzadeh (Tehran University of Medical Sciences, Iran); Ahmad-reza Jamshidi (Tehran University of Medical Sciences, Iran); Arash Tehrani-Banihashemi (Tehran University of Medical Sciences, Iran); Beatriz Olaya (Universitat Autònoma de Barcelona, Spain); Benjamin Longo Mbenza (Université de Kinshasa, Democratic Republic of the Congo); Bibilola Oladeji (University of Ibadan, Nigeria); Brenda Penninx (Amsterdam UMC, The Netherlands); Buddhi Paudyal (Patan Academy of Health Sciences, Nepal); Caique de Melo (Universidade Cidade de São Paulo, Brazil); Chantal Berna-Renella (University of Lausanne, Switzerland); Clarissa Humberg (Witten/Herdecke University, Germany); Claude-Pierre Pascal (Université Paris Est Créteil, France); Elisa Smith (University of Aberdeen, UK); Elizabeth VanDenKerkhof (Queen's University, Canada); Erik J. Giltay (Leiden University Medical Center, The Netherlands); Esther Garcia-Esquinas (Instituto de Salud Carlos III, Spain); Faisal Parlindungan (Universitas Indonesia, Indonesia); Fatima Qayyum Abbasi (Pakistan Institute of Medical Sciences, Islamabad, Pakistan); Fernando Rodríguez Artalejo (Universidad Autónoma de Madrid, Spain); Gareth T Jones (University of Aberdeen, UK); Gary Slade (University of North Carolina at Chapel Hill, USA); Hanifa Bouziri (Université Paris-Cité, France); Herta Flor (Central Institute of Mental Health, Mannheim, Germany; Heidelberg University, Germany); Hiroyasu Iso (Japan Institute for Health Security, Japan); Ian Gilron (Queen's University, Canada); Imad Uthman (American University of Beirut, Lebanon); Ingris Peláez-Ballestas (Hospital General de Mexico Dr. Eduardo Liceaga, Mexico); Jean-Paul Devengi Nzambi (Université de Kinshasa, Democratic Republic of the Congo); Jose Luis Ayuso-Mateos (Universidad Autónoma de Madrid, Spain); Josep Maria Haro (Parc Sanitari Sant Joan de Déu, Spain); Julia Wager (Witten/Herdecke University, Germany); Juliana Barcellos de Souza (Universidade do Estado de Santa Catarina (UDESC), Brazil); Kazutoshi Nakamura (Niigata University Graduate School of Medical and Dental Sciences, Japan); Keiko Yamada (Juntendo University Graduate School of Medicine, Japan); Lisa Marie Rau (Witten/Herdecke University, Germany); Makram Talih (Trinity College Dublin, Ireland); Marcus J Beasley (University of Aberdeen, UK); María Dueñas (Instituto de Saúde Pública da Universidade do Porto, Portugal); Marie Zins (Université Paris-Cité, France); Marta Miret (Universidad Autónoma de Madrid, Spain); Melissa Wake (The University of Melbourne, Australia); Mirza Zaka Pratama (King Salman Armed Forces Hospital, Saudi Arabia); Mohammad Almalki (Hamad Medical Corporation, Qatar); Monique Chaaya (American University of Beirut, Lebanon); Muhammad Anshory (King Salman Armed Forces Hospital, Saudi Arabia); Naoki Kondo (Kyoto University, Japan); Navid Moghadam (Tehran University of Medical Sciences, Iran); Norma Mansor (Universiti Malaya, Malaysia); Omar Alsaed (College of Medical Technology, Libya); Orawan Keeratisiroj (Faculty of Public Health, Naresuan University, Thailand); Oye Gureje (University of Ibadan, Nigeria); Pande Ketut Kurniari (Universitas Udayana, Indonesia); Pardis Noormohammadpour (Tehran University of Medical Sciences, Iran); Pierrot Lebughe (Université de Kinshasa, Democratic Republic of the Congo); Raga A Elzahaf (Changchun University of Chinese Medicine, China); Ramin Kordi (Tehran University of Medical Sciences, Iran); Raquel Lucas (Universidade do Porto, Portugal); Rashmi Parekh Bhandari (Stanford University Medical Center, USA); Rudy Hidayat (Universitas Indonesia, Indonesia); Saba Samreen (Foundation University Islamabad, Pakistan); Samar Al-Emadi (College of Medical

Technology, Libya); Shima RoknSharifi (Tehran University of Medical Sciences, Iran); Shinsuke Inoue (Kaguyama Orthopaedic Clinic, Japan); Sudeep Adhikari (Oxford University Clinical Research Unit, Nepal); Suryo Anggoro Kusumo Wibowo (Universitas Indonesia, Indonesia); Sweekriti Sharma (The University of Sydney, Australia); Sylvia Villeneuve (McGill University, Canada); Takahiro Tabuchi (Tohoku University Graduate School of Medicine, Japan); Tayyeba Khursheed (Pakistan Institute of Medical Sciences, Islamabad, Pakistan); Tie Yamato (The University of Sydney, Australia); Todd Jackson (University of Macau, China); Veronica Souza Santos (McMaster University, Canada); Wantana Siritaratiwat (Khon Kaen University, Thailand); Xianchen Liu (Shandong University School of Public Health, China); Zhen-Zhen Liu (University of Tubingen, Germany); Tobias Banaschewski (IMAGEN; Heidelberg University, Germany); Gareth J. Barker (IMAGEN; King's College London, UK); Arun L.W. Bokde (IMAGEN; Ludwig-Maximilians-Universitat, Germany); Rudiger Bruhl (IMAGEN; Technische Universitat Dresden, Germany); Sylvane Desrivieres (IMAGEN; King's College London, UK); Penny Gowland (IMAGEN; University of Nottingham, UK); Antoine Grigis (IMAGEN; Universite Paris-Saclay, France); Andreas Heinz (IMAGEN; Charite Universitatsmedizin Berlin, Germany); Herve Lemaitre (IMAGEN; Universite Paris-Saclay, France); Jean-Luc Martinot (IMAGEN; Universite Paris-Saclay, France); Marie-Laure Paillere Martinot (IMAGEN; Universite Paris-Saclay, France); Eric Artiges (IMAGEN; Universite Paris-Saclay, France); Frauke Nees (IMAGEN; Physikalisch-Technische Bundesanstalt, Germany); Dimitri Papadopoulos Orfanos (IMAGEN; Universite Paris-Saclay, France); Tomas Paus (IMAGEN; University of Montreal, Canada); Luise Poustka (IMAGEN; University Hospital Heidelberg, Germany); Michael N. Smolka (IMAGEN; Heidelberg University, Germany); Sarah Hohmann (IMAGEN; Heidelberg University, Germany); Nilakshi Vaidya (IMAGEN; University of Tubingen, Germany); Henrik Walter (IMAGEN; University of Tubingen, Germany); Robert Whelan (IMAGEN; Ludwig-Maximilians-Universitat, Germany); Paul Wirsching (IMAGEN; University of Tubingen, Germany); Gunter Schumann (IMAGEN; University of Tubingen, Germany); Susan M Sawyer (The University of Melbourne, Australia); Saman Khalatbari-Soltani (The University of Sydney, Australia); Olof Anna Steingrimsdottir (Norwegian Institute of Public Health, Norway); Gary J Macfarlane (University of Aberdeen, UK); Fiona Blyth (The University of Sydney, Australia); Etienne Vachon-Presseau (McGill University, Canada)*.

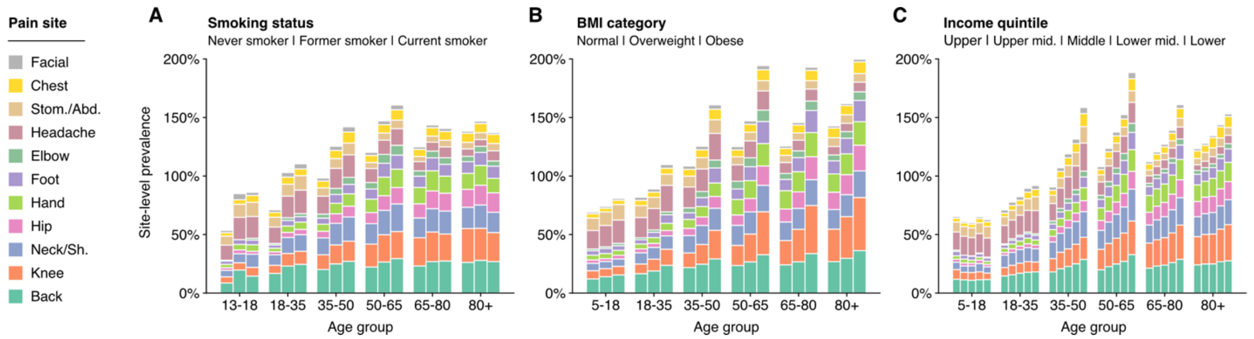


**Extended Data Figure 1: Pain burden across the lifespan by smoking, BMI, and household income.**
Stacked bars show age-group specific, site-level prevalence (percent) for each anatomical pain site, stratified by (A) smoking status (never smoker, former smoker, current smoker); (B) BMI category (normal weight, overweight, obese); and (C) household income quintile (upper to lower). Colours denote pain sites. Because individuals can report pain at multiple sites, site-level prevalences are not mutually exclusive and stacked totals can exceed 100%. Age groups are shown on the x-axis; smoking results are shown from ages 13 and older.

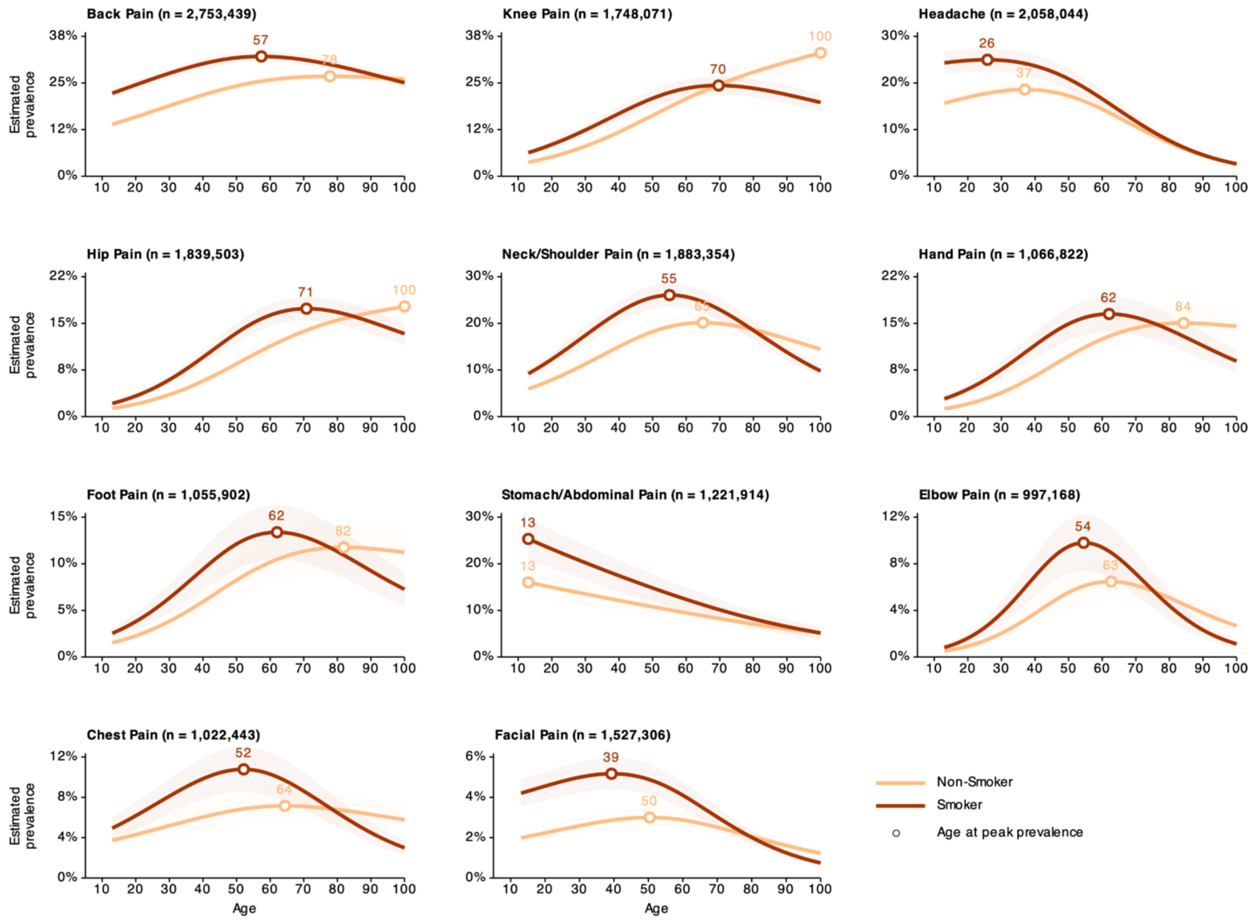


**Extended Data Figure 2: Prevalence trajectories by smoking status.** Estimated prevalence of 11 pain sites across the lifespan by smoking status (smoker vs non-smoker). Shaded bands indicate 95% confidence intervals. Open circles indicate age at peak prevalence.

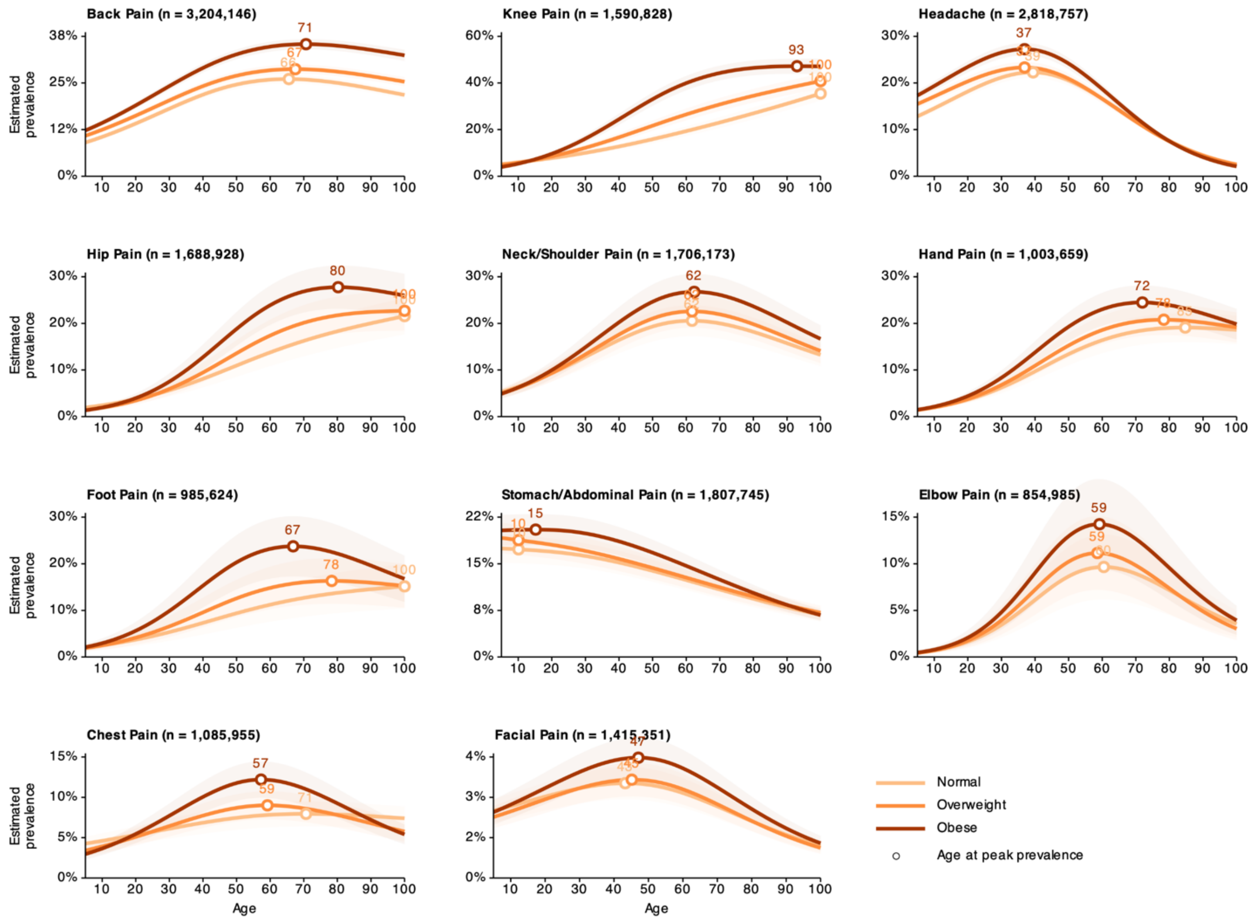


**Extended Data Figure 3: Prevalence trajectories by BMI category.** Estimated prevalence of 11 pain sites across the lifespan by BMI category (normal, overweight, obese). Shaded bands indicate 95% confidence intervals. Open circles indicate age at peak prevalence.

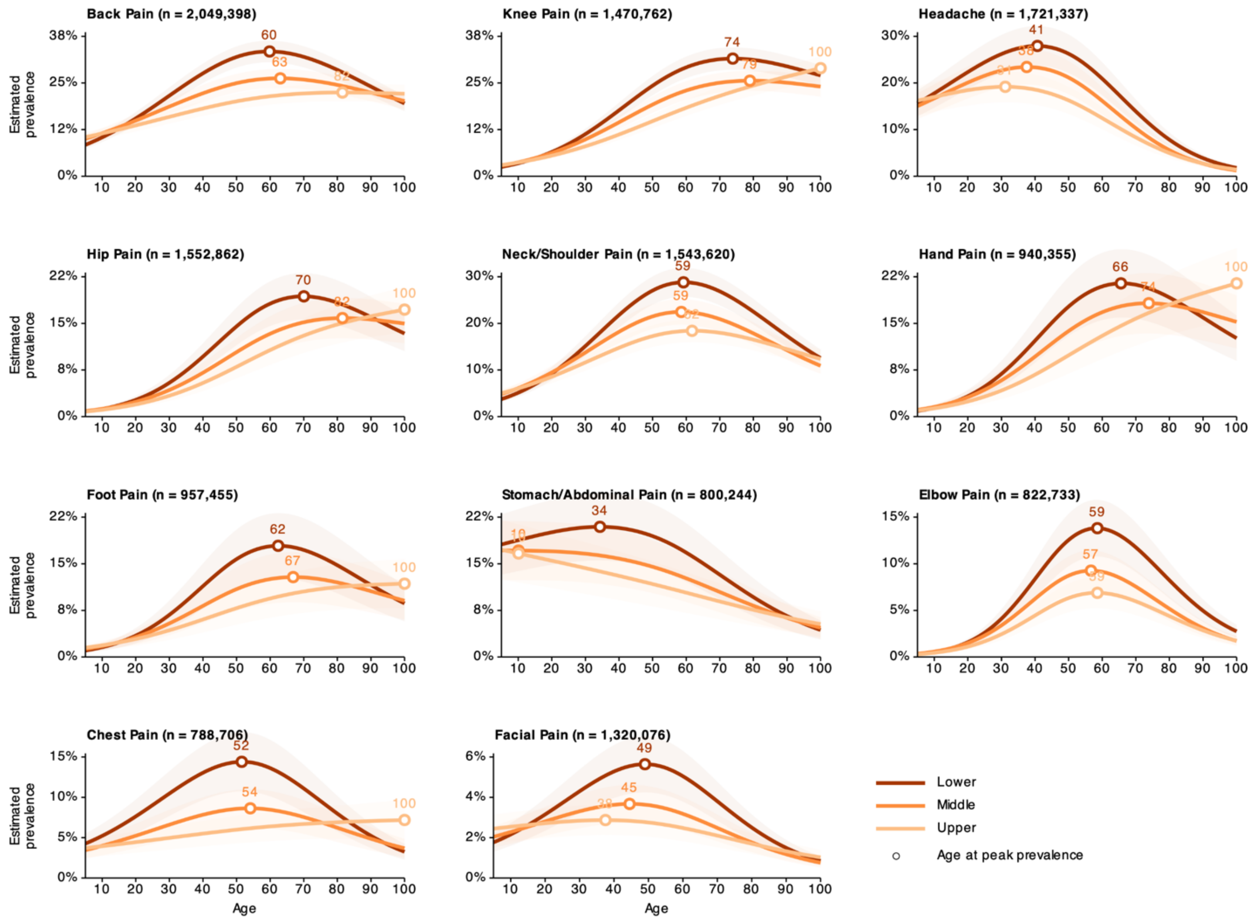


**Extended Data Figure 4: Prevalence trajectories by household income level.** Estimated prevalence of 11 pain sites across the lifespan by three representative household income quintiles (lower, middle, upper). Income was harmonised as study-specific relative rank (equal-frequency quintiles), reflecting within-study socioeconomic position. Shaded bands indicate 95% confidence intervals. Open circles indicate age at peak prevalence.

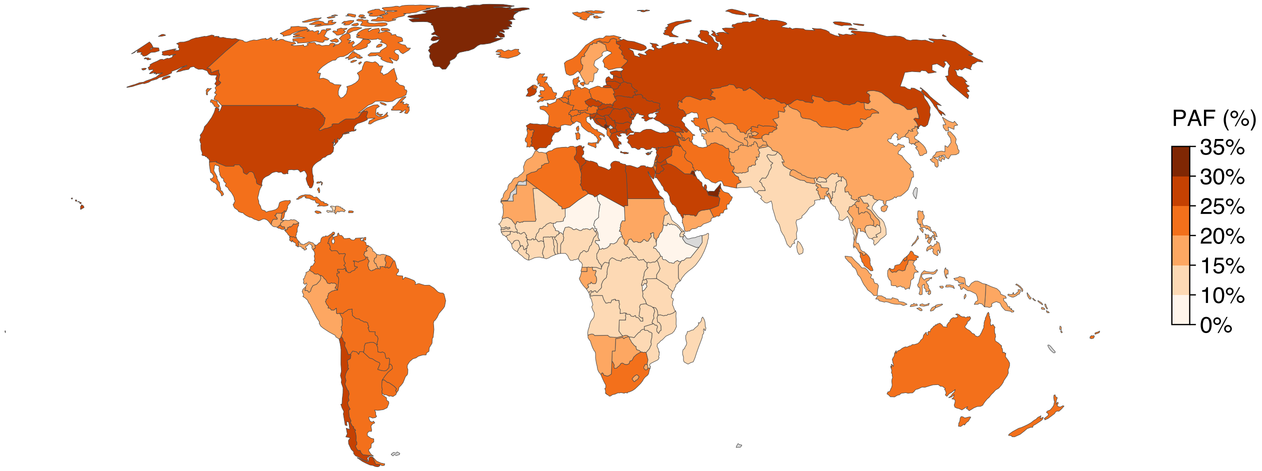


**Extended Data Figure 5: Combined population attributable fraction for pain by country.** Country-level combined population attributable fraction (PAF, %) representing the proportion of site-specific pain burden jointly attributable to obesity, smoking, and low household income. Age- and sex-specific PAFs were computed at the country level by combining study-derived risk ratios with country-specific exposure prevalence from the Global Burden of Disease study (obesity: GBD 2021; smoking: GBD 2015) and population denominators. Income was operationalised as within-study relative rank (quintiles), with a fixed prevalence of 20% per quintile across all countries. Values are case-weighted averages across 11 anatomical pain sites. Grey indicates countries without available exposure data.

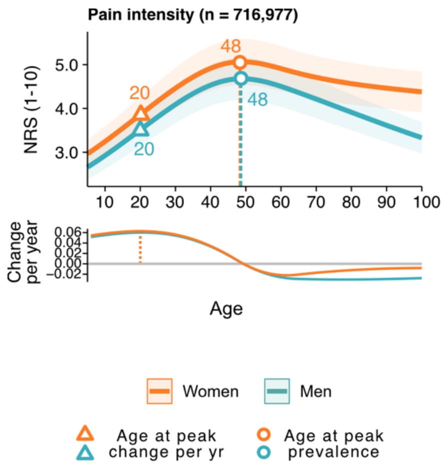


**Extended Data Figure 6: Average pain intensity across the lifespan by sex.** Upper panel: model-estimated average pain intensity (numerical rating scale, 1–10) among individuals reporting pain (n = 716,977), by age and sex. Shaded bands indicate 95% confidence intervals. Open circles indicate the age at peak intensity; open triangles indicate the age of most rapid increase. Lower panel: rate of change in predicted intensity (NRS points per year), with the grey horizontal line indicating zero change.

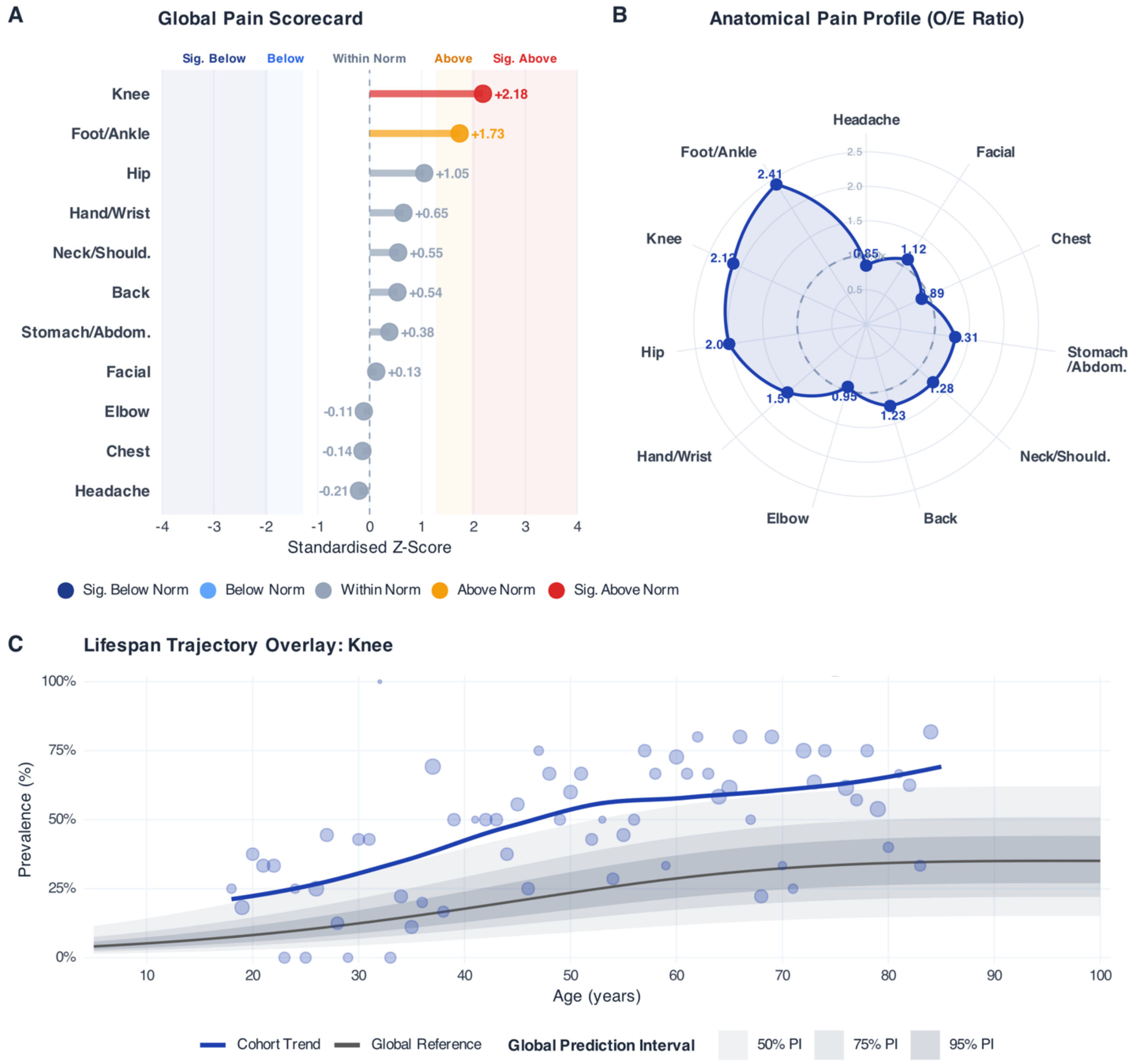


**Extended Data Figure 7: Representative output of the Global Lifespan Pain Benchmarking Tool applied to a hypothetical external cohort.** The tool benchmarks external cohort data against the global normative reference derived from the primary analysis (see Supplementary Section 10 for full statistical methodology and application architecture). (A) Global Pain Scorecard: lollipop chart of standardised Z-scores for 11 anatomical pain sites, with colour-coded deviation zones (Significantly Below Norm, $|Z| \geq 1.96$; Below Norm, $1.28 \leq |Z| < 1.96$; Within Norm, $|Z| < 1.28$; Above Norm; Significantly Above Norm). (B) Anatomical Pain Profile: radar chart mapping O/E ratios across all 11 sites in anatomical order (head to foot). (C) Lifespan Trajectory Overlay: age-prevalence chart for knee pain superimposing the cohort's observed LOESS trend (blue) against the global reference trajectory (grey) and its 50%, 75%, and 95% prediction intervals derived from between-study heterogeneity on the logit scale. Points are sized by sample size per age. The tool is freely accessible at https://evppainlab.shinyapps.io/global-pain-benchmark/.
