## Supplemental methods and analyses for "Pain across the lifespan: global and regional reference curves from 6.1 million individuals in 118 countries"

### Section 1. Global Pain Assessment Database

#### *Collaboration overview*

The present study was made possible by an international collaboration of investigators who contributed individual participant data from population-based surveys and cohort studies. The collaboration was established to map reference curves of pain across the human lifespan and to quantify how these trajectories differ by sociodemographic (sex and total household income), health features (smoking status and BMI), and broader environment (world region and country-level Human Development Index).

### Section 2. Data sources

#### *2.1 Data access*

We compiled individual level data from a wide range of population based national, regional, and international health studies and surveys conducted between 1990 and 2025 that assessed self-reported bodily pain. Major sources included surveys coordinated by the World Health Organization, the Community Oriented Program for Control of Rheumatic Diseases (COPCORD), the National Health Interview Surveys (NHIS), and the United Kingdom Biobank (UKB), alongside additional national and regional population-based studies. A full list of data sources is provided in Supplementary Table 1.

### Supplementary Table 1: Characteristics of included data sources

| No. | Study name | Country | Years of data collection | N (analytic sample) | Age range (years) | Survey type | Pain sites collected | Risk factors analyzed |
|---|---|---|---|---|---|---|---|---|
| 1 | Adolescent Brain Cognitive Development Study (ABCD) | United States | 2017 | 10843 | 11–14 | Cohort | P, Ha, N/S, C, S/A, B, K, H, F, Hnd, Ft, E | BMI; Income |
| 2 | Agincourt Integrated Family Survey (AIFS) | South Africa | 2004 | 1507 | 18–92 | Surveillance/HDSS | P, Ha, C, S/A, B | BMI; Income |
| 3 | Alameda County Health and Ways of Living Study (ACHWLS) | United States | 1994 | 2729 | 46–100 | Panel | Ha, C, S/A, B | Smoking; Income |
| 4 | All of Us Research Program (AoU) | United States | 2018 | 520906 | 18–87 | Cohort | P | BMI; Smoking; Income |
| 5 | Australian Longitudinal Study on Aging (ALSA) | Australia | 1992 | 2080 | 65–100 | Cohort | P, Ha, C, B, K, H, Hnd, Ft, E | Smoking; Income |
| 6 | Austrian Health Information Survey (ATHIS) | Austria | 2006 | 15474 | 17–87 | Household survey | P, Ha, N/S, C, S/A, B, K, H, F, Hnd, Ft | BMI; Smoking; Income |
| 7 | Bangladesh Urban Health Survey (BUHS) | Bangladesh | 2006 | 28010 | 13–59 | Household survey | Ha | BMI; Income |
| 8 | Brazil National Survey of Health (PNS) | Brazil | 2013 | 60202 | 18–100 | Household survey | B | BMI; Income |
| 9 | Brazil National Survey of Health (PNS) | Brazil | 2019 | 90846 | 15–100 | Household survey | B | BMI; Income |
| 10 | Brazilian Children's Musculoskeletal Pain Survey (BCS) | Brazil | 2020 | 1988 | 6–18 | School-based survey | P, N/S, B | BMI |
| 11 | Brazilian Longitudinal Study of Aging (ELSI) | Brazil | 2019 | 9949 | 50–100 | Household survey | P, Ha, N/S, F | BMI; Smoking |
| 12 | Brazilian Pain Survey | Brazil | 2015 | 723 | 16–75 | Household survey | P, Ha, N/S, C, S/A, B, K, H, F, Hnd, Ft | |
| 13 | COPCORD | Argentina | 2011 | 644 | 18–85 | Community survey | P, N/S, B, K, H, Hnd, Ft, E | |
| 14 | COPCORD | Colombia | 2014 | 6690 | 15–99 | Community survey | P, N/S, B, K, H, Hnd, Ft, E | Smoking; Income |
| 15 | COPCORD | Democratic Republic of the Congo | 2012 | 1315 | 7–88 | Community survey | P, N/S, B, K, H, Hnd, Ft, E | BMI |
| 16 | COPCORD | Ecuador | 2014 | 4867 | 18–97 | Community survey | P, N/S, B, K, H, Hnd, Ft, E | Income |
| 17 | COPCORD | Ecuador | 2016 | 2681 | 18–97 | Community survey | P, N/S, B, K, H, Hnd, Ft, E | Income |
| 18 | COPCORD | Indonesia | 2023 | 3898 | 16–100 | Community survey | P, N/S, B, K, Hnd, Ft, E | BMI; Smoking |
| 19 | COPCORD | Iran | 2004 | 10288 | 15–100 | Community survey | P, N/S, B, K, H, Hnd, Ft, E | BMI |
| 20 | COPCORD | Iran | 2008 | 2099 | 15–84 | Community survey | P, N/S, B, K, H, Hnd, Ft, E | BMI |
| 21 | COPCORD | Iran | 2011 | 2654 | 15–100 | Community survey | P, N/S, B, K, H, Hnd, Ft, E | |
| 22 | COPCORD | Iran | 2014 | 2000 | 15–90 | Community survey | P, N/S, B, K, H, Hnd, Ft, E | BMI |
| 23 | COPCORD | Lebanon | 2009 | 3517 | 15–90 | Community survey | P, N/S, B, K, H, Hnd, Ft | BMI; Smoking; Income |
| 24 | COPCORD | Mexico | 2012 | 23181 | 17–100 | Community survey | P, N/S, B, K, H, Hnd, Ft | Income |
| 25 | COPCORD | Nepal | 2021 | 2143 | 18–95 | Community survey | P, N/S, B, K, H, Hnd, Ft, E | Smoking; Income |

| | | | | | | | | |
|---|---|---|---|---|---|---|---|---|
| 26 | COPCORD | Pakistan | 2018 | 5495 | 12–99 | Community survey | P, N/S, B, K, Hnd, Ft | Smoking; Income |
| 27 | COPCORD | Pakistan | 2024 | 7336 | 18–100 | Community survey | P, N/S, B, K, H, Hnd, Ft, E | |
| 28 | COPCORD | Peru | 2010 | 3036 | 17–100 | Community survey | P, N/S, B, K, H, Hnd, Ft, E | BMI; Income |
| 29 | COPCORD | Qatar | 2019 | 1225 | 15–96 | Community survey | P, N/S, B, K, H, Hnd, Ft | BMI |
| 30 | COPCORD | Venezuela | 2011 | 3974 | 17–100 | Community survey | P, N/S, B, K, Hnd, Ft, E | |
| 31 | Cape Area Panel Study (CAPS) | South Africa | 2006 | 2248 | 47–97 | Panel | P, Ha, C, S/A, B | BMI; Income |
| 32 | Childhood to Adolescence Transition Study (CATS) | Australia | 2012 | 1191 | 8–11 | Cohort | P, Ha, N/S, C, S/A, B, K, H, Hnd, Ft, E | BMI; Income |
| 33 | Chilean Gender and Sexuality Survey (ENSSEX) | Chile | 2022 | 20374 | 18–100 | Household survey | P, Ha | Income |
| 34 | Chilean Health Survey (ENS) | Chile | 2003 | 3557 | 16–99 | Household survey | P, N/S, B, K, H, Hnd, Ft, E | BMI; Smoking; Income |
| 35 | Chilean Health Survey (ENS) | Chile | 2009 | 5276 | 15–100 | Household survey | P, N/S, C, B, K, H, Hnd, Ft, E | BMI; Smoking; Income |
| 36 | Chilean Health Survey (ENS) | Chile | 2016 | 6233 | 15–98 | Household survey | P, N/S, B, K, H, F, Hnd, Ft, E | BMI; Smoking; Income |
| 37 | Chilean Workers Survey (ENETS) | Chile | 2009 | 9501 | 15–99 | Household survey | P, Ha, N/S, B, K, H, Hnd, Ft, E | Income |
| 38 | China Health and Retirement Longitudinal Study (CHARLS) | China | 2011 | 24353 | 21–100 | Panel | P, Ha, N/S, C, S/A, B, K, H, Hnd, Ft | BMI; Smoking; Income |
| 39 | Chongqing Pain Survey | China | 2012 | 992 | 15–90 | Household survey | P, Ha, N/S, C, S/A, B, F | |
| 40 | Chronic Headache in Adolescents (CHAP) | Germany | 2017 | 2280 | 10–18 | Cohort | P, Ha, C, S/A, B | BMI; Income |
| 41 | CoLaus | Switzerland | 2015 | 3590 | 45–87 | Cohort | P, N/S, B, K, H, F, Hnd, Ft, E | Smoking; Income |
| 42 | Collaborative Psychiatric Epidemiology Surveys (CPES) | United States | 2001 | 4645 | 18–97 | Cross-section | P, Ha, C, S/A, B | BMI; Smoking; Income |
| 43 | Community-Led Total Sanitation and Hygiene in Ethiopia (CLTSH) | Ethiopia | 2017 | 4726 | 5–80 | Program/Household survey | Ha | |
| 44 | Constances[1] | France | 2016 | 29728 | 24–80 | Cohort | P, N/S, B, K, Hnd, E | BMI; Smoking |
| 45 | Cross Marketing Survey (CMS) | Japan | 2022 | 1946 | 20–79 | Community survey | P, N/S, B, K, H, Hnd, Ft, E | |
| 46 | DOLORisk | United Kingdom | 2016 | 7236 | 24–99 | Cohort | P, Ha, N/S, C, S/A, B, K, H, F, Hnd, Ft | |
| 47 | English Longitudinal Study on Aging (ELSA) | United Kingdom | 2006 | 2777 | 19–99 | Panel | P, C, B, K, H, F, Ft | Smoking; Income |
| 48 | English Longitudinal Study on Aging (ELSA) | United Kingdom | 2008 | 3277 | 24–99 | Panel | P, C, B, K, H, F, Ft | BMI; Smoking; Income |
| 49 | English Longitudinal Study on Aging (ELSA) | United Kingdom | 2010 | 301 | 35–88 | Panel | P, C, B, K, H, F, Ft | Smoking; Income |
| 50 | English Longitudinal Study on Aging (ELSA) | United Kingdom | 2012 | 8119 | 31–89 | Panel | P, B, K, H, F, Ft | BMI; Smoking; Income |

| | | | | | | | | |
|---|---|---|---|---|---|---|---|---|
| 51 | English Longitudinal Study on Aging (ELSA) | United Kingdom | 2014 | 524 | 29–88 | Panel | P, B, K, H, F, Ft | Smoking; Income |
| 52 | English Longitudinal Study on Aging (ELSA) | United Kingdom | 2016 | 40 | 37–79 | Panel | P, B, K, H, F, Ft | Smoking; Income |
| 53 | English Longitudinal Study on Aging (ELSA) | United Kingdom | 2018 | 1156 | 30–82 | Panel | P, B, K, H, F, Ft | Smoking; Income |
| 54 | English Longitudinal Study on Aging (ELSA) | United Kingdom | 2020 | 1872 | 26–88 | Panel | P, B, K, H, F, Ft | Smoking; Income |
| 55 | Epidemiology of Neuropathic Pain in Canada (ENPC) | Canada | 2017 | 1483 | 5–93 | Cross-section | P, Ha, N/S, C, B, K, H, Hnd, Ft, E | Smoking; Income |
| 56 | Eurobarometer | Austria | 2006 | 1008 | 15–92 | Repeat cross-section | P, Ha, N/S, B, K, H, Hnd, Ft, E | Smoking |
| 57 | Eurobarometer | Belgium | 2006 | 1010 | 15–93 | Repeat cross-section | P, Ha, N/S, B, K, H, Hnd, Ft, E | Smoking |
| 58 | Eurobarometer | Bulgaria | 2006 | 1016 | 15–91 | Repeat cross-section | P, Ha, N/S, B, K, H, Hnd, Ft, E | Smoking |
| 59 | Eurobarometer | Croatia | 2006 | 998 | 15–89 | Repeat cross-section | P, Ha, N/S, B, K, H, Hnd, Ft, E | Smoking |
| 60 | Eurobarometer | Cyprus | 2006 | 1006 | 15–96 | Repeat cross-section | P, Ha, N/S, B, K, H, Hnd, Ft, E | Smoking |
| 61 | Eurobarometer | Czech Republic | 2006 | 1047 | 15–87 | Repeat cross-section | P, Ha, N/S, B, K, H, Hnd, Ft, E | Smoking |
| 62 | Eurobarometer | Denmark | 2006 | 1060 | 15–93 | Repeat cross-section | P, Ha, N/S, B, K, H, Hnd, Ft, E | Smoking |
| 63 | Eurobarometer | Estonia | 2006 | 989 | 15–94 | Repeat cross-section | P, Ha, N/S, B, K, H, Hnd, Ft, E | Smoking |
| 64 | Eurobarometer | Finland | 2006 | 1023 | 15–95 | Repeat cross-section | P, Ha, N/S, B, K, H, Hnd, Ft, E | Smoking |
| 65 | Eurobarometer | France | 2006 | 1014 | 15–95 | Repeat cross-section | P, Ha, N/S, B, K, H, Hnd, Ft, E | Smoking |
| 66 | Eurobarometer | Germany | 2006 | 1537 | 15–94 | Repeat cross-section | P, Ha, N/S, B, K, H, Hnd, Ft, E | Smoking |
| 67 | Eurobarometer | Greece | 2006 | 999 | 15–91 | Repeat cross-section | P, Ha, N/S, B, K, H, Hnd, Ft, E | Smoking |
| 68 | Eurobarometer | Hungary | 2006 | 993 | 15–92 | Repeat cross-section | P, Ha, N/S, B, K, H, Hnd, Ft, E | Smoking |
| 69 | Eurobarometer | Ireland | 2006 | 1294 | 15–95 | Repeat cross-section | P, Ha, N/S, B, K, H, Hnd, Ft, E | Smoking |
| 70 | Eurobarometer | Italy | 2006 | 999 | 15–84 | Repeat cross-section | P, Ha, N/S, B, K, H, Hnd, Ft, E | Smoking |
| 71 | Eurobarometer | Latvia | 2006 | 986 | 15–74 | Repeat cross-section | P, Ha, N/S, B, K, H, Hnd, Ft, E | Smoking |
| 72 | Eurobarometer | Lithuania | 2006 | 992 | 15–95 | Repeat cross-section | P, Ha, N/S, B, K, H, Hnd, Ft, E | Smoking |
| 73 | Eurobarometer | Luxembourg | 2006 | 497 | 15–90 | Repeat cross-section | P, Ha, N/S, B, K, H, Hnd, Ft, E | Smoking |
| 74 | Eurobarometer | Malta | 2006 | 499 | 15–89 | Repeat cross-section | P, Ha, N/S, B, K, H, Hnd, Ft, E | Smoking |
| 75 | Eurobarometer | Netherlands | 2006 | 1063 | 15–92 | Repeat cross-section | P, Ha, N/S, B, K, H, Hnd, Ft, E | Smoking |
| 76 | Eurobarometer | Poland | 2006 | 976 | 15–91 | Repeat cross-section | P, Ha, N/S, B, K, H, Hnd, Ft, E | Smoking |
| 77 | Eurobarometer | Portugal | 2006 | 991 | 15–98 | Repeat cross-section | P, Ha, N/S, B, K, H, Hnd, Ft, E | Smoking |

| | | | | | | | | |
|---|---|---|---|---|---|---|---|---|
| 78 | Eurobarometer | Romania | 2006 | 1016 | 15–91 | Repeat cross-section | P, Ha, N/S, B, K, H, Hnd, Ft, E | Smoking |
| 79 | Eurobarometer | Slovakia | 2006 | 1167 | 15–91 | Repeat cross-section | P, Ha, N/S, B, K, H, Hnd, Ft, E | Smoking |
| 80 | Eurobarometer | Slovenia | 2006 | 1036 | 15–91 | Repeat cross-section | P, Ha, N/S, B, K, H, Hnd, Ft, E | Smoking |
| 81 | Eurobarometer | Spain | 2006 | 1022 | 15–94 | Repeat cross-section | P, Ha, N/S, B, K, H, Hnd, Ft, E | Smoking |
| 82 | Eurobarometer | Sweden | 2006 | 1005 | 15–92 | Repeat cross-section | P, Ha, N/S, B, K, H, Hnd, Ft, E | Smoking |
| 83 | Eurobarometer | United Kingdom | 2006 | 1061 | 15–95 | Repeat cross-section | P, Ha, N/S, B, K, H, Hnd, Ft, E | Smoking |
| 84 | European Health Survey in Spain (EESE) | Spain | 2014 | 22840 | 15–100 | Household survey | P, Ha, N/S, B | BMI; Smoking; Income |
| 85 | European Health Survey in Spain (EESE) | Spain | 2020 | 22069 | 15–100 | Household survey | P, Ha, N/S, B | BMI; Smoking; Income |
| 86 | European Social Survey (ESS) | Austria | 2014 | 4143 | 15–96 | Repeat cross-section | P, Ha, B | BMI; Smoking; Income |
| 87 | European Social Survey (ESS) | Belgium | 2014 | 3358 | 15–93 | Repeat cross-section | P, Ha, B | BMI; Smoking; Income |
| 88 | European Social Survey (ESS) | Bulgaria | 2014 | 2236 | 16–90 | Repeat cross-section | P, Ha, B | BMI; Smoking; Income |
| 89 | European Social Survey (ESS) | Croatia | 2014 | 1554 | 15–90 | Repeat cross-section | P, Ha, B | BMI; Smoking; Income |
| 90 | European Social Survey (ESS) | Cyprus | 2014 | 666 | 17–90 | Repeat cross-section | P, Ha, B | BMI; Smoking; Income |
| 91 | European Social Survey (ESS) | Czech Republic | 2014 | 2117 | 15–88 | Repeat cross-section | P, Ha, B | BMI; Smoking; Income |
| 92 | European Social Survey (ESS) | Denmark | 2014 | 1502 | 15–100 | Repeat cross-section | P, Ha, B | BMI; Smoking; Income |
| 93 | European Social Survey (ESS) | Finland | 2014 | 3650 | 15–100 | Repeat cross-section | P, Ha, B | BMI; Smoking; Income |
| 94 | European Social Survey (ESS) | France | 2014 | 3682 | 15–99 | Repeat cross-section | P, Ha, B | BMI; Smoking; Income |
| 95 | European Social Survey (ESS) | Germany | 2014 | 5447 | 15–100 | Repeat cross-section | P, Ha, B | BMI; Smoking; Income |
| 96 | European Social Survey (ESS) | Greece | 2014 | 2755 | 15–90 | Repeat cross-section | P, Ha, B | BMI; Smoking; Income |
| 97 | European Social Survey (ESS) | Hungary | 2014 | 3780 | 15–92 | Repeat cross-section | P, Ha, B | BMI; Smoking; Income |

| | | | | | | | | |
|---|---|---|---|---|---|---|---|---|
| 98 | European Social Survey (ESS) | Iceland | 2014 | 839 | 16–90 | Repeat cross-section | P, Ha, B | BMI; Smoking; Income |
| 99 | European Social Survey (ESS) | Ireland | 2014 | 4376 | 15–97 | Repeat cross-section | P, Ha, B | BMI; Smoking; Income |
| 100 | European Social Survey (ESS) | Israel | 2014 | 3422 | 15–100 | Repeat cross-section | P, Ha, B | BMI; Smoking; Income |
| 101 | European Social Survey (ESS) | Italy | 2014 | 2835 | 15–90 | Repeat cross-section | P, Ha, B | BMI; Smoking; Income |
| 102 | European Social Survey (ESS) | Latvia | 2014 | 1212 | 15–90 | Repeat cross-section | P, Ha, B | BMI; Smoking; Income |
| 103 | European Social Survey (ESS) | Lithuania | 2014 | 3597 | 15–92 | Repeat cross-section | P, Ha, B | BMI; Smoking; Income |
| 104 | European Social Survey (ESS) | Montenegro | 2014 | 1587 | 16–87 | Repeat cross-section | P, Ha, B | BMI; Smoking; Income |
| 105 | European Social Survey (ESS) | Netherlands | 2014 | 3606 | 14–95 | Repeat cross-section | P, Ha, B | BMI; Smoking; Income |
| 106 | European Social Survey (ESS) | Norway | 2014 | 2771 | 15–100 | Repeat cross-section | P, Ha, B | BMI; Smoking; Income |
| 107 | European Social Survey (ESS) | Poland | 2014 | 3029 | 15–90 | Repeat cross-section | P, Ha, B | BMI; Smoking; Income |
| 108 | European Social Survey (ESS) | Portugal | 2014 | 2638 | 15–94 | Repeat cross-section | P, Ha, B | BMI; Smoking; Income |
| 109 | European Social Survey (ESS) | Serbia | 2014 | 1543 | 15–90 | Repeat cross-section | P, Ha, B | BMI; Smoking; Income |
| 110 | European Social Survey (ESS) | Slovakia | 2014 | 1431 | 15–90 | Repeat cross-section | P, Ha, B | BMI; Smoking; Income |
| 111 | European Social Survey (ESS) | Slovenia | 2014 | 2472 | 15–94 | Repeat cross-section | P, Ha, B | BMI; Smoking; Income |
| 112 | European Social Survey (ESS) | Spain | 2014 | 3760 | 15–100 | Repeat cross-section | P, Ha, B | BMI; Smoking; Income |
| 113 | European Social Survey (ESS) | Sweden | 2014 | 3019 | 15–97 | Repeat cross-section | P, Ha, B | BMI; Smoking; Income |
| 114 | European Social Survey (ESS) | Switzerland | 2014 | 2907 | 15–92 | Repeat cross-section | P, Ha, B | BMI; Smoking; Income |
| 115 | European Social Survey (ESS) | United Kingdom | 2014 | 3890 | 15–94 | Repeat cross-section | P, Ha, B | BMI; Smoking; Income |
| 116 | European Working Conditions Survey (EWCS) | Albania | 2010 | 992 | 16–86 | Repeat cross-section | P, Ha, N/S, S/A, B | Income |

| | | | | | | | | |
|---|---|---|---|---|---|---|---|---|
| 117 | European Working Conditions Survey (EWCS) | Albania | 2015 | 1002 | 16–75 | Repeat cross-section | P, Ha, N/S, B | Income |
| 118 | European Working Conditions Survey (EWCS) | Albania | 2021 | 501 | 17–73 | Repeat cross-section | P, Ha, N/S, B | Income |
| 119 | European Working Conditions Survey (EWCS) | Austria | 1995 | 588 | 16–70 | Repeat cross-section | P, Ha, N/S, S/A, B | |
| 120 | European Working Conditions Survey (EWCS) | Austria | 2000 | 704 | 17–72 | Repeat cross-section | P, Ha, N/S, S/A, B | |
| 121 | European Working Conditions Survey (EWCS) | Austria | 2005 | 282 | 16–74 | Repeat cross-section | P, Ha, N/S, S/A, B | |
| 122 | European Working Conditions Survey (EWCS) | Austria | 2010 | 1003 | 15–89 | Repeat cross-section | P, Ha, N/S, S/A, B | Income |
| 123 | European Working Conditions Survey (EWCS) | Austria | 2015 | 1024 | 15–87 | Repeat cross-section | P, Ha, N/S, B | Income |
| 124 | European Working Conditions Survey (EWCS) | Austria | 2021 | 889 | 16–75 | Repeat cross-section | P, Ha, N/S, B | Income |
| 125 | European Working Conditions Survey (EWCS) | Belgium | 1991 | 344 | 17–67 | Repeat cross-section | P, N/S, B | |
| 126 | European Working Conditions Survey (EWCS) | Belgium | 1995 | 398 | 19–67 | Repeat cross-section | P, Ha, N/S, S/A, B | |
| 127 | European Working Conditions Survey (EWCS) | Belgium | 2000 | 788 | 16–88 | Repeat cross-section | P, Ha, N/S, S/A, B | |
| 128 | European Working Conditions Survey (EWCS) | Belgium | 2005 | 287 | 18–66 | Repeat cross-section | P, Ha, N/S, S/A, B | |
| 129 | European Working Conditions Survey (EWCS) | Belgium | 2010 | 3954 | 16–84 | Repeat cross-section | P, Ha, N/S, S/A, B | Income |
| 130 | European Working Conditions Survey (EWCS) | Belgium | 2015 | 2583 | 15–88 | Repeat cross-section | P, Ha, N/S, B | Income |
| 131 | European Working Conditions Survey (EWCS) | Belgium | 2021 | 2099 | 16–83 | Repeat cross-section | P, Ha, N/S, B | Income |
| 132 | European Working Conditions Survey (EWCS) | Bosnia and Herzegovina | 2021 | 568 | 16–73 | Repeat cross-section | P, Ha, N/S, B | Income |
| 133 | European Working Conditions Survey (EWCS) | Bulgaria | 2001 | 689 | 19–64 | Repeat cross-section | P, Ha, N/S, S/A, B | |
| 134 | European Working Conditions Survey (EWCS) | Bulgaria | 2005 | 521 | 17–75 | Repeat cross-section | P, Ha, N/S, S/A, B | |
| 135 | European Working Conditions Survey (EWCS) | Bulgaria | 2010 | 1012 | 18–77 | Repeat cross-section | P, Ha, N/S, S/A, B | Income |
| 136 | European Working Conditions Survey (EWCS) | Bulgaria | 2015 | 1064 | 18–78 | Repeat cross-section | P, Ha, N/S, B | Income |
| 137 | European Working Conditions Survey (EWCS) | Bulgaria | 2021 | 885 | 16–80 | Repeat cross-section | P, Ha, N/S, B | Income |
| 138 | European Working Conditions Survey (EWCS) | Croatia | 2005 | 479 | 18–76 | Repeat cross-section | P, Ha, N/S, S/A, B | |
| 139 | European Working Conditions Survey (EWCS) | Croatia | 2010 | 1089 | 17–80 | Repeat cross-section | P, Ha, N/S, S/A, B | Income |
| 140 | European Working Conditions Survey (EWCS) | Croatia | 2015 | 1006 | 17–84 | Repeat cross-section | P, Ha, N/S, B | Income |
| 141 | European Working Conditions Survey (EWCS) | Croatia | 2021 | 894 | 18–74 | Repeat cross-section | P, Ha, N/S, B | Income |
| 142 | European Working Conditions Survey (EWCS) | Cyprus | 2001 | 297 | 16–71 | Repeat cross-section | P, Ha, N/S, S/A, B | |
| 143 | European Working Conditions Survey (EWCS) | Cyprus | 2005 | 259 | 19–76 | Repeat cross-section | P, Ha, N/S, S/A, B | |
| 144 | European Working Conditions Survey (EWCS) | Cyprus | 2010 | 997 | 17–84 | Repeat cross-section | P, Ha, N/S, S/A, B | Income |
| 145 | European Working Conditions Survey (EWCS) | Cyprus | 2015 | 1001 | 16–82 | Repeat cross-section | P, Ha, N/S, B | Income |
| 146 | European Working Conditions Survey (EWCS) | Cyprus | 2021 | 681 | 18–75 | Repeat cross-section | P, Ha, N/S, B | Income |
| 147 | European Working Conditions Survey (EWCS) | Czech Republic | 2001 | 720 | 18–69 | Repeat cross-section | P, Ha, N/S, S/A, B | |
| 148 | European Working Conditions Survey (EWCS) | Czech Republic | 2005 | 324 | 18–99 | Repeat cross-section | P, Ha, N/S, S/A, B | |
| 149 | European Working Conditions Survey (EWCS) | Czech Republic | 2010 | 992 | 18–85 | Repeat cross-section | P, Ha, N/S, S/A, B | Income |

| | | | | | | | | |
|---|---|---|---|---|---|---|---|---|
| 150 | European Working Conditions Survey (EWCS) | Czech Republic | 2015 | 997 | 18–81 | Repeat cross-section | P, Ha, N/S, B | Income |
| 151 | European Working Conditions Survey (EWCS) | Czech Republic | 2021 | 997 | 17–87 | Repeat cross-section | P, Ha, N/S, B | Income |
| 152 | European Working Conditions Survey (EWCS) | Denmark | 1991 | 589 | 16–75 | Repeat cross-section | P, N/S, B | |
| 153 | European Working Conditions Survey (EWCS) | Denmark | 1995 | 598 | 15–69 | Repeat cross-section | P, Ha, N/S, S/A, B | |
| 154 | European Working Conditions Survey (EWCS) | Denmark | 2000 | 936 | 16–70 | Repeat cross-section | P, Ha, N/S, S/A, B | |
| 155 | European Working Conditions Survey (EWCS) | Denmark | 2005 | 451 | 15–74 | Repeat cross-section | P, Ha, N/S, S/A, B | |
| 156 | European Working Conditions Survey (EWCS) | Denmark | 2010 | 1068 | 15–68 | Repeat cross-section | P, Ha, N/S, S/A, B | Income |
| 157 | European Working Conditions Survey (EWCS) | Denmark | 2015 | 998 | 15–76 | Repeat cross-section | P, Ha, N/S, B | Income |
| 158 | European Working Conditions Survey (EWCS) | Denmark | 2021 | 894 | 16–75 | Repeat cross-section | P, Ha, N/S, B | Income |
| 159 | European Working Conditions Survey (EWCS) | Estonia | 2001 | 771 | 18–74 | Repeat cross-section | P, Ha, N/S, S/A, B | |
| 160 | European Working Conditions Survey (EWCS) | Estonia | 2005 | 349 | 18–74 | Repeat cross-section | P, Ha, N/S, S/A, B | |
| 161 | European Working Conditions Survey (EWCS) | Estonia | 2010 | 1000 | 18–81 | Repeat cross-section | P, Ha, N/S, S/A, B | Income |
| 162 | European Working Conditions Survey (EWCS) | Estonia | 2015 | 997 | 15–86 | Repeat cross-section | P, Ha, N/S, B | Income |
| 163 | European Working Conditions Survey (EWCS) | Estonia | 2021 | 901 | 18–77 | Repeat cross-section | P, Ha, N/S, B | Income |
| 164 | European Working Conditions Survey (EWCS) | Finland | 1995 | 683 | 17–70 | Repeat cross-section | P, Ha, N/S, S/A, B | |
| 165 | European Working Conditions Survey (EWCS) | Finland | 2000 | 1190 | 15–98 | Repeat cross-section | P, Ha, N/S, S/A, B | |
| 166 | European Working Conditions Survey (EWCS) | Finland | 2005 | 445 | 16–64 | Repeat cross-section | P, Ha, N/S, S/A, B | |
| 167 | European Working Conditions Survey (EWCS) | Finland | 2010 | 1027 | 15–74 | Repeat cross-section | P, Ha, N/S, S/A, B | Income |
| 168 | European Working Conditions Survey (EWCS) | Finland | 2015 | 999 | 18–83 | Repeat cross-section | P, Ha, N/S, B | Income |
| 169 | European Working Conditions Survey (EWCS) | Finland | 2021 | 953 | 18–80 | Repeat cross-section | P, Ha, N/S, B | Income |
| 170 | European Working Conditions Survey (EWCS) | France | 1991 | 437 | 16–63 | Repeat cross-section | P, N/S, B | |
| 171 | European Working Conditions Survey (EWCS) | France | 1995 | 521 | 18–63 | Repeat cross-section | P, Ha, N/S, S/A, B | |
| 172 | European Working Conditions Survey (EWCS) | France | 2000 | 953 | 16–68 | Repeat cross-section | P, Ha, N/S, S/A, B | |
| 173 | European Working Conditions Survey (EWCS) | France | 2005 | 298 | 17–60 | Repeat cross-section | P, Ha, N/S, S/A, B | |
| 174 | European Working Conditions Survey (EWCS) | France | 2010 | 3042 | 15–75 | Repeat cross-section | P, Ha, N/S, S/A, B | Income |
| 175 | European Working Conditions Survey (EWCS) | France | 2015 | 1525 | 16–75 | Repeat cross-section | P, Ha, N/S, B | Income |
| 176 | European Working Conditions Survey (EWCS) | France | 2021 | 1603 | 17–74 | Repeat cross-section | P, Ha, N/S, B | Income |
| 177 | European Working Conditions Survey (EWCS) | Germany | 1991 | 1050 | 15–75 | Repeat cross-section | P, N/S, B | |
| 178 | European Working Conditions Survey (EWCS) | Germany | 1995 | 1200 | 16–72 | Repeat cross-section | P, Ha, N/S, S/A, B | |
| 179 | European Working Conditions Survey (EWCS) | Germany | 2000 | 920 | 15–75 | Repeat cross-section | P, Ha, N/S, S/A, B | |
| 180 | European Working Conditions Survey (EWCS) | Germany | 2005 | 231 | 17–65 | Repeat cross-section | P, Ha, N/S, S/A, B | |
| 181 | European Working Conditions Survey (EWCS) | Germany | 2010 | 2130 | 15–77 | Repeat cross-section | P, Ha, N/S, S/A, B | Income |
| 182 | European Working Conditions Survey (EWCS) | Germany | 2015 | 2087 | 15–79 | Repeat cross-section | P, Ha, N/S, B | Income |

| | | | | | | | | |
|---|---|---|---|---|---|---|---|---|
| 183 | European Working Conditions Survey (EWCS) | Germany | 2021 | 2048 | 16–80 | Repeat cross-section | P, Ha, N/S, B | Income |
| 184 | European Working Conditions Survey (EWCS) | Greece | 1991 | 484 | 16–76 | Repeat cross-section | P, N/S, B | |
| 185 | European Working Conditions Survey (EWCS) | Greece | 1995 | 782 | 17–75 | Repeat cross-section | P, Ha, N/S, S/A, B | |
| 186 | European Working Conditions Survey (EWCS) | Greece | 2000 | 1267 | 17–78 | Repeat cross-section | P, Ha, N/S, S/A, B | |
| 187 | European Working Conditions Survey (EWCS) | Greece | 2005 | 707 | 16–85 | Repeat cross-section | P, Ha, N/S, S/A, B | |
| 188 | European Working Conditions Survey (EWCS) | Greece | 2010 | 1036 | 16–80 | Repeat cross-section | P, Ha, N/S, S/A, B | Income |
| 189 | European Working Conditions Survey (EWCS) | Greece | 2015 | 1007 | 18–85 | Repeat cross-section | P, Ha, N/S, B | Income |
| 190 | European Working Conditions Survey (EWCS) | Greece | 2021 | 899 | 17–74 | Repeat cross-section | P, Ha, N/S, B | Income |
| 191 | European Working Conditions Survey (EWCS) | Hungary | 2001 | 612 | 17–79 | Repeat cross-section | P, Ha, N/S, S/A, B | |
| 192 | European Working Conditions Survey (EWCS) | Hungary | 2005 | 458 | 20–74 | Repeat cross-section | P, Ha, N/S, S/A, B | |
| 193 | European Working Conditions Survey (EWCS) | Hungary | 2010 | 1006 | 16–77 | Repeat cross-section | P, Ha, N/S, S/A, B | Income |
| 194 | European Working Conditions Survey (EWCS) | Hungary | 2015 | 1011 | 18–89 | Repeat cross-section | P, Ha, N/S, B | Income |
| 195 | European Working Conditions Survey (EWCS) | Hungary | 2021 | 893 | 18–77 | Repeat cross-section | P, Ha, N/S, B | Income |
| 196 | European Working Conditions Survey (EWCS) | Ireland | 1991 | 281 | 16–72 | Repeat cross-section | P, N/S, B | |
| 197 | European Working Conditions Survey (EWCS) | Ireland | 1995 | 291 | 15–71 | Repeat cross-section | P, Ha, N/S, S/A, B | |
| 198 | European Working Conditions Survey (EWCS) | Ireland | 2000 | 445 | 16–78 | Repeat cross-section | P, Ha, N/S, S/A, B | |
| 199 | European Working Conditions Survey (EWCS) | Ireland | 2005 | 270 | 16–75 | Repeat cross-section | P, Ha, N/S, S/A, B | |
| 200 | European Working Conditions Survey (EWCS) | Ireland | 2010 | 994 | 16–79 | Repeat cross-section | P, Ha, N/S, S/A, B | Income |
| 201 | European Working Conditions Survey (EWCS) | Ireland | 2015 | 1043 | 15–87 | Repeat cross-section | P, Ha, N/S, B | Income |
| 202 | European Working Conditions Survey (EWCS) | Ireland | 2021 | 879 | 16–77 | Repeat cross-section | P, Ha, N/S, B | Income |
| 203 | European Working Conditions Survey (EWCS) | Italy | 1991 | 315 | 15–64 | Repeat cross-section | P, N/S, B | |
| 204 | European Working Conditions Survey (EWCS) | Italy | 1995 | 686 | 15–66 | Repeat cross-section | P, Ha, N/S, S/A, B | |
| 205 | European Working Conditions Survey (EWCS) | Italy | 2000 | 936 | 16–68 | Repeat cross-section | P, Ha, N/S, S/A, B | |
| 206 | European Working Conditions Survey (EWCS) | Italy | 2005 | 369 | 19–80 | Repeat cross-section | P, Ha, N/S, S/A, B | |
| 207 | European Working Conditions Survey (EWCS) | Italy | 2010 | 1466 | 16–76 | Repeat cross-section | P, Ha, N/S, S/A, B | Income |
| 208 | European Working Conditions Survey (EWCS) | Italy | 2015 | 1398 | 18–86 | Repeat cross-section | P, Ha, N/S, B | Income |
| 209 | European Working Conditions Survey (EWCS) | Italy | 2021 | 1563 | 16–84 | Repeat cross-section | P, Ha, N/S, B | Income |
| 210 | European Working Conditions Survey (EWCS) | Kosovo | 2010 | 1013 | 15–79 | Repeat cross-section | P, Ha, N/S, S/A, B | Income |
| 211 | European Working Conditions Survey (EWCS) | Kosovo | 2021 | 573 | 17–64 | Repeat cross-section | P, Ha, N/S, B | Income |
| 212 | European Working Conditions Survey (EWCS) | Latvia | 2001 | 795 | 16–80 | Repeat cross-section | P, Ha, N/S, S/A, B | |
| 213 | European Working Conditions Survey (EWCS) | Latvia | 2005 | 623 | 17–74 | Repeat cross-section | P, Ha, N/S, S/A, B | |
| 214 | European Working Conditions Survey (EWCS) | Latvia | 2010 | 1000 | 18–80 | Repeat cross-section | P, Ha, N/S, S/A, B | Income |
| 215 | European Working Conditions Survey (EWCS) | Latvia | 2015 | 989 | 17–86 | Repeat cross-section | P, Ha, N/S, B | Income |

| | | | | | | | | |
|---|---|---|---|---|---|---|---|---|
| 216 | European Working Conditions Survey (EWCS) | Latvia | 2021 | 893 | 17–77 | Repeat cross-section | P, Ha, N/S, B | Income |
| 217 | European Working Conditions Survey (EWCS) | Lithuania | 2001 | 741 | 19–79 | Repeat cross-section | P, Ha, N/S, S/A, B | |
| 218 | European Working Conditions Survey (EWCS) | Lithuania | 2005 | 481 | 19–75 | Repeat cross-section | P, Ha, N/S, S/A, B | |
| 219 | European Working Conditions Survey (EWCS) | Lithuania | 2010 | 1003 | 16–80 | Repeat cross-section | P, Ha, N/S, S/A, B | Income |
| 220 | European Working Conditions Survey (EWCS) | Lithuania | 2015 | 1003 | 17–86 | Repeat cross-section | P, Ha, N/S, B | Income |
| 221 | European Working Conditions Survey (EWCS) | Lithuania | 2021 | 933 | 19–77 | Repeat cross-section | P, Ha, N/S, B | Income |
| 222 | European Working Conditions Survey (EWCS) | Luxembourg | 1991 | 237 | 18–71 | Repeat cross-section | P, N/S, B | |
| 223 | European Working Conditions Survey (EWCS) | Luxembourg | 1995 | 260 | 17–70 | Repeat cross-section | P, Ha, N/S, S/A, B | |
| 224 | European Working Conditions Survey (EWCS) | Luxembourg | 2000 | 305 | 16–80 | Repeat cross-section | P, Ha, N/S, S/A, B | |
| 225 | European Working Conditions Survey (EWCS) | Luxembourg | 2005 | 220 | 18–67 | Repeat cross-section | P, Ha, N/S, S/A, B | |
| 226 | European Working Conditions Survey (EWCS) | Luxembourg | 2010 | 985 | 16–81 | Repeat cross-section | P, Ha, N/S, S/A, B | Income |
| 227 | European Working Conditions Survey (EWCS) | Luxembourg | 2015 | 1002 | 17–79 | Repeat cross-section | P, Ha, N/S, B | Income |
| 228 | European Working Conditions Survey (EWCS) | Luxembourg | 2021 | 672 | 16–74 | Repeat cross-section | P, Ha, N/S, B | Income |
| 229 | European Working Conditions Survey (EWCS) | Malta | 2001 | 339 | 16–79 | Repeat cross-section | P, Ha, N/S, S/A, B | |
| 230 | European Working Conditions Survey (EWCS) | Malta | 2005 | 298 | 16–74 | Repeat cross-section | P, Ha, N/S, S/A, B | |
| 231 | European Working Conditions Survey (EWCS) | Malta | 2010 | 1000 | 16–78 | Repeat cross-section | P, Ha, N/S, S/A, B | Income |
| 232 | European Working Conditions Survey (EWCS) | Malta | 2015 | 1001 | 16–75 | Repeat cross-section | P, Ha, N/S, B | Income |
| 233 | European Working Conditions Survey (EWCS) | Malta | 2021 | 732 | 16–76 | Repeat cross-section | P, Ha, N/S, B | Income |
| 234 | European Working Conditions Survey (EWCS) | Montenegro | 2010 | 1037 | 18–74 | Repeat cross-section | P, Ha, N/S, S/A, B | Income |
| 235 | European Working Conditions Survey (EWCS) | Montenegro | 2015 | 1005 | 16–83 | Repeat cross-section | P, Ha, N/S, B | Income |
| 236 | European Working Conditions Survey (EWCS) | Montenegro | 2021 | 573 | 17–70 | Repeat cross-section | P, Ha, N/S, B | Income |
| 237 | European Working Conditions Survey (EWCS) | Netherlands | 1991 | 367 | 17–60 | Repeat cross-section | P, N/S, B | |
| 238 | European Working Conditions Survey (EWCS) | Netherlands | 1995 | 478 | 17–63 | Repeat cross-section | P, Ha, N/S, S/A, B | |
| 239 | European Working Conditions Survey (EWCS) | Netherlands | 2000 | 896 | 15–64 | Repeat cross-section | P, Ha, N/S, S/A, B | |
| 240 | European Working Conditions Survey (EWCS) | Netherlands | 2005 | 275 | 17–64 | Repeat cross-section | P, Ha, N/S, S/A, B | |
| 241 | European Working Conditions Survey (EWCS) | Netherlands | 2010 | 1017 | 15–78 | Repeat cross-section | P, Ha, N/S, S/A, B | Income |
| 242 | European Working Conditions Survey (EWCS) | Netherlands | 2015 | 1023 | 15–81 | Repeat cross-section | P, Ha, N/S, B | Income |
| 243 | European Working Conditions Survey (EWCS) | Netherlands | 2021 | 898 | 16–75 | Repeat cross-section | P, Ha, N/S, B | Income |
| 244 | European Working Conditions Survey (EWCS) | North Macedonia | 2010 | 1099 | 17–82 | Repeat cross-section | P, Ha, N/S, S/A, B | Income |
| 245 | European Working Conditions Survey (EWCS) | North Macedonia | 2015 | 1005 | 15–82 | Repeat cross-section | P, Ha, N/S, B | Income |
| 246 | European Working Conditions Survey (EWCS) | North Macedonia | 2021 | 551 | 17–71 | Repeat cross-section | P, Ha, N/S, B | Income |
| 247 | European Working Conditions Survey (EWCS) | Norway | 2005 | 453 | 16–72 | Repeat cross-section | P, Ha, N/S, S/A, B | |
| 248 | European Working Conditions Survey (EWCS) | Norway | 2010 | 1085 | 16–89 | Repeat cross-section | P, Ha, N/S, S/A, B | Income |

| | | | | | | | | |
|---|---|---|---|---|---|---|---|---|
| 249 | European Working Conditions Survey (EWCS) | Norway | 2015 | 1028 | 15–73 | Repeat cross-section | P, Ha, N/S, B | Income |
| 250 | European Working Conditions Survey (EWCS) | Norway | 2021 | 1644 | 16–82 | Repeat cross-section | P, Ha, N/S, B | Income |
| 251 | European Working Conditions Survey (EWCS) | Poland | 2001 | 724 | 17–72 | Repeat cross-section | P, Ha, N/S, S/A, B | |
| 252 | European Working Conditions Survey (EWCS) | Poland | 2005 | 607 | 16–74 | Repeat cross-section | P, Ha, N/S, S/A, B | |
| 253 | European Working Conditions Survey (EWCS) | Poland | 2010 | 1471 | 16–80 | Repeat cross-section | P, Ha, N/S, S/A, B | Income |
| 254 | European Working Conditions Survey (EWCS) | Poland | 2015 | 1142 | 16–85 | Repeat cross-section | P, Ha, N/S, B | Income |
| 255 | European Working Conditions Survey (EWCS) | Poland | 2021 | 1439 | 16–88 | Repeat cross-section | P, Ha, N/S, B | Income |
| 256 | European Working Conditions Survey (EWCS) | Portugal | 1991 | 320 | 15–78 | Repeat cross-section | P, N/S, B | |
| 257 | European Working Conditions Survey (EWCS) | Portugal | 1995 | 696 | 15–76 | Repeat cross-section | P, Ha, N/S, S/A, B | |
| 258 | European Working Conditions Survey (EWCS) | Portugal | 2000 | 880 | 16–76 | Repeat cross-section | P, Ha, N/S, S/A, B | |
| 259 | European Working Conditions Survey (EWCS) | Portugal | 2005 | 380 | 18–75 | Repeat cross-section | P, Ha, N/S, S/A, B | |
| 260 | European Working Conditions Survey (EWCS) | Portugal | 2010 | 999 | 18–87 | Repeat cross-section | P, Ha, N/S, S/A, B | Income |
| 261 | European Working Conditions Survey (EWCS) | Portugal | 2015 | 1034 | 18–87 | Repeat cross-section | P, Ha, N/S, B | Income |
| 262 | European Working Conditions Survey (EWCS) | Portugal | 2021 | 943 | 18–75 | Repeat cross-section | P, Ha, N/S, B | Income |
| 263 | European Working Conditions Survey (EWCS) | Romania | 2001 | 619 | 17–81 | Repeat cross-section | P, Ha, N/S, S/A, B | |
| 264 | European Working Conditions Survey (EWCS) | Romania | 2005 | 499 | 16–89 | Repeat cross-section | P, Ha, N/S, S/A, B | |
| 265 | European Working Conditions Survey (EWCS) | Romania | 2010 | 1010 | 17–84 | Repeat cross-section | P, Ha, N/S, S/A, B | Income |
| 266 | European Working Conditions Survey (EWCS) | Romania | 2015 | 1061 | 18–83 | Repeat cross-section | P, Ha, N/S, B | Income |
| 267 | European Working Conditions Survey (EWCS) | Romania | 2021 | 908 | 18–74 | Repeat cross-section | P, Ha, N/S, B | Income |
| 268 | European Working Conditions Survey (EWCS) | Serbia | 2015 | 1027 | 15–85 | Repeat cross-section | P, Ha, N/S, B | Income |
| 269 | European Working Conditions Survey (EWCS) | Serbia | 2021 | 571 | 17–68 | Repeat cross-section | P, Ha, N/S, B | Income |
| 270 | European Working Conditions Survey (EWCS) | Slovakia | 2001 | 727 | 16–73 | Repeat cross-section | P, Ha, N/S, S/A, B | |
| 271 | European Working Conditions Survey (EWCS) | Slovakia | 2005 | 522 | 20–65 | Repeat cross-section | P, Ha, N/S, S/A, B | |
| 272 | European Working Conditions Survey (EWCS) | Slovakia | 2010 | 1001 | 15–82 | Repeat cross-section | P, Ha, N/S, S/A, B | Income |
| 273 | European Working Conditions Survey (EWCS) | Slovakia | 2015 | 989 | 16–81 | Repeat cross-section | P, Ha, N/S, B | Income |
| 274 | European Working Conditions Survey (EWCS) | Slovakia | 2021 | 894 | 18–80 | Repeat cross-section | P, Ha, N/S, B | Income |
| 275 | European Working Conditions Survey (EWCS) | Slovenia | 2001 | 635 | 18–76 | Repeat cross-section | P, Ha, N/S, S/A, B | |
| 276 | European Working Conditions Survey (EWCS) | Slovenia | 2005 | 361 | 17–76 | Repeat cross-section | P, Ha, N/S, S/A, B | |
| 277 | European Working Conditions Survey (EWCS) | Slovenia | 2010 | 1397 | 16–81 | Repeat cross-section | P, Ha, N/S, S/A, B | Income |
| 278 | European Working Conditions Survey (EWCS) | Slovenia | 2015 | 1599 | 17–86 | Repeat cross-section | P, Ha, N/S, B | Income |
| 279 | European Working Conditions Survey (EWCS) | Slovenia | 2021 | 1308 | 16–70 | Repeat cross-section | P, Ha, N/S, B | Income |
| 280 | European Working Conditions Survey (EWCS) | Spain | 1991 | 347 | 17–89 | Repeat cross-section | P, N/S, B | |
| 281 | European Working Conditions Survey (EWCS) | Spain | 1995 | 567 | 16–67 | Repeat cross-section | P, Ha, N/S, S/A, B | |

| | | | | | | | | |
|---|---|---|---|---|---|---|---|---|
| 282 | European Working Conditions Survey (EWCS) | Spain | 2000 | 987 | 17–75 | Repeat cross-section | P, Ha, N/S, S/A, B | |
| 283 | European Working Conditions Survey (EWCS) | Spain | 2005 | 328 | 17–70 | Repeat cross-section | P, Ha, N/S, S/A, B | |
| 284 | European Working Conditions Survey (EWCS) | Spain | 2010 | 1007 | 17–70 | Repeat cross-section | P, Ha, N/S, S/A, B | Income |
| 285 | European Working Conditions Survey (EWCS) | Spain | 2015 | 3357 | 15–87 | Repeat cross-section | P, Ha, N/S, B | Income |
| 286 | European Working Conditions Survey (EWCS) | Spain | 2021 | 1445 | 18–88 | Repeat cross-section | P, Ha, N/S, B | Income |
| 287 | European Working Conditions Survey (EWCS) | Sweden | 1995 | 741 | 18–70 | Repeat cross-section | P, Ha, N/S, S/A, B | |
| 288 | European Working Conditions Survey (EWCS) | Sweden | 2000 | 1094 | 16–79 | Repeat cross-section | P, Ha, N/S, S/A, B | |
| 289 | European Working Conditions Survey (EWCS) | Sweden | 2005 | 567 | 16–66 | Repeat cross-section | P, Ha, N/S, S/A, B | |
| 290 | European Working Conditions Survey (EWCS) | Sweden | 2010 | 972 | 17–77 | Repeat cross-section | P, Ha, N/S, S/A, B | Income |
| 291 | European Working Conditions Survey (EWCS) | Sweden | 2015 | 1002 | 17–75 | Repeat cross-section | P, Ha, N/S, B | Income |
| 292 | European Working Conditions Survey (EWCS) | Sweden | 2021 | 909 | 16–84 | Repeat cross-section | P, Ha, N/S, B | Income |
| 293 | European Working Conditions Survey (EWCS) | Switzerland | 2005 | 329 | 19–86 | Repeat cross-section | P, Ha, N/S, S/A, B | |
| 294 | European Working Conditions Survey (EWCS) | Switzerland | 2015 | 1003 | 16–88 | Repeat cross-section | P, Ha, N/S, B | Income |
| 295 | European Working Conditions Survey (EWCS) | Switzerland | 2021 | 619 | 16–79 | Repeat cross-section | P, Ha, N/S, B | Income |
| 296 | European Working Conditions Survey (EWCS) | Turkiye | 2005 | 464 | 16–75 | Repeat cross-section | P, Ha, N/S, S/A, B | |
| 297 | European Working Conditions Survey (EWCS) | Turkiye | 2010 | 2099 | 16–80 | Repeat cross-section | P, Ha, N/S, S/A, B | Income |
| 298 | European Working Conditions Survey (EWCS) | Turkiye | 2015 | 1994 | 15–84 | Repeat cross-section | P, Ha, N/S, B | Income |
| 299 | European Working Conditions Survey (EWCS) | United Kingdom | 1991 | 526 | 17–72 | Repeat cross-section | P, N/S, B | |
| 300 | European Working Conditions Survey (EWCS) | United Kingdom | 1995 | 524 | 15–65 | Repeat cross-section | P, Ha, N/S, S/A, B | |
| 301 | European Working Conditions Survey (EWCS) | United Kingdom | 2000 | 711 | 16–72 | Repeat cross-section | P, Ha, N/S, S/A, B | |
| 302 | European Working Conditions Survey (EWCS) | United Kingdom | 2005 | 212 | 17–70 | Repeat cross-section | P, Ha, N/S, S/A, B | |
| 303 | European Working Conditions Survey (EWCS) | United Kingdom | 2010 | 1564 | 16–91 | Repeat cross-section | P, Ha, N/S, S/A, B | Income |
| 304 | European Working Conditions Survey (EWCS) | United Kingdom | 2015 | 1618 | 16–84 | Repeat cross-section | P, Ha, N/S, B | Income |
| 305 | European Working Conditions Survey (EWCS) | United Kingdom | 2021 | 1047 | 18–81 | Repeat cross-section | P, Ha, N/S, B | Income |
| 306 | Gender and Adolescence: Global Evidence | Bangladesh | 2017 | 780 | 10–17 | Household survey | Ha, S/A | |
| 307 | Gender and Adolescence: Global Evidence | Ethiopia | 2017 | 6586 | 10–17 | Household survey | Ha, S/A | |
| 308 | Gender and Adolescence: Global Evidence | Jordan | 2018 | 4101 | 10–18 | Household survey | Ha, S/A | |
| 309 | Generation 21 (G21)[a,f] | Portugal | 2012 | 5814 | 7–7 | Cohort | P, Ha, C, S/A, B | |
| 310 | German Health Interview and Examination Survey for Children and Adolescents (KiGGS)[b] | Germany | 2003 | 14560 | 5–16 | Panel | P, Ha, C, S/A, B, F | BMI; Income |
| 311 | German National Health Interview and Examination Survey 1998 (GNHIES98) | Germany | 1998 | 6968 | 20–77 | Cross-section | P, Ha, N/S, C, S/A, B, K, H, F, Hnd, Ft | BMI; Smoking; Income |
| 312 | Health Behavior in School-aged Children (HBSC) | Albania | 2014 | 4845 | 10–16 | School-based survey | Ha, S/A, B | BMI |
| 313 | Health Behavior in School-aged Children (HBSC) | Albania | 2018 | 1647 | 11–16 | School-based survey | Ha, S/A, B | BMI |

| | | | | | | | | |
|---|---|---|---|---|---|---|---|---|
| 314 | Health Behavior in School-aged Children (HBSC) | Armenia | 2010 | 2394 | 10–16 | School-based survey | Ha, S/A, B | BMI |
| 315 | Health Behavior in School-aged Children (HBSC) | Armenia | 2014 | 3019 | 10–16 | School-based survey | Ha, S/A, B | BMI |
| 316 | Health Behavior in School-aged Children (HBSC) | Armenia | 2018 | 4123 | 11–16 | School-based survey | Ha, S/A, B | BMI |
| 317 | Health Behavior in School-aged Children (HBSC) | Austria | 2001 | 4294 | 10–16 | School-based survey | Ha, S/A, B | BMI |
| 318 | Health Behavior in School-aged Children (HBSC) | Austria | 2006 | 4718 | 10–16 | School-based survey | Ha, S/A, B | BMI |
| 319 | Health Behavior in School-aged Children (HBSC) | Austria | 2010 | 4929 | 10–16 | School-based survey | Ha, S/A, B | BMI |
| 320 | Health Behavior in School-aged Children (HBSC) | Austria | 2014 | 3358 | 11–16 | School-based survey | Ha, S/A, B | BMI |
| 321 | Health Behavior in School-aged Children (HBSC) | Austria | 2018 | 4044 | 10–16 | School-based survey | Ha, S/A, B | BMI |
| 322 | Health Behavior in School-aged Children (HBSC) | Azerbaijan | 2018 | 4397 | 10–16 | School-based survey | Ha, S/A, B | BMI |
| 323 | Health Behavior in School-aged Children (HBSC) | Belgium | 2001 | 10438 | 11–16 | School-based survey | Ha, S/A, B | BMI |
| 324 | Health Behavior in School-aged Children (HBSC) | Belgium | 2006 | 8417 | 11–16 | School-based survey | Ha, S/A, B | BMI |
| 325 | Health Behavior in School-aged Children (HBSC) | Belgium | 2010 | 7943 | 10–16 | School-based survey | Ha, S/A, B | BMI |
| 326 | Health Behavior in School-aged Children (HBSC) | Belgium | 2014 | 10050 | 10–16 | School-based survey | Ha, S/A, B | BMI |
| 327 | Health Behavior in School-aged Children (HBSC) | Belgium | 2018 | 9702 | 10–16 | School-based survey | Ha, S/A, B | BMI |
| 328 | Health Behavior in School-aged Children (HBSC) | Bulgaria | 2006 | 4799 | 11–16 | School-based survey | Ha, S/A, B | BMI |
| 329 | Health Behavior in School-aged Children (HBSC) | Bulgaria | 2014 | 4705 | 11–16 | School-based survey | Ha, S/A, B | BMI |
| 330 | Health Behavior in School-aged Children (HBSC) | Bulgaria | 2018 | 4548 | 11–16 | School-based survey | Ha, S/A, B | BMI |
| 331 | Health Behavior in School-aged Children (HBSC) | Canada | 2001 | 4321 | 11–16 | School-based survey | Ha, S/A, B | BMI |
| 332 | Health Behavior in School-aged Children (HBSC) | Canada | 2006 | 5720 | 11–16 | School-based survey | Ha, S/A, B | BMI |
| 333 | Health Behavior in School-aged Children (HBSC) | Canada | 2010 | 15295 | 11–16 | School-based survey | Ha, S/A, B | BMI |
| 334 | Health Behavior in School-aged Children (HBSC) | Canada | 2014 | 12549 | 11–16 | School-based survey | Ha, S/A, B | BMI |
| 335 | Health Behavior in School-aged Children (HBSC) | Canada | 2018 | 12243 | 11–16 | School-based survey | Ha, S/A, B | BMI |
| 336 | Health Behavior in School-aged Children (HBSC) | Croatia | 2001 | 4317 | 10–16 | School-based survey | Ha, S/A, B | BMI |
| 337 | Health Behavior in School-aged Children (HBSC) | Croatia | 2006 | 4854 | 11–16 | School-based survey | Ha, S/A, B | BMI |
| 338 | Health Behavior in School-aged Children (HBSC) | Croatia | 2010 | 6203 | 10–16 | School-based survey | Ha, S/A, B | BMI |
| 339 | Health Behavior in School-aged Children (HBSC) | Croatia | 2014 | 5335 | 11–16 | School-based survey | Ha, S/A, B | BMI |
| 340 | Health Behavior in School-aged Children (HBSC) | Croatia | 2018 | 4901 | 11–16 | School-based survey | Ha, S/A, B | BMI |
| 341 | Health Behavior in School-aged Children (HBSC) | Czech Republic | 2001 | 4954 | 11–16 | School-based survey | Ha, S/A, B | BMI |
| 342 | Health Behavior in School-aged Children (HBSC) | Czech Republic | 2006 | 4698 | 11–16 | School-based survey | Ha, S/A, B | BMI |
| 343 | Health Behavior in School-aged Children (HBSC) | Czech Republic | 2010 | 4305 | 11–16 | School-based survey | Ha, S/A, B | BMI |
| 344 | Health Behavior in School-aged Children (HBSC) | Czech Republic | 2014 | 5018 | 10–16 | School-based survey | Ha, S/A, B | BMI |
| 345 | Health Behavior in School-aged Children (HBSC) | Czech Republic | 2018 | 11010 | 10–16 | School-based survey | Ha, S/A, B | BMI |
| 346 | Health Behavior in School-aged Children (HBSC) | Denmark | 2001 | 4538 | 11–17 | School-based survey | Ha, S/A, B | BMI |

| | | | | | | | | |
|---|---|---|---|---|---|---|---|---|
| 347 | Health Behavior in School-aged Children (HBSC) | Denmark | 2006 | 5586 | 11–17 | School-based survey | Ha, S/A, B | BMI |
| 348 | Health Behavior in School-aged Children (HBSC) | Denmark | 2010 | 3987 | 11–16 | School-based survey | Ha, S/A, B | BMI |
| 349 | Health Behavior in School-aged Children (HBSC) | Denmark | 2014 | 3660 | 11–16 | School-based survey | Ha, S/A, B | BMI |
| 350 | Health Behavior in School-aged Children (HBSC) | Denmark | 2018 | 3118 | 11–16 | School-based survey | Ha, S/A, B | BMI |
| 351 | Health Behavior in School-aged Children (HBSC) | Estonia | 2001 | 3971 | 10–16 | School-based survey | Ha, S/A, B | BMI |
| 352 | Health Behavior in School-aged Children (HBSC) | Estonia | 2006 | 4407 | 11–16 | School-based survey | Ha, S/A, B | BMI |
| 353 | Health Behavior in School-aged Children (HBSC) | Estonia | 2010 | 4193 | 11–16 | School-based survey | Ha, S/A, B | BMI |
| 354 | Health Behavior in School-aged Children (HBSC) | Estonia | 2014 | 4004 | 11–16 | School-based survey | Ha, S/A, B | BMI |
| 355 | Health Behavior in School-aged Children (HBSC) | Estonia | 2018 | 4677 | 11–16 | School-based survey | Ha, S/A, B | BMI |
| 356 | Health Behavior in School-aged Children (HBSC) | Finland | 2001 | 5298 | 11–17 | School-based survey | Ha, S/A, B | BMI |
| 357 | Health Behavior in School-aged Children (HBSC) | Finland | 2006 | 5158 | 11–16 | School-based survey | Ha, S/A, B | BMI |
| 358 | Health Behavior in School-aged Children (HBSC) | Finland | 2010 | 6570 | 11–16 | School-based survey | Ha, S/A, B | BMI |
| 359 | Health Behavior in School-aged Children (HBSC) | Finland | 2014 | 5798 | 11–16 | School-based survey | Ha, S/A, B | BMI |
| 360 | Health Behavior in School-aged Children (HBSC) | Finland | 2018 | 3117 | 11–16 | School-based survey | Ha, S/A, B | BMI |
| 361 | Health Behavior in School-aged Children (HBSC) | France | 2001 | 7869 | 10–16 | School-based survey | Ha, S/A, B | BMI |
| 362 | Health Behavior in School-aged Children (HBSC) | France | 2006 | 7052 | 11–16 | School-based survey | Ha, S/A, B | BMI |
| 363 | Health Behavior in School-aged Children (HBSC) | France | 2010 | 6018 | 10–16 | School-based survey | Ha, S/A, B | BMI |
| 364 | Health Behavior in School-aged Children (HBSC) | France | 2014 | 5554 | 10–16 | School-based survey | Ha, S/A, B | BMI |
| 365 | Health Behavior in School-aged Children (HBSC) | France | 2018 | 8762 | 11–16 | School-based survey | Ha, S/A, B | BMI |
| 366 | Health Behavior in School-aged Children (HBSC) | Georgia | 2018 | 3864 | 11–16 | School-based survey | Ha, S/A, B | BMI |
| 367 | Health Behavior in School-aged Children (HBSC) | Germany | 2001 | 5485 | 11–17 | School-based survey | Ha, S/A, B | BMI |
| 368 | Health Behavior in School-aged Children (HBSC) | Germany | 2006 | 7133 | 10–16 | School-based survey | Ha, S/A, B | BMI |
| 369 | Health Behavior in School-aged Children (HBSC) | Germany | 2010 | 4915 | 10–16 | School-based survey | Ha, S/A, B | BMI |
| 370 | Health Behavior in School-aged Children (HBSC) | Germany | 2014 | 5846 | 10–16 | School-based survey | Ha, S/A, B | BMI |
| 371 | Health Behavior in School-aged Children (HBSC) | Germany | 2018 | 4286 | 10–16 | School-based survey | Ha, S/A, B | BMI |
| 372 | Health Behavior in School-aged Children (HBSC) | Greece | 2001 | 3718 | 11–16 | School-based survey | Ha, S/A, B | BMI |
| 373 | Health Behavior in School-aged Children (HBSC) | Greece | 2006 | 3655 | 11–16 | School-based survey | Ha, S/A, B | BMI |
| 374 | Health Behavior in School-aged Children (HBSC) | Greece | 2010 | 4835 | 11–16 | School-based survey | Ha, S/A, B | BMI |
| 375 | Health Behavior in School-aged Children (HBSC) | Greece | 2014 | 4074 | 11–16 | School-based survey | Ha, S/A, B | BMI |
| 376 | Health Behavior in School-aged Children (HBSC) | Greece | 2018 | 3799 | 11–16 | School-based survey | Ha, S/A, B | BMI |
| 377 | Health Behavior in School-aged Children (HBSC) | Greenland | 2001 | 817 | 11–17 | School-based survey | Ha, S/A, B | BMI |
| 378 | Health Behavior in School-aged Children (HBSC) | Greenland | 2006 | 1204 | 11–16 | School-based survey | Ha, S/A, B | BMI |
| 379 | Health Behavior in School-aged Children (HBSC) | Greenland | 2010 | 992 | 10–16 | School-based survey | Ha, S/A, B | BMI |

| | | | | | | | | |
|---|---|---|---|---|---|---|---|---|
| 380 | Health Behavior in School-aged Children (HBSC) | Greenland | 2014 | 882 | 10–16 | School-based survey | Ha, S/A, B | BMI |
| 381 | Health Behavior in School-aged Children (HBSC) | Greenland | 2018 | 1018 | 10–16 | School-based survey | Ha, S/A, B | BMI |
| 382 | Health Behavior in School-aged Children (HBSC) | Hungary | 2001 | 4036 | 11–16 | School-based survey | Ha, S/A, B | BMI |
| 383 | Health Behavior in School-aged Children (HBSC) | Hungary | 2006 | 3424 | 11–16 | School-based survey | Ha, S/A, B | BMI |
| 384 | Health Behavior in School-aged Children (HBSC) | Hungary | 2010 | 4742 | 11–16 | School-based survey | Ha, S/A, B | BMI |
| 385 | Health Behavior in School-aged Children (HBSC) | Hungary | 2014 | 3765 | 11–16 | School-based survey | Ha, S/A, B | BMI |
| 386 | Health Behavior in School-aged Children (HBSC) | Hungary | 2018 | 3682 | 11–16 | School-based survey | Ha, S/A, B | BMI |
| 387 | Health Behavior in School-aged Children (HBSC) | Iceland | 2006 | 9323 | 11–16 | School-based survey | Ha, S/A, B | BMI |
| 388 | Health Behavior in School-aged Children (HBSC) | Iceland | 2010 | 10877 | 10–16 | School-based survey | Ha, S/A, B | BMI |
| 389 | Health Behavior in School-aged Children (HBSC) | Iceland | 2014 | 10339 | 11–16 | School-based survey | Ha, S/A, B | BMI |
| 390 | Health Behavior in School-aged Children (HBSC) | Iceland | 2018 | 6842 | 11–16 | School-based survey | Ha, S/A, B | BMI |
| 391 | Health Behavior in School-aged Children (HBSC) | Ireland | 2001 | 2819 | 11–16 | School-based survey | Ha, S/A, B | BMI |
| 392 | Health Behavior in School-aged Children (HBSC) | Ireland | 2006 | 4746 | 11–16 | School-based survey | Ha, S/A, B | BMI |
| 393 | Health Behavior in School-aged Children (HBSC) | Ireland | 2010 | 4526 | 11–16 | School-based survey | Ha, S/A, B | BMI |
| 394 | Health Behavior in School-aged Children (HBSC) | Ireland | 2014 | 4002 | 11–16 | School-based survey | Ha, S/A, B | BMI |
| 395 | Health Behavior in School-aged Children (HBSC) | Ireland | 2018 | 3740 | 11–16 | School-based survey | Ha, S/A, B | BMI |
| 396 | Health Behavior in School-aged Children (HBSC) | Israel | 2001 | 5195 | 11–17 | School-based survey | Ha, S/A, B | BMI |
| 397 | Health Behavior in School-aged Children (HBSC) | Israel | 2006 | 4819 | 11–16 | School-based survey | Ha, S/A, B | BMI |
| 398 | Health Behavior in School-aged Children (HBSC) | Israel | 2010 | 3806 | 11–16 | School-based survey | Ha, S/A, B | BMI |
| 399 | Health Behavior in School-aged Children (HBSC) | Israel | 2014 | 5225 | 11–16 | School-based survey | Ha, S/A, B | BMI |
| 400 | Health Behavior in School-aged Children (HBSC) | Israel | 2018 | 7712 | 11–16 | School-based survey | Ha, S/A, B | BMI |
| 401 | Health Behavior in School-aged Children (HBSC) | Italy | 2001 | 4329 | 11–17 | School-based survey | Ha, S/A, B | BMI |
| 402 | Health Behavior in School-aged Children (HBSC) | Italy | 2006 | 3892 | 11–16 | School-based survey | Ha, S/A, B | BMI |
| 403 | Health Behavior in School-aged Children (HBSC) | Italy | 2010 | 4753 | 10–16 | School-based survey | Ha, S/A, B | BMI |
| 404 | Health Behavior in School-aged Children (HBSC) | Italy | 2014 | 3965 | 11–16 | School-based survey | Ha, S/A, B | BMI |
| 405 | Health Behavior in School-aged Children (HBSC) | Italy | 2018 | 4092 | 11–16 | School-based survey | Ha, S/A, B | BMI |
| 406 | Health Behavior in School-aged Children (HBSC) | Kazakhstan | 2018 | 4499 | 10–16 | School-based survey | Ha, S/A, B | BMI |
| 407 | Health Behavior in School-aged Children (HBSC) | Latvia | 2001 | 3305 | 11–16 | School-based survey | Ha, S/A, B | BMI |
| 408 | Health Behavior in School-aged Children (HBSC) | Latvia | 2006 | 4173 | 11–16 | School-based survey | Ha, S/A, B | BMI |
| 409 | Health Behavior in School-aged Children (HBSC) | Latvia | 2010 | 4110 | 11–16 | School-based survey | Ha, S/A, B | BMI |
| 410 | Health Behavior in School-aged Children (HBSC) | Latvia | 2014 | 5441 | 11–16 | School-based survey | Ha, S/A, B | BMI |
| 411 | Health Behavior in School-aged Children (HBSC) | Latvia | 2018 | 4348 | 11–16 | School-based survey | Ha, S/A, B | BMI |
| 412 | Health Behavior in School-aged Children (HBSC) | Lithuania | 2001 | 5549 | 11–17 | School-based survey | Ha, S/A, B | BMI |

| | | | | | | | | |
|---|---|---|---|---|---|---|---|---|
| 413 | Health Behavior in School-aged Children (HBSC) | Lithuania | 2006 | 5505 | 11–16 | School-based survey | Ha, S/A, B | BMI |
| 414 | Health Behavior in School-aged Children (HBSC) | Lithuania | 2010 | 5251 | 11–16 | School-based survey | Ha, S/A, B | BMI |
| 415 | Health Behavior in School-aged Children (HBSC) | Lithuania | 2018 | 3729 | 10–16 | School-based survey | Ha, S/A, B | BMI |
| 416 | Health Behavior in School-aged Children (HBSC) | Luxembourg | 2006 | 4213 | 11–16 | School-based survey | Ha, S/A, B | BMI |
| 417 | Health Behavior in School-aged Children (HBSC) | Luxembourg | 2010 | 3961 | 10–16 | School-based survey | Ha, S/A, B | BMI |
| 418 | Health Behavior in School-aged Children (HBSC) | Luxembourg | 2014 | 3062 | 11–16 | School-based survey | Ha, S/A, B | BMI |
| 419 | Health Behavior in School-aged Children (HBSC) | Luxembourg | 2018 | 3803 | 10–16 | School-based survey | Ha, S/A, B | BMI |
| 420 | Health Behavior in School-aged Children (HBSC) | Malta | 2001 | 1863 | 11–17 | School-based survey | Ha, S/A, B | BMI |
| 421 | Health Behavior in School-aged Children (HBSC) | Malta | 2006 | 1333 | 11–16 | School-based survey | Ha, S/A, B | BMI |
| 422 | Health Behavior in School-aged Children (HBSC) | Malta | 2014 | 2167 | 11–16 | School-based survey | Ha, S/A, B | BMI |
| 423 | Health Behavior in School-aged Children (HBSC) | Malta | 2018 | 2499 | 11–16 | School-based survey | Ha, S/A, B | BMI |
| 424 | Health Behavior in School-aged Children (HBSC) | Moldova | 2014 | 4642 | 11–16 | School-based survey | Ha, S/A, B | BMI |
| 425 | Health Behavior in School-aged Children (HBSC) | Moldova | 2018 | 4536 | 11–16 | School-based survey | Ha, S/A, B | BMI |
| 426 | Health Behavior in School-aged Children (HBSC) | Netherlands | 2001 | 4206 | 11–16 | School-based survey | Ha, S/A, B | BMI |
| 427 | Health Behavior in School-aged Children (HBSC) | Netherlands | 2006 | 4131 | 11–16 | School-based survey | Ha, S/A, B | BMI |
| 428 | Health Behavior in School-aged Children (HBSC) | Netherlands | 2010 | 4449 | 11–16 | School-based survey | Ha, S/A, B | BMI |
| 429 | Health Behavior in School-aged Children (HBSC) | Netherlands | 2014 | 4167 | 11–16 | School-based survey | Ha, S/A, B | BMI |
| 430 | Health Behavior in School-aged Children (HBSC) | Netherlands | 2018 | 4669 | 11–16 | School-based survey | Ha, S/A, B | BMI |
| 431 | Health Behavior in School-aged Children (HBSC) | North Macedonia | 2001 | 3933 | 10–16 | School-based survey | Ha, S/A, B | BMI |
| 432 | Health Behavior in School-aged Children (HBSC) | North Macedonia | 2006 | 5202 | 11–16 | School-based survey | Ha, S/A, B | BMI |
| 433 | Health Behavior in School-aged Children (HBSC) | North Macedonia | 2010 | 3702 | 10–16 | School-based survey | Ha, S/A, B | BMI |
| 434 | Health Behavior in School-aged Children (HBSC) | North Macedonia | 2014 | 3914 | 11–16 | School-based survey | Ha, S/A, B | BMI |
| 435 | Health Behavior in School-aged Children (HBSC) | North Macedonia | 2018 | 4208 | 11–16 | School-based survey | Ha, S/A, B | BMI |
| 436 | Health Behavior in School-aged Children (HBSC) | Norway | 2001 | 4886 | 10–16 | School-based survey | Ha, S/A, B | BMI |
| 437 | Health Behavior in School-aged Children (HBSC) | Norway | 2006 | 4549 | 11–16 | School-based survey | Ha, S/A, B | BMI |
| 438 | Health Behavior in School-aged Children (HBSC) | Norway | 2010 | 4254 | 11–16 | School-based survey | Ha, S/A, B | BMI |
| 439 | Health Behavior in School-aged Children (HBSC) | Norway | 2014 | 3208 | 11–16 | School-based survey | Ha, S/A, B | BMI |
| 440 | Health Behavior in School-aged Children (HBSC) | Norway | 2018 | 3044 | 11–16 | School-based survey | Ha, S/A, B | BMI |
| 441 | Health Behavior in School-aged Children (HBSC) | Poland | 2001 | 6245 | 11–17 | School-based survey | Ha, S/A, B | BMI |
| 442 | Health Behavior in School-aged Children (HBSC) | Poland | 2006 | 5466 | 11–16 | School-based survey | Ha, S/A, B | BMI |
| 443 | Health Behavior in School-aged Children (HBSC) | Poland | 2010 | 4203 | 11–16 | School-based survey | Ha, S/A, B | BMI |
| 444 | Health Behavior in School-aged Children (HBSC) | Poland | 2014 | 4427 | 11–16 | School-based survey | Ha, S/A, B | BMI |
| 445 | Health Behavior in School-aged Children (HBSC) | Poland | 2018 | 5097 | 11–16 | School-based survey | Ha, S/A, B | BMI |

| | | | | | | | | |
|---|---|---|---|---|---|---|---|---|
| 446 | Health Behavior in School-aged Children (HBSC) | Portugal | 2001 | 2835 | 11–17 | School-based survey | Ha, S/A, B | BMI |
| 447 | Health Behavior in School-aged Children (HBSC) | Portugal | 2006 | 3880 | 11–16 | School-based survey | Ha, S/A, B | BMI |
| 448 | Health Behavior in School-aged Children (HBSC) | Portugal | 2010 | 4002 | 10–16 | School-based survey | Ha, S/A, B | BMI |
| 449 | Health Behavior in School-aged Children (HBSC) | Portugal | 2014 | 4771 | 11–16 | School-based survey | Ha, S/A, B | BMI |
| 450 | Health Behavior in School-aged Children (HBSC) | Portugal | 2018 | 5756 | 11–16 | School-based survey | Ha, S/A, B | BMI |
| 451 | Health Behavior in School-aged Children (HBSC) | Romania | 2006 | 4538 | 11–16 | School-based survey | Ha, S/A, B | BMI |
| 452 | Health Behavior in School-aged Children (HBSC) | Romania | 2010 | 5119 | 11–16 | School-based survey | Ha, S/A, B | BMI |
| 453 | Health Behavior in School-aged Children (HBSC) | Romania | 2014 | 3785 | 10–16 | School-based survey | Ha, S/A, B | BMI |
| 454 | Health Behavior in School-aged Children (HBSC) | Romania | 2018 | 4371 | 10–16 | School-based survey | Ha, S/A, B | BMI |
| 455 | Health Behavior in School-aged Children (HBSC) | Russian Federation | 2001 | 7965 | 11–17 | School-based survey | Ha, S/A, B | BMI |
| 456 | Health Behavior in School-aged Children (HBSC) | Russian Federation | 2006 | 7911 | 11–16 | School-based survey | Ha, S/A, B | BMI |
| 457 | Health Behavior in School-aged Children (HBSC) | Russian Federation | 2010 | 4999 | 11–16 | School-based survey | Ha, S/A, B | BMI |
| 458 | Health Behavior in School-aged Children (HBSC) | Russian Federation | 2014 | 4394 | 11–16 | School-based survey | Ha, S/A, B | BMI |
| 459 | Health Behavior in School-aged Children (HBSC) | Russian Federation | 2018 | 4215 | 10–16 | School-based survey | Ha, S/A, B | BMI |
| 460 | Health Behavior in School-aged Children (HBSC) | Serbia | 2018 | 3789 | 11–16 | School-based survey | Ha, S/A, B | BMI |
| 461 | Health Behavior in School-aged Children (HBSC) | Slovakia | 2006 | 3661 | 10–16 | School-based survey | Ha, S/A, B | BMI |
| 462 | Health Behavior in School-aged Children (HBSC) | Slovakia | 2010 | 5195 | 11–16 | School-based survey | Ha, S/A, B | BMI |
| 463 | Health Behavior in School-aged Children (HBSC) | Slovakia | 2014 | 5933 | 11–16 | School-based survey | Ha, S/A, B | BMI |
| 464 | Health Behavior in School-aged Children (HBSC) | Slovakia | 2018 | 4512 | 11–16 | School-based survey | Ha, S/A, B | BMI |
| 465 | Health Behavior in School-aged Children (HBSC) | Slovenia | 2001 | 3887 | 11–17 | School-based survey | Ha, S/A, B | BMI |
| 466 | Health Behavior in School-aged Children (HBSC) | Slovenia | 2006 | 5088 | 11–16 | School-based survey | Ha, S/A, B | BMI |
| 467 | Health Behavior in School-aged Children (HBSC) | Slovenia | 2010 | 5395 | 11–16 | School-based survey | Ha, S/A, B | BMI |
| 468 | Health Behavior in School-aged Children (HBSC) | Slovenia | 2014 | 4936 | 11–16 | School-based survey | Ha, S/A, B | BMI |
| 469 | Health Behavior in School-aged Children (HBSC) | Slovenia | 2018 | 5603 | 11–16 | School-based survey | Ha, S/A, B | BMI |
| 470 | Health Behavior in School-aged Children (HBSC) | Spain | 2001 | 5748 | 10–16 | School-based survey | Ha, S/A, B | BMI |
| 471 | Health Behavior in School-aged Children (HBSC) | Spain | 2006 | 8735 | 11–16 | School-based survey | Ha, S/A, B | BMI |
| 472 | Health Behavior in School-aged Children (HBSC) | Spain | 2010 | 4969 | 10–16 | School-based survey | Ha, S/A, B | BMI |
| 473 | Health Behavior in School-aged Children (HBSC) | Spain | 2014 | 9966 | 10–16 | School-based survey | Ha, S/A, B | BMI |
| 474 | Health Behavior in School-aged Children (HBSC) | Spain | 2018 | 4286 | 10–16 | School-based survey | Ha, S/A, B | BMI |
| 475 | Health Behavior in School-aged Children (HBSC) | Sweden | 2001 | 3748 | 10–16 | School-based survey | Ha, S/A, B | BMI |
| 476 | Health Behavior in School-aged Children (HBSC) | Sweden | 2006 | 4282 | 10–16 | School-based survey | Ha, S/A, B | BMI |
| 477 | Health Behavior in School-aged Children (HBSC) | Sweden | 2010 | 6451 | 10–16 | School-based survey | Ha, S/A, B | BMI |
| 478 | Health Behavior in School-aged Children (HBSC) | Sweden | 2014 | 7497 | 11–16 | School-based survey | Ha, S/A, B | BMI |

| | | | | | | | | |
|---|---|---|---|---|---|---|---|---|
| 479 | Health Behavior in School-aged Children (HBSC) | Sweden | 2018 | 4031 | 11–16 | School-based survey | Ha, S/A, B | BMI |
| 480 | Health Behavior in School-aged Children (HBSC) | Switzerland | 2001 | 4505 | 11–17 | School-based survey | Ha, S/A, B | BMI |
| 481 | Health Behavior in School-aged Children (HBSC) | Switzerland | 2006 | 4462 | 10–16 | School-based survey | Ha, S/A, B | BMI |
| 482 | Health Behavior in School-aged Children (HBSC) | Switzerland | 2010 | 6470 | 10–16 | School-based survey | Ha, S/A, B | BMI |
| 483 | Health Behavior in School-aged Children (HBSC) | Switzerland | 2014 | 6399 | 10–16 | School-based survey | Ha, S/A, B | BMI |
| 484 | Health Behavior in School-aged Children (HBSC) | Switzerland | 2018 | 7409 | 10–16 | School-based survey | Ha, S/A, B | BMI |
| 485 | Health Behavior in School-aged Children (HBSC) | Turkiye | 2006 | 5268 | 11–16 | School-based survey | Ha, S/A, B | BMI |
| 486 | Health Behavior in School-aged Children (HBSC) | Turkiye | 2010 | 5456 | 11–16 | School-based survey | Ha, S/A, B | BMI |
| 487 | Health Behavior in School-aged Children (HBSC) | Turkiye | 2018 | 5654 | 11–16 | School-based survey | Ha, S/A, B | BMI |
| 488 | Health Behavior in School-aged Children (HBSC) | Ukraine | 2001 | 3990 | 11–17 | School-based survey | Ha, S/A, B | BMI |
| 489 | Health Behavior in School-aged Children (HBSC) | Ukraine | 2006 | 4837 | 11–16 | School-based survey | Ha, S/A, B | BMI |
| 490 | Health Behavior in School-aged Children (HBSC) | Ukraine | 2010 | 5726 | 11–16 | School-based survey | Ha, S/A, B | BMI |
| 491 | Health Behavior in School-aged Children (HBSC) | Ukraine | 2014 | 4312 | 11–16 | School-based survey | Ha, S/A, B | BMI |
| 492 | Health Behavior in School-aged Children (HBSC) | Ukraine | 2018 | 6380 | 10–16 | School-based survey | Ha, S/A, B | BMI |
| 493 | Health Behavior in School-aged Children (HBSC) | United Kingdom | 2001 | 13907 | 11–17 | School-based survey | Ha, S/A, B | BMI |
| 494 | Health Behavior in School-aged Children (HBSC) | United Kingdom | 2006 | 15021 | 11–17 | School-based survey | Ha, S/A, B | BMI |
| 495 | Health Behavior in School-aged Children (HBSC) | United Kingdom | 2010 | 15202 | 10–16 | School-based survey | Ha, S/A, B | BMI |
| 496 | Health Behavior in School-aged Children (HBSC) | United Kingdom | 2014 | 15828 | 11–16 | School-based survey | Ha, S/A, B | BMI |
| 497 | Health Behavior in School-aged Children (HBSC) | United Kingdom | 2018 | 23511 | 11–16 | School-based survey | Ha, S/A, B | BMI |
| 498 | Health Behavior in School-aged Children (HBSC) | United States | 1995 | 9611 | 9–18 | School-based survey | Ha, S/A, B | |
| 499 | Health Behavior in School-aged Children (HBSC) | United States | 1997 | 15558 | 11–17 | School-based survey | Ha, S/A, B | BMI |
| 500 | Health Behavior in School-aged Children (HBSC) | United States | 2001 | 4939 | 11–16 | School-based survey | Ha, S/A, B | BMI |
| 501 | Health Behavior in School-aged Children (HBSC) | United States | 2006 | 3863 | 11–16 | School-based survey | Ha, S/A, B | BMI |
| 502 | Health Behavior in School-aged Children (HBSC) | United States | 2010 | 6051 | 11–16 | School-based survey | Ha, S/A, B | BMI |
| 503 | Health and Retirement Study (HRS) | United States | 2004 | 19926 | 24–100 | Panel | P, Ha, B | BMI |
| 504 | Health and Retirement Study (HRS) | United States | 2008 | 830 | 25–100 | Panel | P, Ha, B | BMI; Income |
| 505 | Health and Retirement Study (HRS) | United States | 2010 | 6599 | 18–96 | Panel | P, Ha, B | BMI; Income |
| 506 | Health and Retirement Study (HRS) | United States | 2012 | 384 | 12–91 | Panel | P, Ha, B | BMI; Income |
| 507 | Health and Retirement Study (HRS) | United States | 2016 | 4714 | 21–90 | Panel | P, Ha, B | BMI |
| 508 | Health and Retirement Study (HRS) | United States | 2020 | 332 | 28–92 | Panel | P, Ha, B | BMI |
| 509 | Health and Retirement Study (HRS) | United States | 2022 | 2840 | 29–100 | Panel | P, Ha, B | BMI |
| 510 | Health and Retirement Study in Thailand (HART) | Thailand | 2014 | 7987 | 44–100 | Panel | P, Ha, N/S, C, S/A, B, K, H, Hnd, Ft | BMI; Smoking; Income |

| | | | | | | | | |
|---|---|---|---|---|---|---|---|---|
| 511 | Healthy Aging in Africa (HAALSI) | South Africa | 2014 | 3914 | 43–100 | Panel | P, Ha, N/S, C, S/A, B, K, H, F, Hnd, Ft, E | BMI |
| 512 | Household Income Expenditure Survey | Kiribati | 2019 | 12481 | 5–80 | Household survey | Ha, S/A | |
| 513 | Household Income Expenditure Survey | Tonga | 2021 | 11061 | 5–80 | Household survey | Ha, S/A | |
| 514 | INDEPTH | Ghana | 2006 | 593 | 50–100 | Surveillance/HDSS | P, B | BMI |
| 515 | INDEPTH | India | 2006 | 373 | 50–90 | Surveillance/HDSS | P, B | BMI |
| 516 | INDEPTH | South Africa | 2006 | 424 | 50–98 | Surveillance/HDSS | P, B | BMI |
| 517 | Ibadan Study on Aging | Nigeria | 2003 | 2014 | 65–100 | Household survey | P, Ha, C, S/A, B, F | BMI |
| 518 | Imaging Genetics Study of Adolescents (IMAGEN) | Germany | 2011 | 1207 | 14–18 | Cohort | P, Ha, C, S/A, B | BMI |
| 519 | Indonesian Family Life Survey (IFLS) | Indonesia | 2008 | 5266 | 15–100 | Household panel | P, Ha, N/S, B, K, H, Hnd, Ft | BMI; Smoking; Income |
| 520 | Indonesian Family Life Survey (IFLS) | Indonesia | 2015 | 34457 | 14–100 | Household panel | P, Ha, N/S, B, K, H, Hnd, Ft | BMI; Smoking; Income |
| 521 | Iranian Back Pain Study | Iran | 2014 | 3521 | 16–90 | Cross-section | B | BMI |
| 522 | Japan COVID-19 and Society Internet Survey (JACSIS) | Japan | 2020 | 8209 | 18–49 | Cross-section | P, Ha, N/S, S/A, H | BMI; Smoking; Income |
| 523 | Japan Gerontological Evaluation Study (JAGES) | Japan | 2019 | 19181 | 65–95 | Panel | P, N/S, B, K, H, Hnd, Ft, E | BMI; Smoking; Income |
| 524 | Japan Gerontological Evaluation Study (JAGES) | Japan | 2022 | 20712 | 24–100 | Panel | P, N/S, B, K, H, Hnd, Ft, E | Smoking; Income |
| 525 | Japan Neuropathic Pain Survey (JNeP) | Japan | 2014 | 5553 | 18–89 | Cross-section | P, Ha, N/S, S/A, B, K, H, F, Hnd, Ft, E | Smoking |
| 526 | Japanese Chronic Pain Survey (JCP) | Japan | 2014 | 2682 | 20–99 | Cross-section | P, Ha, N/S, S/A, B, K, H, F, Hnd, Ft, E | |
| 527 | Khayelitsha | South Africa | 2004 | 3300 | 16–92 | Household survey | P, Ha, C, S/A, B | BMI; Smoking; Income |
| 528 | Korean General Social Survey (KGSS) | South Korea | 2010 | 1570 | 18–92 | Cross-section | P, N/S, B, K, Hnd | BMI; Income |
| 529 | Korean Longitudinal Study on Aging (KLoSA) | South Korea | 2005 | 11174 | 45–98 | Panel | P, Ha, N/S, C, S/A, B, K, H, Hnd, Ft | BMI; Smoking; Income |
| 530 | Libyan Telephone Pain Survey | Libya | 2011 | 1206 | 20–75 | Household survey | P, Ha, N/S, C, S/A, B, K, H, Hnd, Ft | |
| 531 | LifeLines | Netherlands | 2006 | 92438 | 5–96 | Cohort | P, Ha, N/S, C, S/A, B, H, F | BMI; Smoking; Income |
| 532 | Living Standards Measurement Survey (LSMS)[e] | Kazakhstan | 1996 | 2038 | 5–17 | Household survey | Ha | |
| 533 | Longitudinal Ageing Study in India (LASI) | India | 2017 | 73157 | 18–100 | Panel | P, Ha, C, B | BMI; Smoking |
| 534 | Longitudinal Study of Australian Children (LSAC)[2] | Australia | 2004 | 3244 | 15–20 | Cohort | P, Ha, N/S, C, S/A, B, K, H, Hnd, Ft, E | BMI; Income |
| 535 | MAmMOTH (EpiGroup) | United Kingdom | 2015 | 17888 | 17–100 | Cohort | P, Ha, N/S, C, S/A, B, K, H, F, Hnd, Ft, E | |
| 536 | MUSICIAN (EpiGroup) | United Kingdom | 2010 | 15302 | 18–100 | Cohort | P, Ha, N/S, C, S/A, B, K, H, Hnd, Ft, E | BMI |

| | | | | | | | | |
|---|---|---|---|---|---|---|---|---|
| 537 | Malaysian Aging and Retirement Study (MARS) | Malaysia | 2018 | 5613 | 40–95 | Cross-section | P, Ha, N/S, C, S/A, B, K, H, Hnd, Ft | BMI; Smoking; Income |
| 538 | Manicaland General Population Cohort | Zimbabwe | 2014 | 2838 | 5–17 | Household survey | Ha | |
| 539 | MeMaps | Germany | 2019 | 1562 | 8–17 | Cohort | P, Ha, C, S/A, B | BMI |
| 540 | Mexican Family Life Survey (ENNVIH) | Mexico | 2002 | 19779 | 15–98 | Household panel | P, Ha, C, S/A | BMI; Smoking |
| 541 | Mexican Family Life Survey (ENNVIH) | Mexico | 2005 | 20319 | 14–99 | Household panel | P, Ha, C, S/A | BMI; Smoking |
| 542 | Mexican Family Life Survey (ENNVIH) | Mexico | 2010 | 23208 | 11–99 | Household panel | P, Ha, C, S/A | BMI; Smoking |
| 543 | Mexican Health and Aging Study (MHAS) | Mexico | 2001 | 14981 | 18–100 | Panel | P, S/A | BMI; Income |
| 544 | Mexican Health and Aging Study (MHAS) | Mexico | 2003 | 162 | 33–99 | Panel | P, S/A | BMI; Income |
| 545 | Mexican Health and Aging Study (MHAS) | Mexico | 2012 | 5716 | 21–100 | Panel | P, S/A | BMI; Income |
| 546 | Mexican Health and Aging Study (MHAS) | Mexico | 2015 | 499 | 30–87 | Panel | P, S/A | BMI; Income |
| 547 | Mexican Health and Aging Study (MHAS) | Mexico | 2018 | 4762 | 16–86 | Panel | P, S/A | BMI; Income |
| 548 | Mexican Health and Aging Study (MHAS) | Mexico | 2021 | 156 | 39–99 | Panel | P, S/A | BMI; Income |
| 549 | Midlife in Japan Study (MIDJA) | Japan | 2008 | 1025 | 30–79 | Cohort | P, Ha, B | Smoking |
| 550 | Midlife in the United States (MIDUS) | United States | 2009 | 6523 | 23–84 | Panel | P, Ha, N/S, C, B, K, H | BMI; Smoking; Income |
| 551 | Murakami Cohort Study[f] | Japan | 2012 | 14205 | 42–72 | Cohort | P, Ha, N/S, B, K, Hnd, E | |
| 552 | National Childhood Development Study (NCDS) | United Kingdom | 2002 | 8499 | 44–46 | Cohort | P, Ha, N/S, C, S/A, B, K, H, Hnd, Ft | BMI; Smoking |
| 553 | National Health Interview Survey (NHIS) | United States | 1997 | 36051 | 18–85 | Repeat cross-section | P, Ha, N/S, B, K, H, F, Hnd, Ft, E | BMI; Smoking; Income |
| 554 | National Health Interview Survey (NHIS) | United States | 1998 | 32407 | 18–85 | Repeat cross-section | P, Ha, N/S, B, K, H, F, Hnd, Ft, E | BMI; Smoking; Income |
| 555 | National Health Interview Survey (NHIS) | United States | 1999 | 30791 | 18–85 | Repeat cross-section | P, Ha, N/S, B, K, H, F, Hnd, Ft, E | BMI; Smoking; Income |
| 556 | National Health Interview Survey (NHIS) | United States | 2000 | 32347 | 18–85 | Repeat cross-section | P, Ha, N/S, B, K, H, F, Hnd, Ft, E | BMI; Smoking; Income |
| 557 | National Health Interview Survey (NHIS) | United States | 2001 | 33310 | 18–85 | Repeat cross-section | P, Ha, N/S, B, K, H, F, Hnd, Ft, E | BMI; Smoking; Income |
| 558 | National Health Interview Survey (NHIS) | United States | 2002 | 31008 | 18–85 | Repeat cross-section | P, Ha, N/S, B, K, H, F, Hnd, Ft, E | BMI; Smoking; Income |
| 559 | National Health Interview Survey (NHIS) | United States | 2003 | 30824 | 18–85 | Repeat cross-section | P, Ha, N/S, B, K, H, F, Hnd, Ft, E | BMI; Smoking; Income |
| 560 | National Health Interview Survey (NHIS) | United States | 2004 | 31282 | 18–85 | Repeat cross-section | P, Ha, N/S, B, K, H, F, Hnd, Ft, E | BMI; Smoking; Income |

| | | | | | | | | |
|---|---|---|---|---|---|---|---|---|
| 561 | National Health Interview Survey (NHIS) | United States | 2005 | 31409 | 18–85 | Repeat cross-section | P, Ha, N/S, B, K, H, F, Hnd, Ft, E | BMI; Smoking; Income |
| 562 | National Health Interview Survey (NHIS) | United States | 2006 | 24244 | 18–85 | Repeat cross-section | P, Ha, N/S, B, K, H, F, Hnd, Ft, E | BMI; Smoking; Income |
| 563 | National Health Interview Survey (NHIS) | United States | 2007 | 23381 | 18–85 | Repeat cross-section | P, Ha, N/S, B, K, H, F, Hnd, Ft, E | BMI; Smoking; Income |
| 564 | National Health Interview Survey (NHIS) | United States | 2008 | 21777 | 18–85 | Repeat cross-section | P, Ha, N/S, B, K, H, F, Hnd, Ft, E | BMI; Smoking; Income |
| 565 | National Health Interview Survey (NHIS) | United States | 2009 | 27721 | 18–85 | Repeat cross-section | P, Ha, N/S, B, K, H, F, Hnd, Ft, E | BMI; Smoking; Income |
| 566 | National Health Interview Survey (NHIS) | United States | 2010 | 27152 | 18–85 | Repeat cross-section | P, Ha, N/S, B, K, H, F, Hnd, Ft, E | BMI; Smoking; Income |
| 567 | National Health Interview Survey (NHIS) | United States | 2011 | 33002 | 18–85 | Repeat cross-section | P, Ha, N/S, B, K, H, F, Hnd, Ft, E | BMI; Smoking; Income |
| 568 | National Health Interview Survey (NHIS) | United States | 2012 | 34514 | 18–85 | Repeat cross-section | P, Ha, N/S, B, K, H, F, Hnd, Ft, E | BMI; Smoking; Income |
| 569 | National Health Interview Survey (NHIS) | United States | 2013 | 34542 | 18–85 | Repeat cross-section | P, Ha, N/S, B, K, H, F, Hnd, Ft, E | BMI; Smoking; Income |
| 570 | National Health Interview Survey (NHIS) | United States | 2014 | 36683 | 18–85 | Repeat cross-section | P, Ha, N/S, B, K, H, F, Hnd, Ft, E | BMI; Smoking; Income |
| 571 | National Health Interview Survey (NHIS) | United States | 2015 | 33655 | 18–85 | Repeat cross-section | P, Ha, N/S, B, K, H, F, Hnd, Ft, E | BMI; Smoking; Income |
| 572 | National Health Interview Survey (NHIS) | United States | 2016 | 33018 | 18–85 | Repeat cross-section | P, Ha, N/S, B, K, H, F, Hnd, Ft, E | BMI; Smoking; Income |
| 573 | National Health Interview Survey (NHIS) | United States | 2017 | 26735 | 18–85 | Repeat cross-section | P, Ha, N/S, B, K, H, F, Hnd, Ft, E | BMI; Smoking; Income |
| 574 | National Health Interview Survey (NHIS) | United States | 2018 | 25407 | 18–85 | Repeat cross-section | P, Ha, N/S, B, K, H, F, Hnd, Ft, E | BMI; Smoking; Income |
| 575 | National Health Interview Survey (NHIS) | United States | 2019 | 31200 | 18–85 | Repeat cross-section | P, Ha, S/A, B, F | Smoking |
| 576 | National Health Interview Survey (NHIS) | United States | 2021 | 28671 | 18–85 | Repeat cross-section | P, Ha, S/A, B, F | Smoking |
| 577 | National Health Interview Survey (NHIS) | United States | 2023 | 28512 | 18–85 | Repeat cross-section | P, Ha, S/A, B, F | Smoking |
| 578 | National Health Measurement Survey 2010 (NHMS) | United States | 2008 | 3844 | 35–89 | Cross-section | P, Ha, C, S/A, B, H, F | BMI; Smoking; Income |
| 579 | National Health and Aging Trends Study (NHATS) | United States | 2011 | 17285 | 67–92 | Panel | P, Ha, N/S, S/A, B, K, H, Hnd, Ft | BMI; Smoking; Income |
| 580 | National Health and Nutrition Examination Survey (NHANES) | United States | 1999 | 4878 | 20–85 | Repeated cross-section | P, Ha, N/S, S/A, B, K, H, F, Hnd, Ft | BMI; Smoking; Income |

| | | | | | | | | |
|---|---|---|---|---|---|---|---|---|
| 581 | National Health and Nutrition Examination Survey (NHANES) | United States | 2001 | 5410 | 20–85 | Repeated cross-section | P, Ha, N/S, C, S/A, B, K, H, F, Hnd, Ft | BMI; Smoking; Income |
| 582 | National Health and Nutrition Examination Survey (NHANES) | United States | 2003 | 5040 | 20–85 | Repeated cross-section | P, Ha, N/S, C, S/A, B, K, H, F, Hnd, Ft | BMI; Smoking; Income |
| 583 | National Health and Nutrition Examination Survey (NHANES) | United States | 2009 | 5106 | 20–69 | Repeated cross-section | P, N/S, C, B, H | BMI; Smoking; Income |
| 584 | National Income Dynamics Study (NIDS) | South Africa | 2008 | 676 | 15–100 | Panel | P, Ha, C, S/A, B | BMI; Smoking; Income |
| 585 | National Income Dynamics Study (NIDS) | South Africa | 2010 | 1752 | 15–100 | Panel | P, Ha, C, S/A, B | BMI; Smoking; Income |
| 586 | National Income Dynamics Study (NIDS) | South Africa | 2012 | 2552 | 15–100 | Panel | P, Ha, C, S/A, B | BMI; Smoking; Income |
| 587 | National Income Dynamics Study (NIDS) | South Africa | 2014 | 4111 | 15–100 | Panel | P, Ha, C, B | BMI; Smoking; Income |
| 588 | National Income Dynamics Study (NIDS) | South Africa | 2017 | 23869 | 14–100 | Panel | P, Ha, C, B | BMI; Smoking; Income |
| 589 | National Social Life, Health, and Aging Project (NSHAP) | United States | 2010 | 2600 | 62–91 | Panel | P, Ha, N/S, C, S/A, B, Hnd, Ft | BMI; Smoking; Income |
| 590 | National Survey of Health and Nutrition in Mexico 2000 (ENSANUT) | Mexico | 2000 | 44095 | 20–100 | Household survey | P, Ha, N/S, K, H, Hnd, Ft, E | BMI; Smoking |
| 591 | National Survey of Self-Care and Aging (NSSA) | United States | 1994 | 3482 | 68–100 | Household survey | P, N/S, S/A, B, K, H, Hnd, Ft, E | Smoking; Income |
| 592 | New Zealand Health Survey (NZHS)[f] | New Zealand | 2017 | 13598 | 20–85 | Household survey | P, Ha, C, S/A, B | |
| 593 | Ontario Health Study (OHS) | Canada | 2015 | 20051 | 18–100 | Cohort | P, N/S, B, K, H, F, Hnd, Ft, E | BMI; Smoking; Income |
| 594 | Orofacial Pain: Prospective Evaluation and Risk Assessment (OPPERA) | United States | 2015 | 655 | 22–53 | Cohort | P, N/S, C, B, K, F, Hnd, Ft, E | |
| 595 | PREVENT-AD | Canada | 2021 | 339 | 56–88 | Cohort | P, Ha, N/S, C, S/A, B, K, H, Hnd, Ft, E | Smoking |
| 596 | Pain in Schoolchildren Study (PSC) | United Kingdom | 2003 | 1442 | 11–16 | School-based Survey | P, Ha, N/S, C, S/A, B, K, H, Hnd, Ft, E | BMI |
| 597 | Pain-Associated Cross-Sectional Epidemiological (PACE) Study[3] | Japan | 2009 | 5000 | 20–79 | Cross-section | P, Ha, N/S, S/A, B, K, H, F, Hnd, Ft, E | |
| 598 | Puerto Rican Elderly: Health Conditions Project (PREHCO) | Puerto Rico | 2002 | 3713 | 60–100 | Household survey | P, C, B | BMI; Smoking; Income |
| 599 | SHAMA (EpiGroup) | United Kingdom | 2015 | 1604 | 30–90 | Cross-section | P, Ha, N/S, C, S/A, B, K, H, F, Hnd, Ft, E | |
| 600 | Sacramento Area Latino Study on Aging (SALSA) | United States | 1996 | 1716 | 59–100 | Cohort | P, C, B, K, H, Hnd, Ft | BMI; Smoking; Income |
| 601 | Saudi Online Pain Survey | Saudi Arabia | 2018 | 26372 | 24–65 | Household survey | P, Ha, N/S, C, S/A, B | Income |

| | | | | | | | | |
|---|---|---|---|---|---|---|---|---|
| 602 | Scottish Family Health Study (SFHS) | United Kingdom | 2008 | 24073 | 18–100 | Cohort | P, Ha, N/S, C, S/A, B | BMI; Smoking; Income |
| 603 | Scottish Health Study (SHS) | United Kingdom | 2010 | 4656 | 12–97 | Household survey | P, C | BMI; Smoking; Income |
| 604 | Scottish Health Study (SHS) | United Kingdom | 2022 | 4732 | 12–97 | Household survey | P, Ha, N/S, C, S/A, B | BMI; Smoking; Income |
| 605 | Shandong Adolescent Behavior & Health Cohort (SABHC) | China | 2015 | 6571 | 10–20 | School-based survey | P, Ha, S/A | BMI; Income |
| 606 | South African National Demographic Survey 2016 (SADS) | South Africa | 2016 | 10336 | 15–95 | Household survey | P, Ha, N/S, C, S/A, B, F | Smoking |
| 607 | Spanish Chronic Pain Survey | Spain | 2013 | 1957 | 18–92 | Household survey | P, Ha, N/S, B | |
| 608 | Spanish Longitudinal Ageing Study (SLAS) | Spain | 2011 | 2378 | 21–100 | Cohort | P, B | BMI; Smoking; Income |
| 609 | Study of Health and Aging in Europe (SHARE) | Austria | 2019 | 1014 | 45–100 | Panel | P, B, K, H, F | BMI; Smoking; Income |
| 610 | Study of Health and Aging in Europe (SHARE) | Austria | 2020 | 938 | 44–98 | Panel | P, B, K, H, F | BMI; Smoking; Income |
| 611 | Study of Health and Aging in Europe (SHARE) | Austria | 2021 | 27 | 56–92 | Panel | P, B, K, H, F | BMI; Smoking; Income |
| 612 | Study of Health and Aging in Europe (SHARE) | Austria | 2022 | 1959 | 40–99 | Panel | P, B, K, H, F | BMI; Smoking; Income |
| 613 | Study of Health and Aging in Europe (SHARE) | Belgium | 2019 | 480 | 46–97 | Panel | P, B, K, H, F | BMI; Smoking; Income |
| 614 | Study of Health and Aging in Europe (SHARE) | Belgium | 2020 | 1790 | 36–99 | Panel | P, B, K, H, F | BMI; Smoking; Income |
| 615 | Study of Health and Aging in Europe (SHARE) | Belgium | 2021 | 222 | 47–91 | Panel | P, B, K, H, F | BMI; Smoking; Income |
| 616 | Study of Health and Aging in Europe (SHARE) | Belgium | 2022 | 2439 | 31–100 | Panel | P, B, K, H, F | BMI; Smoking; Income |
| 617 | Study of Health and Aging in Europe (SHARE) | Bulgaria | 2019 | 311 | 42–92 | Panel | P, B, K, H, F | BMI; Smoking; Income |
| 618 | Study of Health and Aging in Europe (SHARE) | Bulgaria | 2020 | 593 | 40–93 | Panel | P, B, K, H, F | BMI; Smoking; Income |
| 619 | Study of Health and Aging in Europe (SHARE) | Bulgaria | 2021 | 10 | 52–88 | Panel | P, B, K, H, F | BMI; Smoking; Income |
| 620 | Study of Health and Aging in Europe (SHARE) | Bulgaria | 2022 | 159 | 49–100 | Panel | P, B, K, H, F | BMI; Smoking; Income |
| 621 | Study of Health and Aging in Europe (SHARE) | Croatia | 2019 | 319 | 34–95 | Panel | P, B, K, H, F | BMI; Smoking; Income |

| | | | | | | | | |
|---|---|---|---|---|---|---|---|---|
| 622 | Study of Health and Aging in Europe (SHARE) | Croatia | 2020 | 1703 | 32–98 | Panel | P, B, K, H, F | BMI; Smoking; Income |
| 623 | Study of Health and Aging in Europe (SHARE) | Croatia | 2021 | 195 | 53–94 | Panel | P, B, K, H, F | BMI; Smoking; Income |
| 624 | Study of Health and Aging in Europe (SHARE) | Croatia | 2022 | 2749 | 36–100 | Panel | P, B, K, H, F | BMI; Smoking; Income |
| 625 | Study of Health and Aging in Europe (SHARE) | Cyprus | 2019 | 162 | 50–95 | Panel | P, B, K, H, F | BMI; Smoking; Income |
| 626 | Study of Health and Aging in Europe (SHARE) | Cyprus | 2020 | 372 | 51–99 | Panel | P, B, K, H, F | BMI; Smoking; Income |
| 627 | Study of Health and Aging in Europe (SHARE) | Cyprus | 2021 | 97 | 27–90 | Panel | P, B, K, H, F | BMI; Smoking; Income |
| 628 | Study of Health and Aging in Europe (SHARE) | Cyprus | 2022 | 295 | 46–98 | Panel | P, B, K, H, F | BMI; Smoking; Income |
| 629 | Study of Health and Aging in Europe (SHARE) | Czech Republic | 2019 | 1785 | 30–97 | Panel | P, B, K, H, F | BMI; Smoking; Income |
| 630 | Study of Health and Aging in Europe (SHARE) | Czech Republic | 2020 | 1246 | 38–96 | Panel | P, B, K, H, F | BMI; Smoking; Income |
| 631 | Study of Health and Aging in Europe (SHARE) | Czech Republic | 2021 | 41 | 54–93 | Panel | P, B, K, H, F | BMI; Smoking; Income |
| 632 | Study of Health and Aging in Europe (SHARE) | Czech Republic | 2022 | 1224 | 30–96 | Panel | P, B, K, H, F | BMI; Smoking; Income |
| 633 | Study of Health and Aging in Europe (SHARE) | Denmark | 2019 | 581 | 37–100 | Panel | P, B, K, H, F | BMI; Smoking; Income |
| 634 | Study of Health and Aging in Europe (SHARE) | Denmark | 2020 | 1783 | 40–98 | Panel | P, B, K, H, F | BMI; Smoking; Income |
| 635 | Study of Health and Aging in Europe (SHARE) | Denmark | 2021 | 40 | 54–88 | Panel | P, B, K, H, F | BMI; Smoking; Income |
| 636 | Study of Health and Aging in Europe (SHARE) | Denmark | 2022 | 625 | 20–97 | Panel | P, B, K, H, F | BMI; Smoking; Income |
| 637 | Study of Health and Aging in Europe (SHARE) | Estonia | 2019 | 1396 | 35–95 | Panel | P, B, K, H, F | BMI; Smoking; Income |
| 638 | Study of Health and Aging in Europe (SHARE) | Estonia | 2020 | 1998 | 40–100 | Panel | P, B, K, H, F | BMI; Smoking; Income |
| 639 | Study of Health and Aging in Europe (SHARE) | Estonia | 2021 | 370 | 43–95 | Panel | P, B, K, H, F | BMI; Smoking; Income |

| | | | | | | | | |
|---|---|---|---|---|---|---|---|---|
| 640 | Study of Health and Aging in Europe (SHARE) | Estonia | 2022 | 1372 | 40–97 | Panel | P, B, K, H, F | BMI; Smoking; Income |
| 641 | Study of Health and Aging in Europe (SHARE) | Finland | 2019 | 790 | 37–96 | Panel | P, B, K, H, F | BMI; Smoking; Income |
| 642 | Study of Health and Aging in Europe (SHARE) | Finland | 2020 | 370 | 42–92 | Panel | P, B, K, H, F | BMI; Smoking; Income |
| 643 | Study of Health and Aging in Europe (SHARE) | Finland | 2022 | 942 | 31–99 | Panel | P, B, K, H, F | BMI; Smoking; Income |
| 644 | Study of Health and Aging in Europe (SHARE) | France | 2019 | 1616 | 39–99 | Panel | P, B, K, H, F | BMI; Smoking; Income |
| 645 | Study of Health and Aging in Europe (SHARE) | France | 2020 | 1392 | 39–100 | Panel | P, B, K, H, F | BMI; Smoking; Income |
| 646 | Study of Health and Aging in Europe (SHARE) | France | 2021 | 65 | 52–96 | Panel | P, B, K, H, F | BMI; Smoking; Income |
| 647 | Study of Health and Aging in Europe (SHARE) | France | 2022 | 715 | 40–99 | Panel | P, B, K, H, F | BMI; Smoking; Income |
| 648 | Study of Health and Aging in Europe (SHARE) | Germany | 2019 | 1858 | 47–97 | Panel | P, B, K, H, F | BMI; Smoking; Income |
| 649 | Study of Health and Aging in Europe (SHARE) | Germany | 2020 | 2004 | 28–95 | Panel | P, B, K, H, F | BMI; Smoking; Income |
| 650 | Study of Health and Aging in Europe (SHARE) | Germany | 2021 | 84 | 46–93 | Panel | P, B, K, H, F | BMI; Smoking; Income |
| 651 | Study of Health and Aging in Europe (SHARE) | Germany | 2022 | 1547 | 38–97 | Panel | P, B, K, H, F | BMI; Smoking; Income |
| 652 | Study of Health and Aging in Europe (SHARE) | Greece | 2019 | 1016 | 46–96 | Panel | P, B, K, H, F | BMI; Smoking; Income |
| 653 | Study of Health and Aging in Europe (SHARE) | Greece | 2020 | 1979 | 42–98 | Panel | P, B, K, H, F | BMI; Smoking; Income |
| 654 | Study of Health and Aging in Europe (SHARE) | Greece | 2021 | 93 | 57–97 | Panel | P, B, K, H, F | BMI; Smoking; Income |
| 655 | Study of Health and Aging in Europe (SHARE) | Greece | 2022 | 600 | 45–95 | Panel | P, B, K, H, F | BMI; Smoking; Income |
| 656 | Study of Health and Aging in Europe (SHARE) | Hungary | 2019 | 107 | 50–94 | Panel | P, B, K, H, F | BMI; Smoking; Income |
| 657 | Study of Health and Aging in Europe (SHARE) | Hungary | 2020 | 1102 | 39–94 | Panel | P, B, K, H, F | BMI; Smoking; Income |

| | | | | | | | | |
|---|---|---|---|---|---|---|---|---|
| 658 | Study of Health and Aging in Europe (SHARE) | Hungary | 2021 | 112 | 50–95 | Panel | P, B, K, H, F | BMI; Smoking; Income |
| 659 | Study of Health and Aging in Europe (SHARE) | Hungary | 2022 | 828 | 21–98 | Panel | P, B, K, H, F | BMI; Smoking; Income |
| 660 | Study of Health and Aging in Europe (SHARE) | Israel | 2019 | 637 | 44–100 | Panel | P, B, K, H, F | BMI; Smoking; Income |
| 661 | Study of Health and Aging in Europe (SHARE) | Israel | 2020 | 763 | 46–100 | Panel | P, B, K, H, F | BMI; Smoking; Income |
| 662 | Study of Health and Aging in Europe (SHARE) | Israel | 2022 | 160 | 46–95 | Panel | P, B, K, H, F | BMI; Smoking; Income |
| 663 | Study of Health and Aging in Europe (SHARE) | Italy | 2019 | 798 | 40–100 | Panel | P, B, K, H, F | BMI; Smoking; Income |
| 664 | Study of Health and Aging in Europe (SHARE) | Italy | 2020 | 1366 | 46–100 | Panel | P, B, K, H, F | BMI; Smoking; Income |
| 665 | Study of Health and Aging in Europe (SHARE) | Italy | 2021 | 114 | 49–92 | Panel | P, B, K, H, F | BMI; Smoking; Income |
| 666 | Study of Health and Aging in Europe (SHARE) | Italy | 2022 | 1671 | 43–98 | Panel | P, B, K, H, F | BMI; Smoking; Income |
| 667 | Study of Health and Aging in Europe (SHARE) | Latvia | 2019 | 433 | 39–100 | Panel | P, B, K, H, F | BMI; Smoking; Income |
| 668 | Study of Health and Aging in Europe (SHARE) | Latvia | 2020 | 785 | 34–99 | Panel | P, B, K, H, F | BMI; Smoking; Income |
| 669 | Study of Health and Aging in Europe (SHARE) | Latvia | 2022 | 718 | 33–92 | Panel | P, B, K, H, F | BMI; Smoking; Income |
| 670 | Study of Health and Aging in Europe (SHARE) | Lithuania | 2019 | 812 | 45–94 | Panel | P, B, K, H, F | BMI; Smoking; Income |
| 671 | Study of Health and Aging in Europe (SHARE) | Lithuania | 2020 | 623 | 48–96 | Panel | P, B, K, H, F | BMI; Smoking; Income |
| 672 | Study of Health and Aging in Europe (SHARE) | Lithuania | 2021 | 111 | 45–93 | Panel | P, B, K, H, F | BMI; Smoking; Income |
| 673 | Study of Health and Aging in Europe (SHARE) | Lithuania | 2022 | 109 | 39–93 | Panel | P, B, K, H, F | BMI; Smoking; Income |
| 674 | Study of Health and Aging in Europe (SHARE) | Luxembourg | 2019 | 474 | 45–100 | Panel | P, B, K, H, F | BMI; Smoking; Income |
| 675 | Study of Health and Aging in Europe (SHARE) | Luxembourg | 2020 | 474 | 38–97 | Panel | P, B, K, H, F | BMI; Smoking; Income |

| | | | | | | | | |
|---|---|---|---|---|---|---|---|---|
| 676 | Study of Health and Aging in Europe (SHARE) | Luxembourg | 2021 | 7 | 61–79 | Panel | P, B, K, H, F | BMI; Smoking; Income |
| 677 | Study of Health and Aging in Europe (SHARE) | Luxembourg | 2022 | 147 | 58–93 | Panel | P, B, K, H, F | BMI; Smoking; Income |
| 678 | Study of Health and Aging in Europe (SHARE) | Malta | 2019 | 408 | 48–95 | Panel | P, B, K, H, F | BMI; Smoking; Income |
| 679 | Study of Health and Aging in Europe (SHARE) | Malta | 2020 | 396 | 48–97 | Panel | P, B, K, H, F | BMI; Smoking; Income |
| 680 | Study of Health and Aging in Europe (SHARE) | Malta | 2021 | 22 | 56–90 | Panel | P, B, K, H, F | BMI; Smoking; Income |
| 681 | Study of Health and Aging in Europe (SHARE) | Malta | 2022 | 217 | 52–95 | Panel | P, B, K, H, F | BMI; Smoking; Income |
| 682 | Study of Health and Aging in Europe (SHARE) | Netherlands | 2019 | 1035 | 49–96 | Panel | P, B, K, H, F | BMI; Smoking; Income |
| 683 | Study of Health and Aging in Europe (SHARE) | Netherlands | 2020 | 898 | 40–97 | Panel | P, B, K, H, F | BMI; Smoking; Income |
| 684 | Study of Health and Aging in Europe (SHARE) | Netherlands | 2021 | 189 | 55–91 | Panel | P, B, K, H, F | BMI; Smoking; Income |
| 685 | Study of Health and Aging in Europe (SHARE) | Netherlands | 2022 | 480 | 51–99 | Panel | P, B, K, H, F | BMI; Smoking; Income |
| 686 | Study of Health and Aging in Europe (SHARE) | Poland | 2019 | 783 | 45–98 | Panel | P, B, K, H, F | BMI; Smoking; Income |
| 687 | Study of Health and Aging in Europe (SHARE) | Poland | 2020 | 2064 | 34–100 | Panel | P, B, K, H, F | BMI; Smoking; Income |
| 688 | Study of Health and Aging in Europe (SHARE) | Poland | 2021 | 11 | 57–80 | Panel | P, B, K, H, F | BMI; Smoking; Income |
| 689 | Study of Health and Aging in Europe (SHARE) | Poland | 2022 | 2558 | 35–98 | Panel | P, B, K, H, F | BMI; Smoking; Income |
| 690 | Study of Health and Aging in Europe (SHARE) | Portugal | 2022 | 1632 | 40–99 | Panel | P, B, K, H, F | BMI; Smoking; Income |
| 691 | Study of Health and Aging in Europe (SHARE) | Romania | 2019 | 210 | 42–86 | Panel | P, B, K, H, F | BMI; Smoking; Income |
| 692 | Study of Health and Aging in Europe (SHARE) | Romania | 2020 | 1069 | 40–94 | Panel | P, B, K, H, F | BMI; Smoking; Income |
| 693 | Study of Health and Aging in Europe (SHARE) | Romania | 2021 | 42 | 52–93 | Panel | P, B, K, H, F | BMI; Smoking; Income |

| | | | | | | | | |
|---|---|---|---|---|---|---|---|---|
| 694 | Study of Health and Aging in Europe (SHARE) | Romania | 2022 | 361 | 50–100 | Panel | P, B, K, H, F | BMI; Smoking; Income |
| 695 | Study of Health and Aging in Europe (SHARE) | Slovakia | 2019 | 425 | 31–100 | Panel | P, B, K, H, F | BMI; Smoking; Income |
| 696 | Study of Health and Aging in Europe (SHARE) | Slovakia | 2020 | 572 | 44–92 | Panel | P, B, K, H, F | BMI; Smoking; Income |
| 697 | Study of Health and Aging in Europe (SHARE) | Slovakia | 2021 | 76 | 49–86 | Panel | P, B, K, H, F | BMI; Smoking; Income |
| 698 | Study of Health and Aging in Europe (SHARE) | Slovakia | 2022 | 79 | 53–80 | Panel | P, B, K, H, F | BMI; Smoking; Income |
| 699 | Study of Health and Aging in Europe (SHARE) | Slovenia | 2019 | 886 | 44–100 | Panel | P, B, K, H, F | BMI; Smoking; Income |
| 700 | Study of Health and Aging in Europe (SHARE) | Slovenia | 2020 | 2150 | 40–100 | Panel | P, B, K, H, F | BMI; Smoking; Income |
| 701 | Study of Health and Aging in Europe (SHARE) | Slovenia | 2021 | 243 | 54–99 | Panel | P, B, K, H, F | BMI; Smoking; Income |
| 702 | Study of Health and Aging in Europe (SHARE) | Slovenia | 2022 | 1667 | 38–100 | Panel | P, B, K, H, F | BMI; Smoking; Income |
| 703 | Study of Health and Aging in Europe (SHARE) | Spain | 2019 | 1094 | 51–99 | Panel | P, B, K, H, F | BMI; Smoking; Income |
| 704 | Study of Health and Aging in Europe (SHARE) | Spain | 2020 | 1017 | 37–100 | Panel | P, B, K, H, F | BMI; Smoking; Income |
| 705 | Study of Health and Aging in Europe (SHARE) | Spain | 2021 | 318 | 30–97 | Panel | P, B, K, H, F | BMI; Smoking; Income |
| 706 | Study of Health and Aging in Europe (SHARE) | Spain | 2022 | 814 | 40–97 | Panel | P, B, K, H, F | BMI; Smoking; Income |
| 707 | Study of Health and Aging in Europe (SHARE) | Sweden | 2019 | 1754 | 37–99 | Panel | P, B, K, H, F | BMI; Smoking; Income |
| 708 | Study of Health and Aging in Europe (SHARE) | Sweden | 2020 | 778 | 38–99 | Panel | P, B, K, H, F | BMI; Smoking; Income |
| 709 | Study of Health and Aging in Europe (SHARE) | Sweden | 2021 | 95 | 55–93 | Panel | P, B, K, H, F | BMI; Smoking; Income |
| 710 | Study of Health and Aging in Europe (SHARE) | Sweden | 2022 | 383 | 47–100 | Panel | P, B, K, H, F | BMI; Smoking; Income |
| 711 | Study of Health and Aging in Europe (SHARE) | Switzerland | 2019 | 1353 | 44–98 | Panel | P, B, K, H, F | BMI; Smoking; Income |

| | | | | | | | | |
|---|---|---|---|---|---|---|---|---|
| 712 | Study of Health and Aging in Europe (SHARE) | Switzerland | 2020 | 735 | 33–100 | Panel | P, B, K, H, F | BMI; Smoking; Income |
| 713 | Study of Health and Aging in Europe (SHARE) | Switzerland | 2021 | 40 | 60–90 | Panel | P, B, K, H, F | BMI; Smoking; Income |
| 714 | Study of Health and Aging in Europe (SHARE) | Switzerland | 2022 | 216 | 51–94 | Panel | P, B, K, H, F | BMI; Smoking; Income |
| 715 | Study on Nutrition and Cardiovascular Risk in Spain (ENRICA) | Spain | 2010 | 2451 | 62–97 | Household survey | P, B | |
| 716 | Survey on Aging in Latin America and Caribbean (SABE) | Argentina | 1999 | 1041 | 60–94 | Household survey | P, Ha, C, B | Smoking |
| 717 | Survey on Aging in Latin America and Caribbean (SABE) | Barbados | 1999 | 1508 | 60–97 | Household survey | P, Ha, C, B | BMI; Smoking |
| 718 | Survey on Aging in Latin America and Caribbean (SABE) | Brazil | 1999 | 2143 | 60–100 | Household survey | P, Ha, C, B | BMI; Smoking |
| 719 | Survey on Aging in Latin America and Caribbean (SABE) | Chile | 1999 | 1300 | 60–99 | Household survey | P, Ha, C, B | BMI; Smoking |
| 720 | Survey on Aging in Latin America and Caribbean (SABE) | Cuba | 1999 | 1905 | 60–100 | Household survey | P, Ha, C, B | Smoking |
| 721 | Survey on Aging in Latin America and Caribbean (SABE) | Mexico | 1999 | 1870 | 50–98 | Household survey | P, Ha, C, B | BMI; Smoking |
| 722 | Survey on Aging in Latin America and Caribbean (SABE) | Uruguay | 1999 | 1449 | 60–97 | Household survey | P, Ha, C, B | BMI; Smoking |
| 723 | Survey on Social, Health and Overall Wellbeing of Older People (SSHW) | Kenya | 2008 | 2585 | 44–100 | Household survey | P, B | |
| 724 | Swedish Health Assets Project 2008 (SHA) | Sweden | 2008 | 7835 | 19–64 | Household survey | P, Ha, N/S, C, S/A, B | BMI; Income |
| 725 | Swiss Health Panel (SHP) | Switzerland | 2000 | 7074 | 13–92 | Panel | Ha, B | Income |
| 726 | Swiss Health Panel (SHP) | Switzerland | 2001 | 659 | 13–86 | Panel | Ha, B | Income |
| 727 | Swiss Health Panel (SHP) | Switzerland | 2002 | 252 | 13–85 | Panel | Ha, B | Income |
| 728 | Swiss Health Panel (SHP) | Switzerland | 2003 | 211 | 13–79 | Panel | Ha, B | Income |
| 729 | Swiss Health Panel (SHP) | Switzerland | 2004 | 3879 | 13–94 | Panel | Ha, B | BMI; Income |
| 730 | Swiss Health Panel (SHP) | Switzerland | 2005 | 381 | 13–85 | Panel | Ha, B | BMI; Income |
| 731 | Swiss Health Panel (SHP) | Switzerland | 2006 | 413 | 14–84 | Panel | Ha, B | BMI; Income |
| 732 | Swiss Health Panel (SHP) | Switzerland | 2007 | 526 | 14–86 | Panel | Ha, B | BMI; Income |
| 733 | Swiss Health Panel (SHP) | Switzerland | 2008 | 398 | 14–84 | Panel | Ha, B | BMI; Income |
| 734 | Swiss Health Panel (SHP) | Switzerland | 2009 | 365 | 14–86 | Panel | Ha, B | BMI; Income |
| 735 | Swiss Health Panel (SHP) | Switzerland | 2010 | 377 | 14–92 | Panel | Ha, B | BMI; Smoking; Income |
| 736 | Swiss Health Panel (SHP) | Switzerland | 2011 | 284 | 14–82 | Panel | Ha, B | BMI; Smoking; Income |
| 737 | Swiss Health Panel (SHP) | Switzerland | 2012 | 274 | 14–82 | Panel | Ha, B | BMI; Income |
| 738 | Swiss Health Panel (SHP) | Switzerland | 2013 | 242 | 14–78 | Panel | Ha, B | BMI; Income |
| 739 | Swiss Health Panel (SHP) | Switzerland | 2014 | 5451 | 14–96 | Panel | Ha, B | BMI; Smoking; Income |

| | | | | | | | | |
|---|---|---|---|---|---|---|---|---|
| 740 | Swiss Health Panel (SHP) | Switzerland | 2015 | 479 | 14–95 | Panel | Ha, B | BMI; Smoking; Income |
| 741 | Swiss Health Panel (SHP) | Switzerland | 2016 | 415 | 13–94 | Panel | Ha, B | BMI; Smoking; Income |
| 742 | Swiss Health Panel (SHP) | Switzerland | 2017 | 333 | 14–88 | Panel | Ha, B | BMI; Smoking; Income |
| 743 | Swiss Health Panel (SHP) | Switzerland | 2018 | 326 | 14–87 | Panel | Ha, B | BMI; Smoking; Income |
| 744 | Swiss Health Panel (SHP) | Switzerland | 2019 | 289 | 14–88 | Panel | Ha, B | BMI; Smoking; Income |
| 745 | Swiss Health Panel (SHP) | Switzerland | 2020 | 7712 | 14–100 | Panel | Ha, B | BMI; Smoking; Income |
| 746 | Swiss Health Panel (SHP) | Switzerland | 2021 | 586 | 14–92 | Panel | Ha, B | BMI; Smoking; Income |
| 747 | Swiss Health Panel (SHP) | Switzerland | 2022 | 302 | 14–92 | Panel | Ha, B | BMI; Smoking; Income |
| 748 | Swiss Health Panel (SHP) | Switzerland | 2023 | 327 | 14–81 | Panel | Ha, B | BMI; Smoking; Income |
| 749 | Taiwan MJ Biobank (MJH) | Taiwan | 1996 | 58985 | 5–96 | Biobank/Repeat cross-section | S/A, B | Smoking |
| 750 | Taiwan MJ Biobank (MJH) | Taiwan | 1997 | 74029 | 5–97 | Biobank/Repeat cross-section | S/A, B | Smoking |
| 751 | Taiwan MJ Biobank (MJH) | Taiwan | 1998 | 73397 | 5–98 | Biobank/Repeat cross-section | S/A, B | Smoking |
| 752 | Taiwan MJ Biobank (MJH) | Taiwan | 1999 | 68439 | 5–99 | Biobank/Repeat cross-section | B | Smoking |
| 753 | Taiwan MJ Biobank (MJH) | Taiwan | 2000 | 73149 | 5–100 | Biobank/Repeat cross-section | B | Smoking |
| 754 | Taiwan MJ Biobank (MJH) | Taiwan | 2001 | 66810 | 5–98 | Biobank/Repeat cross-section | B | Smoking |
| 755 | Taiwan MJ Biobank (MJH) | Taiwan | 2002 | 68353 | 5–100 | Biobank/Repeat cross-section | B | Smoking |
| 756 | Taiwan MJ Biobank (MJH) | Taiwan | 2003 | 231 | 6–86 | Biobank/Repeat cross-section | B | Smoking |
| 757 | Taiwan MJ Biobank (MJH) | Taiwan | 2014 | 28731 | 7–95 | Biobank/Repeat cross-section | P | Smoking |
| 758 | Taiwan MJ Biobank (MJH) | Taiwan | 2015 | 31167 | 6–96 | Biobank/Repeat cross-section | P | Smoking |
| 759 | Taiwan MJ Biobank (MJH) | Taiwan | 2016 | 25817 | 8–95 | Biobank/Repeat cross-section | P | Smoking |
| 760 | Taiwan MJ Biobank (MJH) | Taiwan | 2017 | 26952 | 7–92 | Biobank/Repeat cross-section | P, N/S, B, K, H, E | Smoking |
| 761 | Taiwan MJ Biobank (MJH) | Taiwan | 2018 | 23009 | 8–93 | Biobank/Repeat cross-section | P, N/S, B, K, H, E | Smoking |
| 762 | Taiwan MJ Biobank (MJH) | Taiwan | 2019 | 22646 | 9–98 | Biobank/Repeat cross-section | P, N/S, B, K, H, E | Smoking |

| | | | | | | | | |
|---|---|---|---|---|---|---|---|---|
| 763 | Taiwan MJ Biobank (MJH) | Taiwan | 2020 | 18553 | 6–95 | Biobank/Repeat cross-section | P, N/S, B, K, H, E | Smoking |
| 764 | Taiwan MJ Biobank (MJH) | Taiwan | 2021 | 13414 | 7–95 | Biobank/Repeat cross-section | P, N/S, B, K, H, E | Smoking |
| 765 | Taiwan MJ Biobank (MJH) | Taiwan | 2022 | 13316 | 9–92 | Biobank/Repeat cross-section | P, N/S, B, K, H, E | Smoking |
| 766 | Thai Students Survey | Thailand | 2009 | 2750 | 9–19 | School-based survey | P, Ha, N/S, B, K, H, Hnd, Ft, E | BMI |
| 767 | The Filipino American Community Epidemiological Study (FACES) | United States | 1996 | 2297 | 7–65 | Cross-section | Ha, C, B | BMI |
| 768 | The Japan Society and New Tobacco Internet Survey (JASTIS) | Japan | 2025 | 28000 | 15–84 | Panel | P, Ha, N/S, C, S/A, B, H | BMI; Smoking; Income |
| 769 | The Longitudinal Study of Ageing and Health in the Philippines (LSAHP) | Philippines | 2018 | 4411 | 65–85 | Household survey | P, Ha, N/S, B, K, H, Ft | Smoking; Income |
| 770 | The National Longitudinal Study of Adolescent to Adult Health (ADD Health) | United States | 1995 | 6496 | 12–21 | Panel | P, Ha, C, S/A | BMI; Smoking; Income |
| 771 | The Netherlands Study of Depression and Anxiety (NESDA)[c] | Netherlands | 2004 | 652 | 18–65 | Cohort | P, Ha, C, S/A, B, F | BMI; Smoking |
| 772 | The Trøndelag Health Study (HUNT Study) | Norway | 1996 | 28357 | 19–100 | Cohort | P, N/S, C, B, K, H, Hnd, Ft, E | BMI; Smoking |
| 773 | The Trøndelag Health Study (HUNT Study) | Norway | 2007 | 25275 | 13–97 | Cohort | P, Ha, N/S, C, S/A, B, K, H, Hnd, Ft, E | BMI; Smoking |
| 774 | The Trøndelag Health Study (HUNT Study) | Norway | 2018 | 48965 | 13–100 | Cohort | P, Ha, N/S, C, S/A, B, K, H, F, Hnd, Ft, E | BMI; Smoking; Income |
| 775 | UK Biobank (UKB) | United Kingdom | 2008 | 453392 | 37–74 | Cohort | P, Ha, N/S, C, S/A, B, K, H, F | BMI; Smoking; Income |
| 776 | UK Biobank (UKB) | United Kingdom | 2016 | 48990 | 45–83 | Cohort | P, Ha, N/S, C, S/A, B, K, H, F | BMI; Smoking; Income |
| 777 | UK Biobank (UKB)[d] | United Kingdom | 2020 | 167225 | 49–84 | Cohort | P, Hnd, Ft | |
| 778 | Understanding America Survey (UAS) | United States | 2015 | 231 | 18–88 | Panel | P, Ha, B | BMI; Income |
| 779 | Understanding America Survey (UAS) | United States | 2016 | 560 | 18–91 | Panel | P, Ha, B | BMI; Income |
| 780 | Understanding America Survey (UAS) | United States | 2017 | 303 | 18–95 | Panel | P, Ha, B | BMI; Income |
| 781 | Understanding America Survey (UAS) | United States | 2018 | 504 | 18–89 | Panel | P, Ha, B | BMI; Income |
| 782 | Understanding America Survey (UAS) | United States | 2019 | 628 | 17–100 | Panel | P, Ha, B | BMI; Income |
| 783 | Understanding America Survey (UAS) | United States | 2020 | 851 | 18–100 | Panel | P, Ha, B | BMI; Income |
| 784 | Understanding America Survey (UAS) | United States | 2021 | 694 | 18–100 | Panel | P, Ha, B | BMI; Income |
| 785 | Understanding America Survey (UAS) | United States | 2022 | 882 | 18–100 | Panel | P, Ha, B | BMI; Income |
| 786 | Understanding America Survey (UAS) | United States | 2023 | 4126 | 18–100 | Panel | P, Ha, B | BMI; Income |
| 787 | Understanding America Survey (UAS) | United States | 2024 | 5760 | 18–97 | Panel | P, Ha, B | BMI; Income |
| 788 | Understanding America Survey (UAS) | United States | 2025 | 3303 | 18–90 | Panel | P, Ha, B | BMI; Income |
| 789 | WHEST (EpiGroup) | United Kingdom | 2015 | 2088 | 25–94 | Cohort | P, Ha, N/S, C, S/A, B, K, H, Hnd, Ft, E | Income |

| | | | | | | | | |
|---|---|---|---|---|---|---|---|---|
| 790 | WHO Multi-Country Survey (MCSS) | China | 2000 | 9434 | 18–92 | Household survey | P, C, B | BMI; Income |
| 791 | WHO Multi-Country Survey (MCSS) | Colombia | 2000 | 6015 | 17–99 | Household survey | P, C, B | BMI; Income |
| 792 | WHO Multi-Country Survey (MCSS) | Egypt | 2000 | 4471 | 18–95 | Household survey | P, C, B | BMI; Income |
| 793 | WHO Multi-Country Survey (MCSS) | Georgia | 2000 | 9837 | 18–100 | Household survey | P, C, B | BMI; Income |
| 794 | WHO Multi-Country Survey (MCSS) | India | 2000 | 5178 | 18–100 | Household survey | P, C, B | BMI; Income |
| 795 | WHO Multi-Country Survey (MCSS) | Indonesia | 2000 | 9927 | 17–100 | Household survey | P, C, B | BMI; Income |
| 796 | WHO Multi-Country Survey (MCSS) | Iran | 2000 | 9555 | 18–100 | Household survey | P, C, B | BMI; Income |
| 797 | WHO Multi-Country Survey (MCSS) | Lebanon | 2000 | 3233 | 18–100 | Household survey | P, C, B | BMI; Income |
| 798 | WHO Multi-Country Survey (MCSS) | Mexico | 2000 | 4800 | 18–96 | Household survey | P, C, B | BMI; Income |
| 799 | WHO Multi-Country Survey (MCSS) | Nigeria | 2000 | 5037 | 18–100 | Household survey | P, C, B | BMI; Income |
| 800 | WHO Multi-Country Survey (MCSS) | Singapore | 2000 | 6216 | 24–85 | Household survey | P, C, B | BMI |
| 801 | WHO Multi-Country Survey (MCSS) | Slovakia | 2000 | 1174 | 18–87 | Household survey | P, C, B | BMI; Income |
| 802 | WHO Multi-Country Survey (MCSS) | Syria | 2000 | 8603 | 18–100 | Household survey | P, C, B | BMI; Income |
| 803 | WHO Multi-Country Survey (MCSS) | Turkiye | 2000 | 5135 | 18–85 | Household survey | P, C, B | BMI; Income |
| 804 | WHO STEPS Nepal | Nepal | 2019 | 5593 | 15–69 | Household survey | P, Ha, B | BMI; Smoking |
| 805 | WHO Study on Global Aging and Adult Health (SAGE) | Bangladesh | 2007 | 4004 | 50–100 | Household survey | P | |
| 806 | WHO Study on Global Aging and Adult Health (SAGE) | China | 2007 | 14607 | 18–99 | Household survey | P, Ha, S/A, B | BMI; Smoking; Income |
| 807 | WHO Study on Global Aging and Adult Health (SAGE) | Ghana | 2007 | 9398 | 18–100 | Household survey | P, Ha, S/A, B | BMI; Smoking; Income |
| 808 | WHO Study on Global Aging and Adult Health (SAGE) | India | 2007 | 11227 | 18–100 | Household survey | P, Ha, S/A, B | BMI; Smoking; Income |
| 809 | WHO Study on Global Aging and Adult Health (SAGE) | Indonesia | 2007 | 11733 | 50–100 | Household survey | P | |
| 810 | WHO Study on Global Aging and Adult Health (SAGE) | Kenya | 2007 | 1991 | 50–100 | Household survey | P | |
| 811 | WHO Study on Global Aging and Adult Health (SAGE) | Mexico | 2007 | 2622 | 22–100 | Household survey | P, Ha, S/A, B | BMI; Smoking; Income |
| 812 | WHO Study on Global Aging and Adult Health (SAGE) | Russian Federation | 2007 | 4338 | 18–100 | Household survey | P, Ha, S/A, B | BMI; Smoking; Income |
| 813 | WHO Study on Global Aging and Adult Health (SAGE) | South Africa | 2007 | 4145 | 18–100 | Household survey | P, Ha, S/A, B | BMI; Smoking; Income |
| 814 | WHO Study on Global Aging and Adult Health (SAGE) | Tanzania | 2007 | 5010 | 50–97 | Household survey | P | |
| 815 | WHO Study on Global Aging and Adult Health (SAGE) | Vietnam | 2007 | 8515 | 50–99 | Household survey | P | |
| 816 | WHO World Health Survey (WHS) | Austria | 2003 | 1051 | 18–92 | Cross-section | P, B | BMI |
| 817 | WHO World Health Survey (WHS) | Bangladesh | 2003 | 5544 | 18–100 | Cross-section | P, B | BMI |
| 818 | WHO World Health Survey (WHS) | Belgium | 2003 | 1004 | 17–98 | Cross-section | P, B | BMI |

| | | | | | | | | |
|---|---|---|---|---|---|---|---|---|
| 819 | WHO World Health Survey (WHS) | Bosnia and Herzegovina | 2003 | 1028 | 18–84 | Cross-section | P, B | BMI |
| 820 | WHO World Health Survey (WHS) | Brazil | 2003 | 5000 | 18–95 | Cross-section | P, B | BMI |
| 821 | WHO World Health Survey (WHS) | Burkina Faso | 2003 | 4821 | 18–100 | Cross-section | P, B | BMI |
| 822 | WHO World Health Survey (WHS) | Chad | 2003 | 4626 | 17–100 | Cross-section | P, B | BMI |
| 823 | WHO World Health Survey (WHS) | China | 2003 | 3993 | 18–99 | Cross-section | P, B | BMI |
| 824 | WHO World Health Survey (WHS) | Comoros | 2003 | 1759 | 18–90 | Cross-section | P, B | BMI |
| 825 | WHO World Health Survey (WHS) | Congo | 2003 | 2246 | 17–85 | Cross-section | P, B | BMI |
| 826 | WHO World Health Survey (WHS) | Croatia | 2003 | 989 | 18–93 | Cross-section | P, B | BMI |
| 827 | WHO World Health Survey (WHS) | Czech Republic | 2003 | 929 | 18–93 | Cross-section | P, B | BMI |
| 828 | WHO World Health Survey (WHS) | Côte d'Ivoire | 2003 | 3147 | 18–95 | Cross-section | P, B | BMI |
| 829 | WHO World Health Survey (WHS) | Denmark | 2003 | 1003 | 17–92 | Cross-section | P, B | BMI |
| 830 | WHO World Health Survey (WHS) | Dominican Republic | 2003 | 4531 | 18–100 | Cross-section | P, B | |
| 831 | WHO World Health Survey (WHS) | Ecuador | 2003 | 4622 | 17–100 | Cross-section | P, B | BMI |
| 832 | WHO World Health Survey (WHS) | Estonia | 2003 | 1011 | 17–92 | Cross-section | P, B | BMI |
| 833 | WHO World Health Survey (WHS) | Eswatini | 2003 | 2059 | 17–98 | Cross-section | P, B | BMI |
| 834 | WHO World Health Survey (WHS) | Ethiopia | 2003 | 4929 | 18–100 | Cross-section | P, B | BMI |
| 835 | WHO World Health Survey (WHS) | Finland | 2003 | 1013 | 18–93 | Cross-section | P, B | BMI |
| 836 | WHO World Health Survey (WHS) | France | 2003 | 1007 | 18–92 | Cross-section | P, B | BMI |
| 837 | WHO World Health Survey (WHS) | Georgia | 2003 | 2755 | 18–100 | Cross-section | P, B | BMI |
| 838 | WHO World Health Survey (WHS) | Germany | 2003 | 1258 | 18–92 | Cross-section | P, B | BMI |
| 839 | WHO World Health Survey (WHS) | Ghana | 2003 | 3923 | 18–100 | Cross-section | P, B | BMI |
| 840 | WHO World Health Survey (WHS) | Greece | 2003 | 1000 | 18–95 | Cross-section | P, B | BMI |
| 841 | WHO World Health Survey (WHS) | Guatemala | 2003 | 4747 | 18–99 | Cross-section | P, B | BMI |
| 842 | WHO World Health Survey (WHS) | Hungary | 2003 | 1419 | 18–95 | Cross-section | P, B | BMI |
| 843 | WHO World Health Survey (WHS) | India | 2003 | 9870 | 17–100 | Cross-section | P, B | BMI |
| 844 | WHO World Health Survey (WHS) | Ireland | 2003 | 1011 | 18–92 | Cross-section | P, B | BMI |
| 845 | WHO World Health Survey (WHS) | Israel | 2003 | 1208 | 18–91 | Cross-section | P, B | BMI |
| 846 | WHO World Health Survey (WHS) | Italy | 2003 | 999 | 18–96 | Cross-section | P, B | BMI |
| 847 | WHO World Health Survey (WHS) | Kazakhstan | 2003 | 4495 | 18–89 | Cross-section | P, B | BMI |
| 848 | WHO World Health Survey (WHS) | Kenya | 2003 | 4406 | 18–100 | Cross-section | P, B | BMI |
| 849 | WHO World Health Survey (WHS) | Laos | 2003 | 4887 | 17–98 | Cross-section | P, B | BMI |
| 850 | WHO World Health Survey (WHS) | Latvia | 2003 | 855 | 18–89 | Cross-section | P, B | BMI |
| 851 | WHO World Health Survey (WHS) | Luxembourg | 2003 | 700 | 18–90 | Cross-section | P, B | BMI |

| | | | | | | | | |
|---|---|---|---|---|---|---|---|---|
| 852 | WHO World Health Survey (WHS) | Malawi | 2003 | 5227 | 18–96 | Cross-section | P, B | BMI |
| 853 | WHO World Health Survey (WHS) | Malaysia | 2003 | 6016 | 18–97 | Cross-section | P, B | BMI |
| 854 | WHO World Health Survey (WHS) | Mali | 2003 | 3649 | 17–98 | Cross-section | P, B | BMI |
| 855 | WHO World Health Survey (WHS) | Mauritania | 2003 | 3763 | 17–95 | Cross-section | P, B | BMI |
| 856 | WHO World Health Survey (WHS) | Mauritius | 2003 | 3888 | 18–93 | Cross-section | P, B | BMI |
| 857 | WHO World Health Survey (WHS) | Mexico | 2003 | 38745 | 18–100 | Cross-section | P, B | BMI |
| 858 | WHO World Health Survey (WHS) | Morocco | 2003 | 5000 | 18–98 | Cross-section | P, B | BMI |
| 859 | WHO World Health Survey (WHS) | Myanmar | 2003 | 5886 | 18–99 | Cross-section | P, B | |
| 860 | WHO World Health Survey (WHS) | Namibia | 2003 | 3988 | 18–100 | Cross-section | P, B | BMI |
| 861 | WHO World Health Survey (WHS) | Nepal | 2003 | 8686 | 18–100 | Cross-section | P, B | BMI |
| 862 | WHO World Health Survey (WHS) | Netherlands | 2003 | 1091 | 18–84 | Cross-section | P, B | BMI |
| 863 | WHO World Health Survey (WHS) | Norway | 2003 | 969 | 18–93 | Cross-section | P, B | BMI |
| 864 | WHO World Health Survey (WHS) | Pakistan | 2003 | 6370 | 18–100 | Cross-section | P, B | BMI |
| 865 | WHO World Health Survey (WHS) | Paraguay | 2003 | 5139 | 18–95 | Cross-section | P, B | BMI |
| 866 | WHO World Health Survey (WHS) | Philippines | 2003 | 10075 | 18–99 | Cross-section | P, B | BMI |
| 867 | WHO World Health Survey (WHS) | Portugal | 2003 | 1030 | 18–94 | Cross-section | P, B | BMI |
| 868 | WHO World Health Survey (WHS) | Russian Federation | 2003 | 4419 | 18–94 | Cross-section | P, B | BMI |
| 869 | WHO World Health Survey (WHS) | Senegal | 2003 | 2959 | 17–99 | Cross-section | P, B | BMI |
| 870 | WHO World Health Survey (WHS) | Slovakia | 2003 | 1788 | 18–98 | Cross-section | P, B | BMI |
| 871 | WHO World Health Survey (WHS) | Slovenia | 2003 | 584 | 18–91 | Cross-section | P, B | BMI |
| 872 | WHO World Health Survey (WHS) | South Africa | 2003 | 2342 | 18–100 | Cross-section | P, B | BMI |
| 873 | WHO World Health Survey (WHS) | Spain | 2003 | 6363 | 18–100 | Cross-section | P, B | BMI |
| 874 | WHO World Health Survey (WHS) | Sri Lanka | 2003 | 6728 | 18–100 | Cross-section | P, B | BMI |
| 875 | WHO World Health Survey (WHS) | Sweden | 2003 | 1000 | 18–92 | Cross-section | P, B | BMI |
| 876 | WHO World Health Survey (WHS) | Tunisia | 2003 | 5059 | 18–100 | Cross-section | P, B | BMI |
| 877 | WHO World Health Survey (WHS) | Turkiye | 2003 | 11204 | 18–99 | Cross-section | P | BMI |
| 878 | WHO World Health Survey (WHS) | Ukraine | 2003 | 2844 | 18–95 | Cross-section | P, B | BMI |
| 879 | WHO World Health Survey (WHS) | United Arab Emirates | 2003 | 1180 | 18–90 | Cross-section | P, B | BMI |
| 880 | WHO World Health Survey (WHS) | United Kingdom | 2003 | 1197 | 18–99 | Cross-section | P, B | BMI |
| 881 | WHO World Health Survey (WHS) | Uruguay | 2003 | 2979 | 18–93 | Cross-section | P, B | BMI |
| 882 | WHO World Health Survey (WHS) | Vietnam | 2003 | 3490 | 18–95 | Cross-section | P, B | BMI |
| 883 | WHO World Health Survey (WHS) | Zambia | 2003 | 3802 | 18–99 | Cross-section | P, B | BMI |
| 884 | WHO World Health Survey (WHS) | Zimbabwe | 2003 | 4051 | 17–100 | Cross-section | P, B | BMI |

| | | | | | | | | |
|---|---|---|---|---|---|---|---|---|
| 885 | WHO World Health Survey Plus (WHS+) | Cambodia | 2023 | 5320 | 18–88 | Cross-section | P, B | BMI |
| 886 | Wisconsin Longitudinal Study (WLS) | United States | 2010 | 4989 | 70–74 | Cohort | P, Ha, N/S, C, S/A, B, K, H, Hnd, Ft | BMI; Smoking; Income |
| 887 | Young Lives: Caregiver Survey | Ethiopia | 2002 | 1979 | 10–58 | Household survey | Ha | |
| 888 | Young Lives: Caregiver Survey | India | 2002 | 2000 | 12–54 | Household survey | Ha | |
| 889 | Young Lives: Caregiver Survey | Peru | 2002 | 2046 | 14–58 | Household survey | Ha | |
| 890 | Young Lives: Caregiver Survey | Vietnam | 2002 | 1906 | 15–59 | Household survey | Ha | |
| 891 | Young Lives: Child Survey | Ethiopia | 2016 | 11761 | 11–29 | School-based survey | Ha, S/A | |
| 892 | Young Lives: Child Survey | India | 2016 | 9720 | 12–18 | School-based survey | Ha, S/A | |
| 893 | Young Lives: Child Survey | Vietnam | 2016 | 8378 | 14–24 | School-based survey | Ha, S/A | |
| 894 | Yucatan Aging Study (YAS) | Mexico | 2015 | 6675 | 68–100 | Household survey | P | |

*Abbreviations: P, any pain; Ha, headache; N/S, neck or shoulder pain; C, chest pain; S/A, stomach or abdominal pain; B, back pain; K, knee pain; H, hip pain; Hnd, hand pain; Ft, foot pain; F, facial pain; E, elbow pain.*
*a. G21: At the Year 7 wave, pain items were parent-reported (mother or father responded on behalf of the child).*
*b. KiGGS: Pain items were parent-reported for children aged 3–10 years; self-reported for participants aged 11–17 years.*
*c. NESDA: Only the control sample (participants not selected on the basis of a depressive or anxiety disorder history) was included.*
*d. UK Biobank 2020: Refers to the UK Biobank "Experience of Pain" follow-up questionnaire; this instrument was used only to extract Hand, Foot, and Elbow pain.*
*e. LSMS: Headache item was parent-reported for adolescents aged 3–17 years.*
*f. Generation 21, New Zealand Health Survey, and Murakami Cohort contributed summary statistics rather than individual-participant data.*

Data were identified and accessed through a multi-stage process that combined literature searches, repository searches, and direct contact with investigators. We searched PubMed, Google Scholar, and Global Health Data Exchange (GHDx) using terms related to overall and site-specific pain, including "pain", "bodily pain", "back pain", "knee pain", "headache", "neck pain", "pain prevalence", and others, to identify population-based studies that met our inclusion criteria. In parallel, we reviewed major public microdata repositories, including Inter-university Consortium for Political and Social Research (ICPSR), the World Health Organization (WHO) Microdata Library, the World Bank Microdata Catalog, Harvard Dataverse, the UK Data Service, and national statistics portals. Publicly available datasets were downloaded directly from their respective repositories.

For studies that were not publicly available or for which pain variables were not included in public releases, we contacted study investigators to request access to individual participant data. We also submitted data-access applications to large population cohorts and biobanks such as Lifelines, UK Biobank, and CONSTANCES, which met inclusion criteria. Custodians of COPCORD and related rheumatology programs were contacted separately to request participation and data sharing.

All but three studies shared individual participant data. Generation 21, the New Zealand Health Survey, and the Murakami Cohort were governed by data-protection protocols that did not permit transfer of individual-level data outside the host institution. For these studies, local analysts generated age and sex stratified summary statistics for each pain site using a standardized protocol. All other studies provided full individual participant level data suitable for harmonization of pain variables and relevant covariates.

#### *2.2 Data inclusion criteria*

We included population-based surveys and cohort studies that recruited participants from the general population or from broadly defined community samples without restricting eligibility based on health status or specific clinical diagnoses. Studies were eligible if they included at least one self-report item assessing pain at one or more of the target bodily sites (head, face, neck or shoulder, chest, back, stomach or abdomen, hip, knee, hand or wrist, foot or ankle, or elbow), or a self-report measure of overall or average pain intensity recorded on a 0–10 numerical rating scale or visual analogue scale with 0 indicating no pain. Pain had to be assessed directly through participant self-report. Studies were excluded if they selectively sampled individuals on the basis of particular clinical conditions, did not assess any of the target pain sites or pain intensity, or relied on clinician ratings, proxy reports, or observational assessments rather than self-report. For a small number of paediatric cohorts, parent-reported pain items were accepted for young children below the age of reliable self-report (supplementary table 1, footnotes 1, 2, 5). For inclusion in the pooled analyses, individual-level data had to be available for at least age, sex, and one harmonizable pain variable.

### Section 3. Variable definitions, harmonization, and derived outcomes

#### *3.1 Pain outcome harmonization*

Across contributing studies, bodily pain was assessed using a range of instruments and question formats. Question wording varied substantially and included both time bound items, such as "*Over the past month, have you had any aches or pains in your body*?" or "*In the last 12 months, have you had pain that interfered with your life?*", and non-time specific items, such as "*Are you* *currently suffering from bodily pain?*" or "*Do you have pain in your [body site]?*". Recall periods ranged from very short windows, such as current pain or past day, through short to intermediate windows, such as past week or past month, to extended windows, such as past 3 months, 6 months, up to past 12 months. We did not restrict inclusion based on the exact question syntax or recall window. Instead, pain variables were harmonized by anatomical site and recall period was treated as a methodological covariate in the statistical models. For each pain item, we recorded the stated recall period and created a harmonized recall variable with three levels: current or short-term recall (current to past week), intermediate recall (past month), and extended recall (3 months or longer, including past 3, 6, or 12 months). This recall variable was included as a covariate in all models, and model-based prevalence estimates were standardized to a past month recall by setting the recall category to the intermediate (past month) level when generating predictions.

Past-month recall was selected as the standardisation reference based on measurement evidence and practical considerations. Validation studies comparing retrospective pain ratings against prospective momentary assessments have shown that recall accuracy degrades as the reporting period extends, but that correspondence between recalled and daily ratings remains acceptable at 28 days ($r \geq 0{\cdot}80$)[4,5]. Direct validation of the 1-month reference frame for the Graded Chronic Pain Scale demonstrated good psychometric properties and higher reporting stability (ICC = 0·89) compared with the 6-month version (ICC = 0·66), indicating a distinct recency advantage for the 30-day period[6]. Shorter recall periods (past week or less) risk underestimating the burden of symptoms that fluctuate day-to-day, while longer periods (3–12 months) introduce greater recall inflation and may conflate current with historical pain experience[7,8]. Past-month recall therefore provides a favourable balance between accuracy and representativeness. Additionally, past-month was the most common recall period across contributing data sources (used in 48% of studies compared to 25% for past week and 27% for past 3 months or longer), maximising the proportion of observations requiring no model-based adjustment.

We additionally included a study level indicator for pain case definition because 19 of 134 cohorts assessed chronic pain explicitly, typically requiring pain duration of at least 3 months, whereas other cohorts assessed pain without a chronicity requirement. Chronic explicit case definitions are expected to yield systematically lower prevalence than general pain questions[9] so this indicator was included as a fixed effect alongside recall window to adjust for systematic differences in outcome definition across cohorts. Prevalence estimates were then standardized to the general pain definition by fixing this indicator to the general pain level when generating predictions.

*For interpretability, the standardized outcome can be understood as pain, aching, or discomfort in a given anatomical region reported as experienced within the past month.*

We identified a core set of eleven anatomical sites that were sufficiently common across datasets and affected by pain across life to support pooled modelling: head, face, neck or shoulder, chest, back, stomach or abdomen, hip, knee, hand or wrist, foot or ankle, and elbow. Study specific pain items were mapped to these harmonized categories using study provided codebooks, original questionnaires where available, and, when necessary, consultation with study investigators. As an example, terms such as "oral", "jaw", and "orofacial" were grouped under facial pain, and items referring to "spine" or "lower back" were grouped as back pain. Where studies assessed neck and shoulder pain separately, these were combined into a single neck or shoulder category to maximize comparability with surveys that used a joint neck or shoulder item. A complete mapping from study specific item labels, question texts, and body map codes to the harmonized anatomical categories is provided in supplementary table 2.

**Supplementary Table 2: Harmonization definitions and inclusion criteria for anatomical pain sites**

| Site | Operational anatomical definition | Included source phrasings | Notes/edge rules |
|---|---|---|---|
| Head | Scalp/cranial regions: forehead, temples, crown, occiput | “headache”, “head pain/discomfort”, “pain in the head” | Exclude face or jaw items; exclude ear pain |
| Face | Orofacial regions: jaw, cheeks, mouth, peri-oral | “facial pain/discomfort”, “jaw pain”, “mouth pain”, “orofacial pain” | |
| Neck/ Shoulder | Cervical region and shoulder girdle: anterior/posterior neck, trapezius, acromion/clavicle, periscapular | “neck pain/discomfort”, “shoulder pain/discomfort”, “stiff neck”, “shoulder blade pain” | Map “upper back” here only when trapezius or shoulder blade is specified; otherwise count as back |
| Chest | Anterior thorax: sternum, ribs, upper front torso | “chest pain/discomfort”, “rib pain”, “thoracic pain/discomfort” | Exclude diagnostic specifications such as “angina”; include symptom wording only. Exclude breast pain and heartburn |
| Back | Posterior thorax and lumbar regions: upper, mid, lower back; spine; waist | “back pain/discomfort”, “back ache”, “lower back”, “lumbar”, “mid-back”, “upper back”, “spine”, “waist pain” | If “upper back” lacks trapezius/shoulder cues, treat as back (not neck/shoulder) |
| Stomach/ Abdomen | Anterior abdomen/stomach: upper or lower stomach or abdomen, belly | “abdominal pain/discomfort”, “stomach pain/discomfort”, “stomach ache”, “belly ache” | |
| Hip | Hip joint and peri-pelvic/inguinal: groin, lateral hip, buttocks/gluteal | “hip pain/discomfort”, “groin pain”, “buttocks pain”, “outer pelvis”, “hip stiffness”, “hip swelling” | |
| Knee | Knee joint: anterior/posterior, patella | “knee pain/discomfort”, “pain in the knee”, “knee joint”, “knee stiffness”, “knee swelling” | |
| Hand/Wrist | Distal upper limb: wrist, hand, palm, fingers, thumb | “hand pain/discomfort”, “wrist pain/discomfort”, “finger pain”, “thumb pain” | |
| Elbow | Elbow region | “elbow pain/discomfort”, “pain in elbow” | |
| Foot/Ankle | Distal lower limb: ankle, heel, foot, toes | “foot pain/discomfort”, “ankle pain/discomfort”, “heel pain”, “toe pain” | |
| Any Bodily Pain | Composite outcome representing pain present at any assessed location or globally | “general pain”, “bodily pain/discomfort”, “pain intensity > | Derived variable. Study inclusion restricted to those assessing ≥5 specific sites OR a general bodily pain item OR a general |

| | | | |
|---|---|---|---|
| | | 0”, or endorsement of ≥1 specific anatomical site | pain/intensity item. Participants coded as present if reporting pain on any eligible item. |
| Joint Pain | Composite phenotype representing pain in appendicular joints (shoulder, elbow, wrist/hand, hip, knee, ankle/foot) | “joint pain”, “aching joints”, “painful joints”, “stiff joints” | Derived variable: coded present if ≥1 joint site reported or if a study assessed general joint pain without anatomical specification. |

We derived a composite "any bodily pain" outcome for studies that assessed at least five anatomical sites, included a general pain item (defined as a single question assessing overall bodily pain without site specification; for example, "During the past month, have you had any bodily pain?"), or collected pain intensity ratings on a numerical scale. Participants were classified as having "any bodily pain" if they reported pain at one or more specific anatomical sites, endorsed the general pain item, or reported pain intensity greater than 0. Studies that assessed fewer than five anatomical sites and did not include a general pain item were excluded from this composite outcome.

The primary outcomes for site specific analyses were binary indicators of pain presence at each anatomical site. Because contributing cohorts used heterogeneous instruments, question wording, and response options, we followed established principles for retrospective harmonization and integrative data analysis by prespecifying a target construct and applying transparent, rule-based recoding to derive comparable variables across studies[10,11]. Most cohorts provided site specific pain outcomes that were already dichotomous (116 of 134), whereas a smaller subset used ordinal response options (18 of 134), requiring prespecified thresholding to derive binary indicators. For ordinal frequency items, categories reflecting pain occurring at least weekly (e.g., 'once a week', 'often', 'daily') were coded as present. For ordinal severity items, responses indicating more than minimal discomfort were coded as present; responses such as 'no pain', 'a little', or 'slight' were coded as absent. Items whose wording or response structure could not be mapped transparently to this binary definition were excluded from pooled analyses for the relevant site.

#### *3.2 Secondary pain outcomes*

Secondary outcomes captured both pain severity and the spatial extent of pain. Where available, average or general pain intensity rated on a 0–10 numerical rating scale or visual analogue scale with 0 indicating no pain was retained as a continuous variable. We restricted these analyses to items that explicitly asked about "average", "usual", or "overall" pain intensity, and excluded items that referred only to "worst pain", "least pain", or "pain at its most intense". Intensity items coding only coarse ordinal response categories such as mild, moderate, or severe were not included in the intensity analyses, as our focus was on numerical and analogue scales that are widely used and validated as core outcome measures in clinical pain research[12,13].

As with pain presence, pain intensity items differed in recall period. We recorded the stated recall window for each intensity question and applied the same harmonized recall structure used for site-specific prevalence. Recall was grouped into short-term (current or past week), intermediate (past month), and extended (past 3 months). This recall variable was included as a covariate in all intensity models so that differences in reported intensity attributable to recall length were partially adjusted for. When generating model-based intensity trajectories, we standardized predictions to a past week recall, aligning with recommended guidelines in clinical pain assessment[5,14].

Spatial extent of pain was quantified using generalized widespread pain criteria defined by the 2016 fibromyalgia diagnostic criteria and subsequent updates by Wolfe and colleagues[15,16]. The aim was not to diagnose fibromyalgia, but rather to characterize the point prevalence of highly burdensome, anatomically widespread pain in the general population using contemporary consensus criteria. For each participant, we assigned the collected pain sites to five body regions (left upper, right upper, left lower, right lower, and axial). Regions were coded as positive if any mapped site within that region was reported as painful; for example, a positive response for any of left hand, left elbow, left shoulder, or left arm would count as positive for the left upper region. Generalized widespread pain was then defined as pain present in at least four of the five bodily regions[15]. Only studies that collected sufficient laterality and axial site detail to construct all five regions were eligible for these analyses. In total, 28 of 134 studies met these criteria and were included. For models of generalized widespread pain, recall period was again included as a covariate, and predicted trajectories were standardized to a past week recall to match the recall horizon used in the underlying criteria[15]. Because many contributing studies did not collect the full symptom severity components (WPI/SSS), and chronicity (≥3 months) was not consistently assessed even among those with adequate site detail, we report generalised pain distribution rather than the full ACR fibromyalgia case definition.

#### *3.3 Socioeconomic and geographic variables*

*Total household income*

Total household income was used as a pragmatic indicator of socioeconomic position, capturing material resources and purchasing power at the household level, which are widely used dimensions of socioeconomic circumstances in population health research[17,18]. For children and adolescents, this variable reflects parent or guardian reported total household income.

Because income was collected using different currencies, scales, and reporting conventions across cohorts, we operationalized income as a within study relative rank rather than as an absolute monetary value. Where self-reported total household income was available, we grouped participants into within study income quintiles (lowest 20% through highest 20% among respondents with non-missing income) and used these ordered categories in modelling. Income quintiles should be interpreted as relative socioeconomic position within each cohort, not as an absolute measure of poverty across countries or survey years.

*World region classification*

Countries were assigned to world regions using the United Nations SDG Indicators regional groupings, which are based on the United Nations Statistics Division M49 geographic regions[19]. We used a slightly modified version of these SDG regional groupings which disaggregated the combined “Europe and Northern America” region into three groups: Northern America, Western Europe, and Eastern Europe, to improve interpretability and geographic resolution in high income settings. Each study observation was assigned a world region based on the country or territory in which the survey was conducted. The mapping of every country and territory in the pooled dataset to its assigned world region is provided in Supplementary Table 3.

**Supplementary Table 3: Classification of countries and territories by world region**

| Country | Global Region |
|---|---|
| Bangladesh | Central Asia |
| India | Central Asia |
| Pakistan | Central Asia |
| Kazakhstan | Central Asia |
| Sri Lanka | Central Asia |
| Nepal | Central Asia |
| Vietnam | East and SE Asia |
| Thailand | East and SE Asia |
| Taiwan | East and SE Asia |
| South Korea | East and SE Asia |
| Singapore | East and SE Asia |
| Philippines | East and SE Asia |
| Malaysia | East and SE Asia |
| Laos | East and SE Asia |
| Japan | East and SE Asia |
| Indonesia | East and SE Asia |
| China | East and SE Asia |
| Cambodia | East and SE Asia |
| Myanmar | East and SE Asia |
| Montenegro | Eastern Europe |
| North Macedonia | Eastern Europe |
| Poland | Eastern Europe |
| Romania | Eastern Europe |
| Slovenia | Eastern Europe |
| Serbia | Eastern Europe |
| Slovakia | Eastern Europe |
| Ukraine | Eastern Europe |
| Moldova | Eastern Europe |
| Russian Federation | Eastern Europe |
| Latvia | Eastern Europe |
| Lithuania | Eastern Europe |
| Kosovo | Eastern Europe |
| Albania | Eastern Europe |
| Armenia | Eastern Europe |
| Bosnia and Herzegovina | Eastern Europe |
| Bulgaria | Eastern Europe |
| Azerbaijan | Eastern Europe |

| | |
|---|---|
| Czech Republic | Eastern Europe |
| Estonia | Eastern Europe |
| Georgia | Eastern Europe |
| Hungary | Eastern Europe |
| Croatia | Eastern Europe |
| Mexico | Latin America |
| Uruguay | Latin America |
| Puerto Rico | Latin America |
| Peru | Latin America |
| Paraguay | Latin America |
| Venezuela | Latin America |
| Guatemala | Latin America |
| Colombia | Latin America |
| Dominican Republic | Latin America |
| Cuba | Latin America |
| Chile | Latin America |
| Brazil | Latin America |
| Barbados | Latin America |
| Argentina | Latin America |
| Ecuador | Latin America |
| Saudi Arabia | North Africa and West Asia |
| United Arab Emirates | North Africa and West Asia |
| Turkiye | North Africa and West Asia |
| Tunisia | North Africa and West Asia |
| Syria | North Africa and West Asia |
| Qatar | North Africa and West Asia |
| Iran | North Africa and West Asia |
| Libya | North Africa and West Asia |
| Lebanon | North Africa and West Asia |
| Jordan | North Africa and West Asia |
| Israel | North Africa and West Asia |
| Morocco | North Africa and West Asia |
| Egypt | North Africa and West Asia |
| Canada | Northern America |
| Greenland | Northern America |
| United States | Northern America |
| Tonga | Oceania |
| New Zealand | Oceania |
| Australia | Oceania |

| | |
|---|---|
| Kiribati | Oceania |
| Mali | Sub-saharan Africa |
| Zambia | Sub-saharan Africa |
| Tanzania | Sub-saharan Africa |
| South Africa | Sub-saharan Africa |
| Senegal | Sub-saharan Africa |
| Nigeria | Sub-saharan Africa |
| Namibia | Sub-saharan Africa |
| Zimbabwe | Sub-saharan Africa |
| Mauritania | Sub-saharan Africa |
| Malawi | Sub-saharan Africa |
| Mauritius | Sub-saharan Africa |
| Ghana | Sub-saharan Africa |
| Ethiopia | Sub-saharan Africa |
| Eswatini | Sub-saharan Africa |
| Democratic Republic of the Congo | Sub-saharan Africa |
| Cote d'Ivoire | Sub-saharan Africa |
| Congo | Sub-saharan Africa |
| Comoros | Sub-saharan Africa |
| Chad | Sub-saharan Africa |
| Burkina Faso | Sub-saharan Africa |
| Kenya | Sub-saharan Africa |
| Luxembourg | Western Europe |
| Malta | Western Europe |
| Netherlands | Western Europe |
| Sweden | Western Europe |
| Portugal | Western Europe |
| Spain | Western Europe |
| Italy | Western Europe |
| Norway | Western Europe |
| Ireland | Western Europe |
| Finland | Western Europe |
| Greece | Western Europe |
| Germany | Western Europe |
| France | Western Europe |
| Denmark | Western Europe |
| Cyprus | Western Europe |
| Belgium | Western Europe |
| Austria | Western Europe |

| | |
|---|---|
| Switzerland | Western Europe |
| Iceland | Western Europe |
| United Kingdom | Western Europe |

*Human Development Index (HDI)*

The Human Development Index (HDI) is a composite measure of human well-being developed by the United Nations Development Programme (UNDP). It combines three normalised dimensions: (1) health, measured by life expectancy at birth; (2) education, measured by mean years and expected years of schooling; and (3) standard of living, measured by gross national income (GNI) per capita. Each dimension is expressed on a scale from 0 to 1, and the HDI is calculated as the geometric mean of the three indices. Country level Human Development Index (HDI) values were obtained from the United Nations Development Programme (UNDP) Human Development Reports database (https://hdr.undp.org/data-center/human-development-index#/indicies/HDI), which provides annual HDI estimates by country. HDI values were linked to individual participants based on the country in which the study was conducted and the calendar year of data collection. When a study spanned multiple years, the HDI value corresponding to the midpoint year of data collection was used. If HDI values were not available for a specific country year, values were interpolated using adjacent years where possible. HDI was analyzed as a contextual, country level indicator of socioeconomic development and was not interpreted as an individual level socioeconomic measure.

### Section 4. Data processing and quality control

We applied a common data processing pipeline to all studies. Across all contributing studies, the raw pooled dataset contained reported ages ranging from approximately 3 to 117 years. A small number of observations fell outside biologically plausible limits (e.g., ages >110) and were judged to reflect data-entry or coding errors; these records were removed prior to harmonization. To ensure consistent representation of early-life ages across studies, ages <5 years were winsorized to 5 years. To stabilize estimates at the upper end of the distribution, ages >100 years were winsorized to 100 years. After these procedures, the analytic dataset comprised participants aged 5–100+ years.

Implausible values for BMI and underlying anthropometrics were identified using global thresholds and set to missing. For adults, BMI values below 10 kg/m² or above 80 kg/m² were treated as implausible. For pediatric participants, BMI was handled using age and sex specific BMI for age reference categories based on established WHO growth charts[20]. BMI was analyzed as categories (normal weight, overweight, obese), using standard adult cut points and pediatric percentile-based definitions[21].

The final pooled sample included 134 study programmes (defined as overarching survey or cohort frameworks with a consistent questionnaire structure and design) spanning 118 countries and territories. Within these study programmes, 894 country- or wave-specific datasets were identified (for example, individual survey years or country implementations), contributing approximately 6 million cross-sectional individual-level observations with information on pain at one or more body sites or a pain intensity rating. Data collection spanned calendar years 1990 to 2025.

### Section 5. Statistical methods

#### *5.1 Primary prevalence models*

We estimated age-specific prevalence trajectories using hierarchical generalized linear mixed-effects models (GLMM) with a binomial distribution and logit link function. This framework accounted for the hierarchical and partially crossed structure of the data, adjusting for methodological heterogeneity across sources. All analyses were performed in R (version 4.4.1). For an individual $i$ within study programme $p$ and world region $r$, the probability of reporting pain ($p_{ipr}$) was modeled as:

$$\text{logit}(p_{ipr}) = \beta_0 + f(\text{Age}_i, \text{Sex}_i) + \beta_{TW}\text{TimeWindow}_i + \beta_{CD}\text{CaseDefinition}_i + u_r + v_p$$

Non-linear age trajectories were modeled using natural cubic splines ($f$) with 3 degrees of freedom. An interaction term between the age spline and sex allowed the shape of the lifespan trajectory to differ freely between females and males. To harmonize diverse survey instruments, the model included fixed effects for the recall Time Window ($TW$; e.g., past week, past month, past 3-12 months) and Case Definition ($CD$; general pain vs. explicit chronic definition). To account for the complex clustering of the data, we specified a crossed random effects structure. This included random intercepts for world region ($u_r$) and study programme ($v_p$).

We derived globally comparable reference curves using estimated marginal means. Predicted probabilities were generated on a grid of ages from 5 to 100 years and standardized to a harmonized reference profile defined as "past month" recall and "general" pain definition. Random effects for world region and study were included during model fitting to account for clustering and heterogeneity. For reporting reference curves and contrasts, predictions were computed from fixed effects only, with random effects set to zero, corresponding to a reference region and study in the model. Disparities were summarized as age specific risk differences and risk ratios on the probability scale. Uncertainty for predicted probabilities and age-specific contrasts was quantified using Wald-based standard errors derived from the fixed-effects variance–covariance matrix of the fitted mixed-effects model, and we report pointwise 95% confidence intervals.

#### *5.2 Disparity metrics and dynamics*

We quantified group disparities at each age using the Risk Difference (RD) ($p_{\text{Group A}} - p_{\text{Group B}}$) and Risk Ratio (RR) ($p_{\text{Group A}}/p_{\text{Group B}}$). When reporting summary contrasts within pre-specified age bands (5–17, 18–35, 35–50, 50–65, 65–80, and $\geq$ 80 years), predicted probabilities were averaged across ages within the band before computing the RD and RR.

To characterize changes in prevalence across the life course, we computed the rate of change of the predicted prevalence curve using finite differences on a one-year grid ($[p_{a+1} - p_a]/1$ year). These rates are reported as percentage points per year to highlight phases of acceleration and deceleration.

#### *5.3 Exposure analyses (BMI, smoking, income), HDI, region*

To assess how pain trajectories are modified by sociodemographic and lifestyle factors, we extended the primary framework by interacting age with the exposure of interest. To ensure stable estimation of age-varying effects within smaller stratified subgroups, the age spline complexity was reduced to 2 degrees of freedom. For categorical risk factors (BMI category, smoking status, household income quintile), we estimated age-specific prevalence trajectories and age-adjusted prevalence estimates for each subgroup. For national development, we modeled the Human

Development Index (HDI) as a continuous variable interacting with the age spline. For presentation, predicted trajectories were generated at four representative HDI values (0·55, 0·70, 0·80, 0·90) aligned with United Nations thresholds for low, medium, high, and very high human development. We derived adjusted prevalence estimates, risk differences, and risk ratios using estimated marginal means. All estimates were standardized to a harmonized reference profile ("past month" recall, "general" pain definition) and computed from fixed effects to represent the global expectation.

#### *5.4 Population attributable fraction (PAF) analysis*

We estimated population attributable fractions (PAFs) to quantify the proportion of the pain burden associated with obesity, smoking, and low household income rank. We combined age- and sex-specific risk ratios (RRs) derived from the pooled cohort (Section 5.3) with country-, age-, and sex-specific exposure prevalence and population denominators from the Global Burden of Disease (GBD) study. Obesity prevalence was obtained from GBD 2021 estimates, merging child and adolescent (ages 5–24 years)[22] and adult (ages ≥ 25 years)[23] datasets to ensure continuity across the life course. Smoking prevalence was derived from the most recent GBD release providing granular age-sex resolution (GBD 2015)[24]. All external inputs were harmonised to 5-year age-by-sex strata and aligned to the analytic age range (ages ≥ 5 years, with the oldest group capped at ≥ 80 years). For obesity and smoking, PAFs were computed using the standard binary formulation:

$$PAF = \frac{P(RR - 1)}{1 + P(RR - 1)}$$

where $P$ denotes the country-, age-, and sex-specific exposure prevalence, and $RR$ denotes the corresponding age- and sex-specific risk ratio.

For income, socioeconomic position was operationalised as within-study household income quintiles (relative rank), with the quintile prevalence fixed at $P = 0.20$ under a relative-income framework. We estimated a partial polytomous PAF to quantify the pain burden specifically attributable to the lowest income quintile. This metric represents the proportion of total pain cases that would be eliminated if individuals in the bottom 20% of the population attained the health status of the top 20% (the reference group), while correctly accounting for the population-average risk implied by the full socioeconomic gradient. This was calculated as:

$$PAF_{lowest} = \frac{P_{lowest}(RR_{lowest} - 1)}{\sum_{i=1}^{5} P_i \, RR_i}$$

where $i$ indexes the income quintiles, $P_i = 0.20$ for all quintiles, and $RR_i$ is the risk ratio for the $i$-th quintile relative to the highest income quintile. Because income was operationalised as within-study relative rank, the prevalence of each quintile is fixed at 0.20 across all countries by definition; cross-national variation in income PAF therefore reflects differences in demographic structure interacting with age- and sex-specific risk ratios rather than differences in exposure prevalence.

To estimate the joint burden attributable to all three exposures simultaneously, we computed a combined PAF using the multiplicative independence formula:

$$PAF_{combined} = 1 - (1 - PAF_{obesity})(1 - PAF_{smoking})(1 - PAF_{income})$$

which accounts for co-occurrence of exposures. Country- and regional-level summary PAFs were obtained by summing exposure-attributable cases and total pain cases across age-sex strata and across the 11 anatomical pain sites, then computing the ratio. Regional and global estimates therefore represent case-weighted averages across the 11 site-specific models. Results are reported in supplementary p.81 (Section 8).

#### *5.5 Secondary outcome models (average pain intensity, high-intensity pain, generalized widespread pain)*

We analyzed average pain intensity (1–10 NRS; Extended Data Figure 6) and high-intensity pain (NRS $\geq 7$) using the hierarchical framework described above, adjusted for recall window and clustering within study and region. Pain intensity was modeled using a generalized linear mixed model with a Gamma distribution and log link function to account for the non-negative, right-skewed distribution of symptom ratings. High-intensity pain and generalized widespread pain were modeled using a binomial distribution with a logit link.

### Section 6. Sensitivity analyses

We conducted a series of sensitivity analyses to assess the robustness of our findings. These included: (1) a leave-one-study-out (LOSO) analysis to quantify single-study influence; (2) covariate adjustments for survey methodology (e.g., mode, setting); (3) data-exclusion scenarios based on recall period stringency; (4) alternative random-effects specifications (random slopes); (5) model selection for functional form (spline complexity); and (6) an imputation analysis to evaluate the impact of missing exposure variables (e.g., BMI, smoking status, and household income) on exposure–pain associations.

The estimated reference curves demonstrated high stability across these sensitivity checks. The exclusion of individual studies and adjustment for minor methodological covariates yielded negligible shifts in prevalence estimates (typically < 1.0 percentage points). While recall period was identified as a primary source of heterogeneity, our fixed-effect adjustment strategy produced trajectories highly concordant with stricter data-inclusion subsets. Although random-slope specifications suggested slightly altered trajectory shapes for headache and back pain specifically (shifting the peak age or flattening the late-life decline), the fundamental conclusions regarding the magnitude of sex disparities, the relative ranking of pain sites, and the ubiquitous burden of pain across the lifespan remained consistent across all valid sensitivity scenarios.

#### Section 6.1 Leave-One-Study-Out Sensitivity Analysis

To evaluate the robustness of the estimated reference curves, we performed a leave-one-study-out (LOSO) sensitivity analysis. We iteratively removed one study at a time from the dataset, refit the generalized linear mixed models, and recalculated age-specific predicted prevalence using the same protocol applied in the main analysis. The influence of each study was quantified as the Mean Absolute Difference (MAD) which is defined as the mean absolute difference in predicted prevalence between the LOSO model and the full model, averaged across the modeled age range and sex.

The analysis demonstrated high consistency across all pain sites (Supplementary Figure 1). The trajectories were robust to the exclusion of individual datasets: all studies produced an MAE of less than 2.5 percentage points, with a median deviation of approximately 0.06 percentage points.

As shown in the study influence matrix (Supplementary Figure 2), the datasets with the greatest influence were primarily those providing unique coverage of specific age groups or world regions. These included the WHO Study on global Ageing and adult health (SAGE), the WHO Multi-Country Survey Study (MCSS), and the Libyan Pain Survey, which provide critical data from non-Western populations. Additionally, cohorts focused on childhood and adolescence such as CATS (Childhood to Adult Transition Study), PSC (Pain in School Children), HBSC (Health Behavior in School Age Children), and ABCD (Adolescent Brain Cognitive Development Study), were among the most influential. This is expected, as these studies anchor the early-life portion of the lifespan trajectory where data is sparser compared to adulthood. However, even when excluding these key datasets, the magnitude of change remained small (mean MAE ≈ 1-2.5%) and did not alter the fundamental shape or relative ordering of the trajectories. This supports that the reported reference curves are not driven by the idiosyncratic features of any single study.

**Supplementary Figure 1: Stability of global trajectories in leave-one-study-out sensitivity analysis**

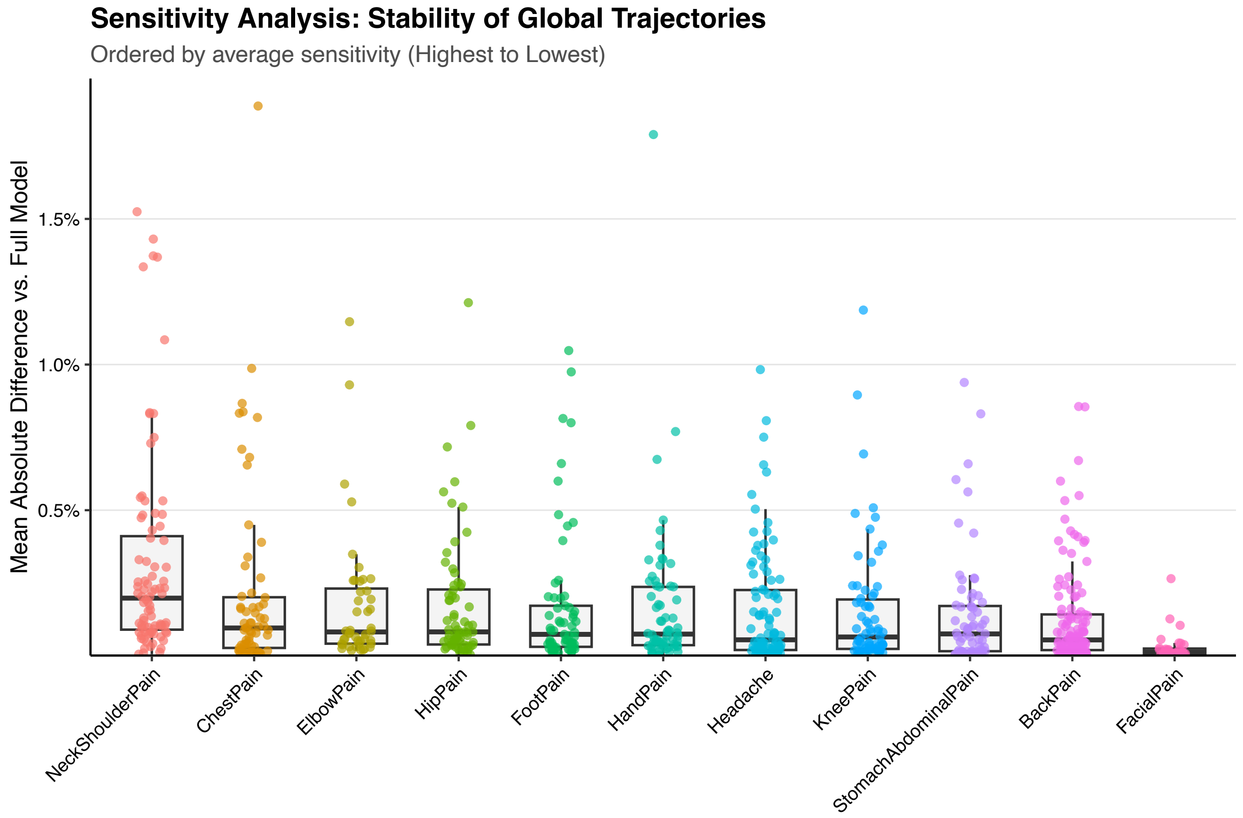

*Distribution of the Mean Absolute Difference (MAD) in predicted prevalence between the full model and leave-one-study-out models. Each point represents a single excluded study. Boxplots indicate the median and interquartile range.*

### Supplementary Figure 2: Matrix of individual study influence on prevalence estimates

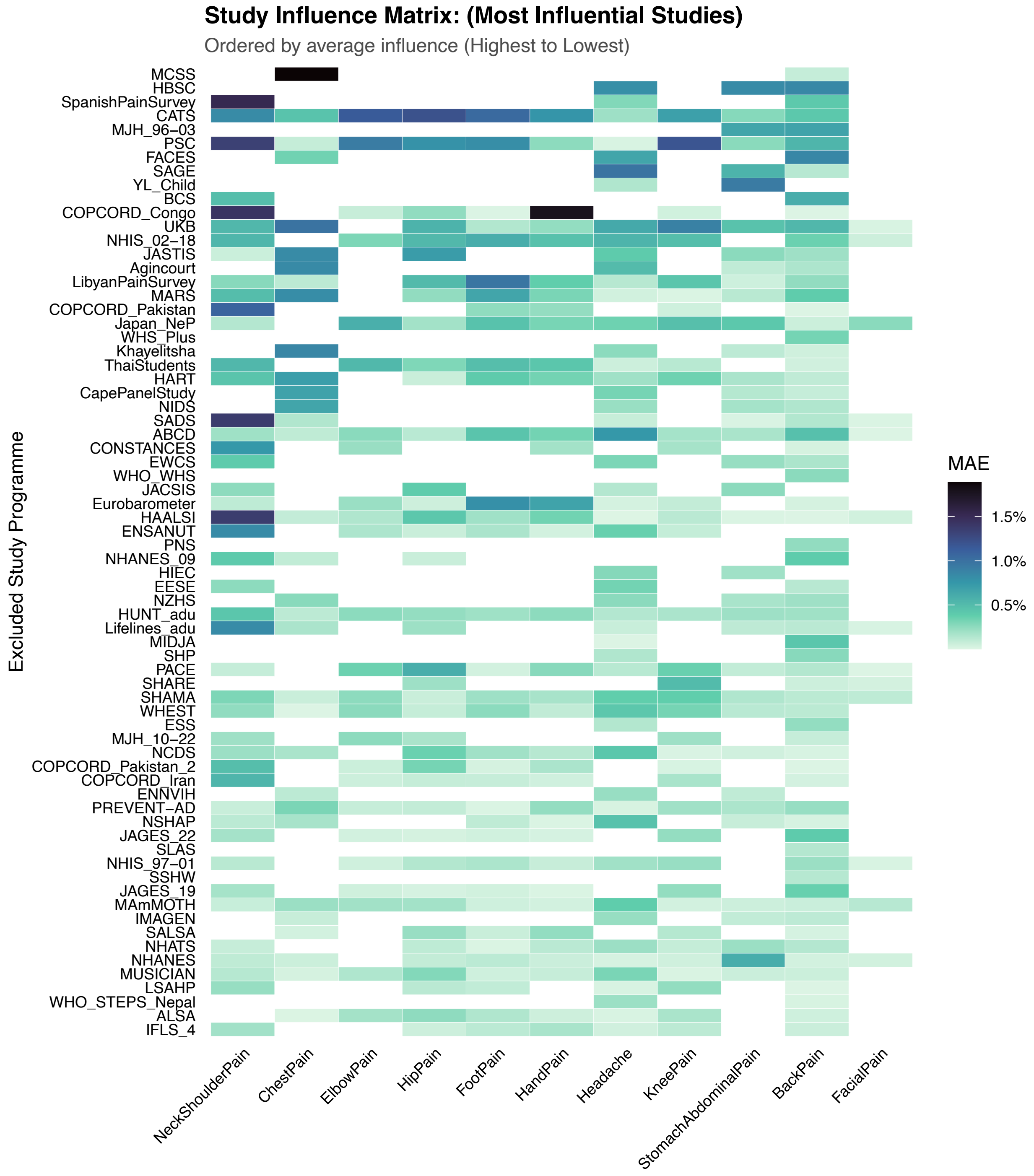

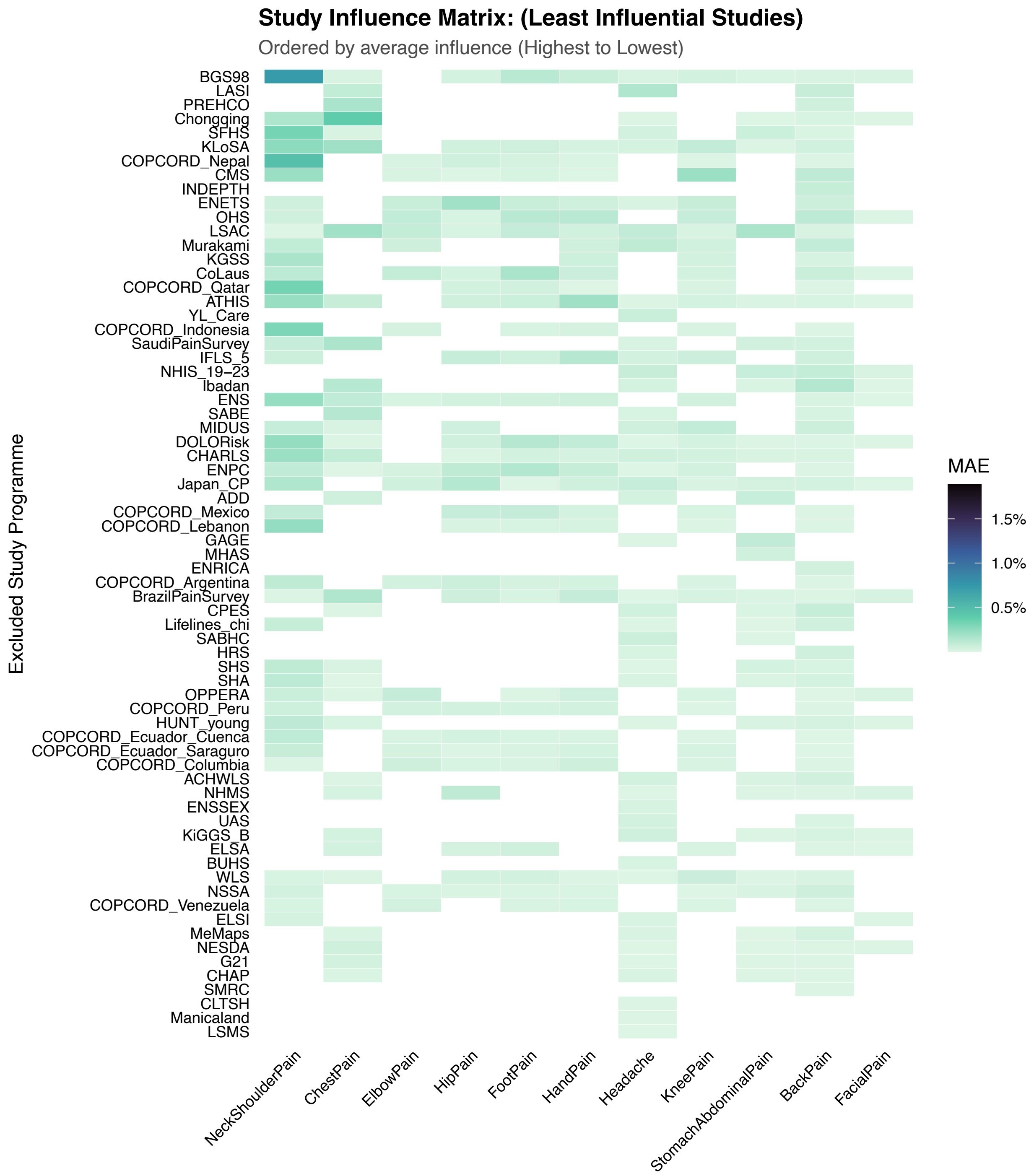

*Mean Absolute Difference (MAD) in predicted prevalence when specific studies (y-axis) are excluded from the model. Studies are ordered by average influence across all pain sites.*

**Section 6.2. Sensitivity Analysis of Methodological Covariates**

To assess the robustness of the estimated age–sex trajectories to study-level heterogeneity, we performed a sensitivity analysis ranking the influence of seven methodological covariates. We defined these covariates as follows:

- **Recall Period:** The reference time frame for pain reporting (e.g., "past week", "past month" or “past 3-12 months“).
- **Case Definition:** The stringency of the criteria used to classify pain (e.g., "chronic/frequent" vs. "general").
- **Survey Format:** The mode of administration (e.g., oral interview vs. written/online questionnaire).
- **Survey Setting:** The physical environment of data collection (e.g., in-person vs. remote).
- **Question Valence:** The phrasing of the pain item (True if pain questionnaire qualified by terms like “Bothered”, “Troubled”, “Suffered”, or “Interfered”, otherwise False).
- **Question Format:** The structure of response options (e.g., binary Yes/No vs. ordinal scales).
- **Sampling Strategy:** The method of participant recruitment (e.g., random probability sampling, convenience/web-based, or volunteer/biobank).

We specified a "naive" baseline mixed-effects model (natural spline for age [df=3], age–sex interaction, and random intercepts for study programme and world region) without methodological adjustments. We compared this baseline against separate models, each adding a single covariate as a fixed main effect. To quantify the population-level impact, we calculated weighted marginal predictions for all models. Deviations from the baseline were summarized as the Mean Absolute Difference (MAD) in percentage points (pp).

The analysis demonstrated high stability of the reference curves (Supplementary Figure 3). Across all pain sites and methodological factors, the mean absolute difference between the naive and adjusted models remained below 2.4 percentage points. This indicates that the specified random effects structure absorbed the majority of measurable methodological heterogeneity. As shown in Supplementary Figure 4, even under the adjustment of the maximally influencing covariate for each site, the magnitude of this shift was minimal.

The choice of recall period was the primary consistent driver of variation. Adjusting for recall period resulted in a mean trajectory shift of 1.2–2.4 pp for high-prevalence sites. Analysis of effect directionality (Supplementary Figure 5) confirmed that broader recall periods consistently yielded higher prevalence estimates than point prevalence. Consequently, recall period was retained as a fixed effect to ensure comparable reference standards. Case definition was also retained due to its clinical relevance, despite a moderate statistical impact (mean shift $\approx$ 0.5 pp).
In contrast, covariates like survey format, setting, sampling strategy, and valence had limited impacts (average mean shift $\approx$ 0.5 pp). Furthermore, these factors displayed inconsistent directionality across pain sites (e.g., "oral" surveys raised prevalence for some sites but lowered it for others), suggesting they may not introduce a systematic bias requiring global correction. Therefore, these factors were excluded from the final models.

### Supplementary Figure 3: Sensitivity of prevalence estimates to methodological covariates

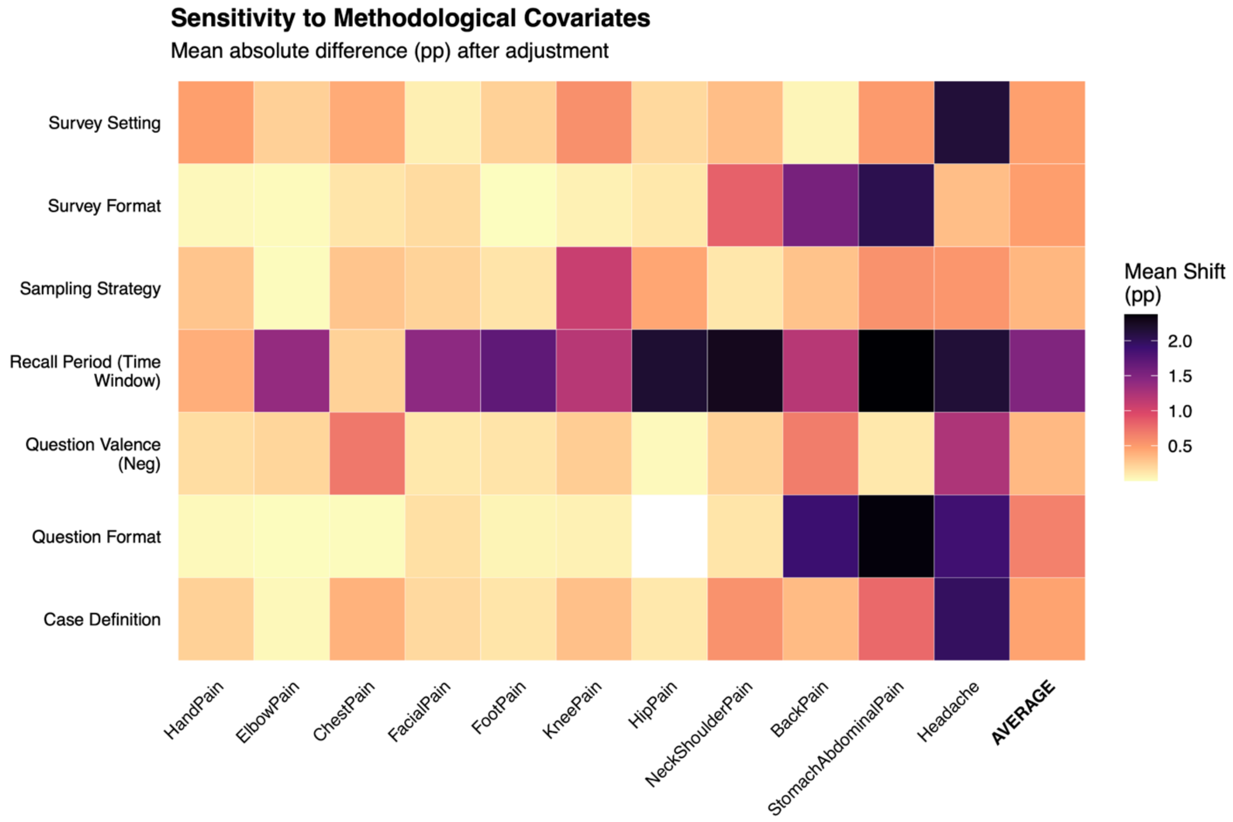

*Mean absolute shift in predicted prevalence (percentage points) when adjusting for specific study-level factors compared to the naive baseline model.*

**Supplementary Figure 4: Trajectory robustness under maximum methodological adjustment**

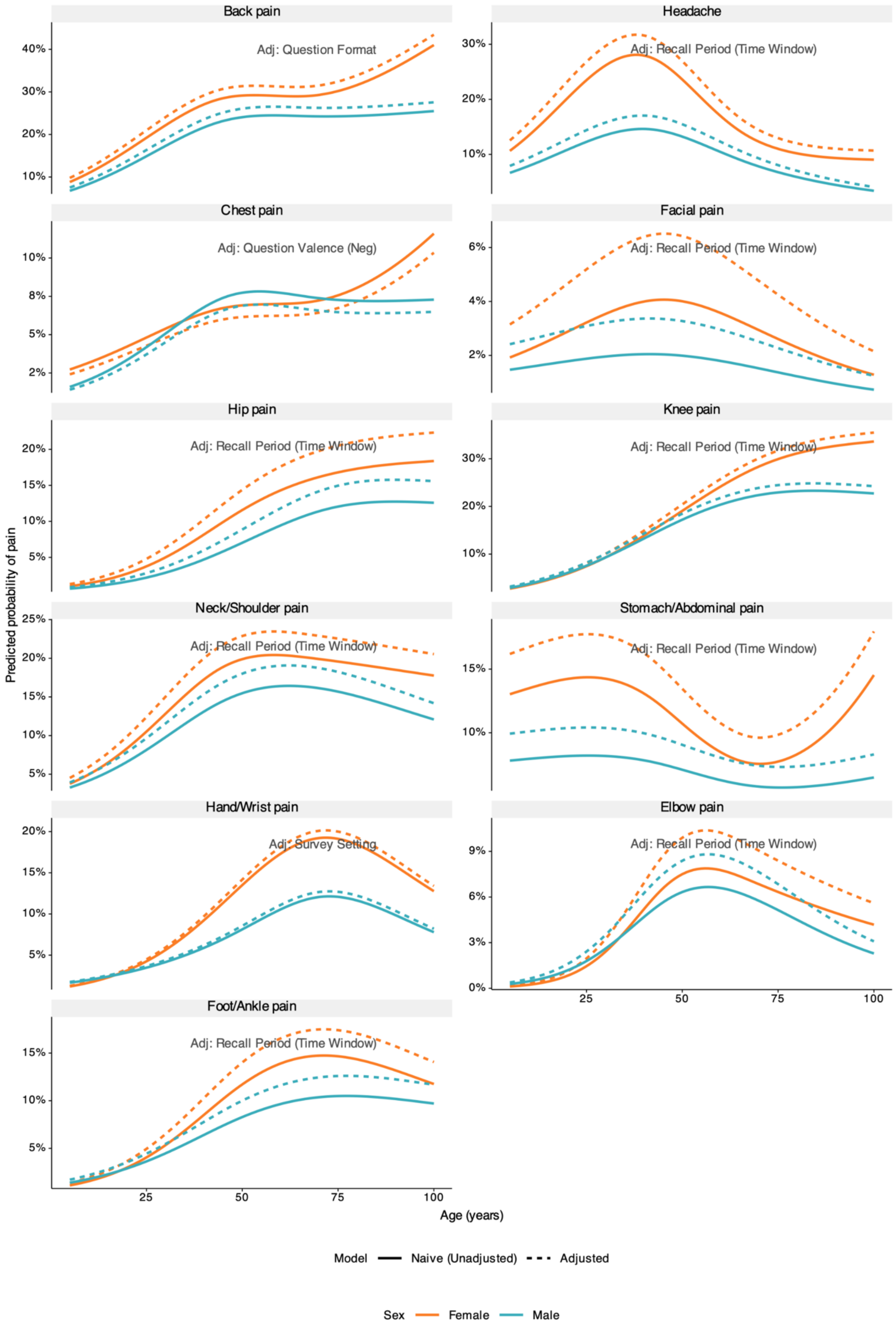

*Comparison of the naive model (solid lines) against the model adjusted for the single most influential covariate for each pain site (dashed lines).*

### Supplementary Figure 5: Directionality of methodological effects on reported prevalence

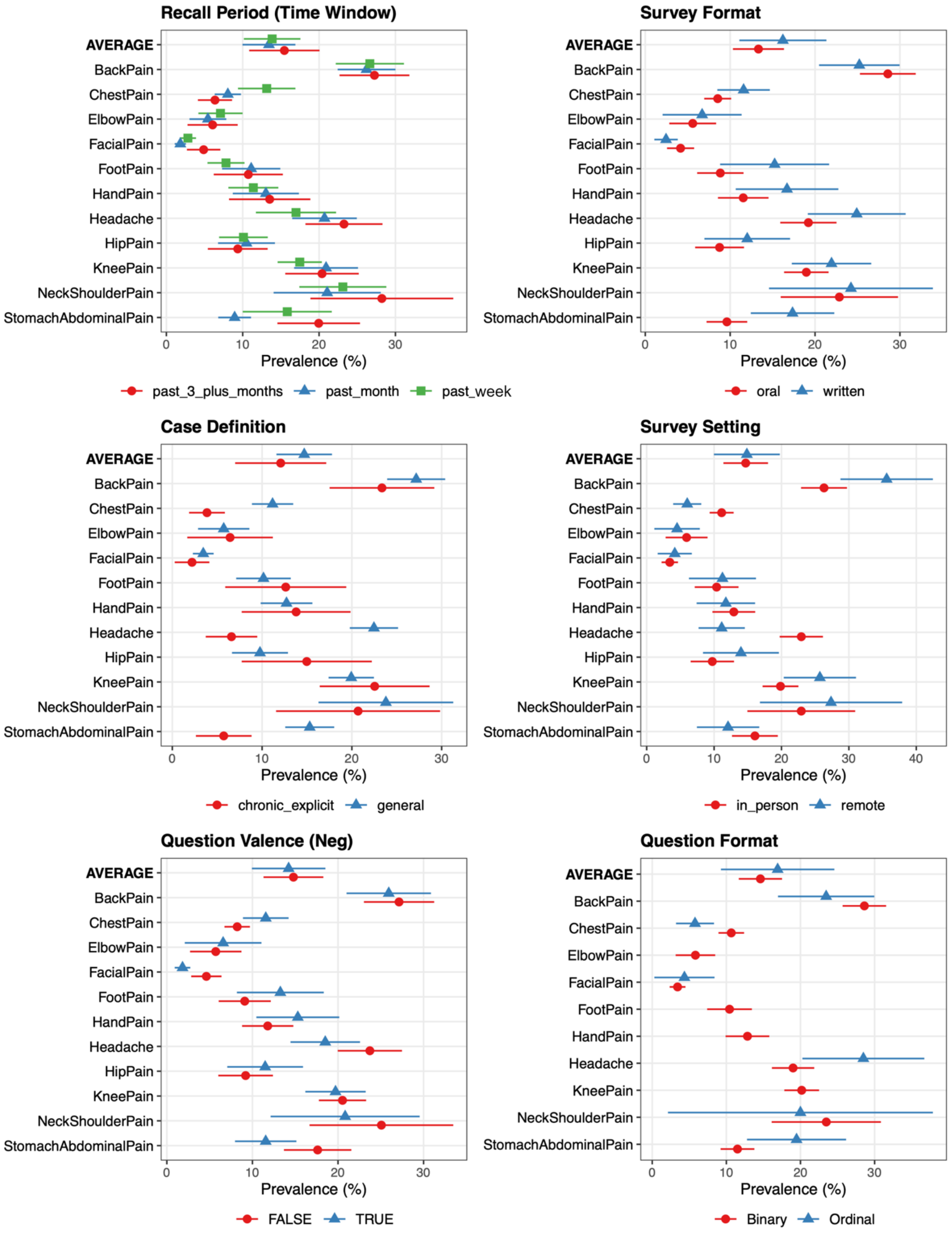

*Marginal predicted prevalence by covariate category, standardized to the global age-sex distribution. Points indicate the mean estimate; error bars indicate 95% confidence intervals.*

**Section 6.3. Sensitivity Analysis of Data Exclusion based on Recall Period**

To evaluate whether statistical adjustment for recall period materially influenced estimates, we conducted a data-exclusion sensitivity analysis comparing three analytical tiers. Tier Main (primary approach) included all recall periods (past week, past month, past ≥ 3 months) and incorporated a fixed-effect adjustment standardized to a past-month reference. This was compared with two restricted subsets. Tier A (Acute-Focused) excluded studies using past ≥ 3 month recall windows or explicit chronic pain definitions, retaining only past-week or past-month recall without chronic specification. Tier B (Strict) retained only studies explicitly using a past-month recall period. For Tier A, models were refit and standardized to past month; for Tier B, no recall adjustment was required. Absolute deviations between the primary model and the restricted subsets were calculated across the lifespan.

Trajectories from both restricted subsets showed high concordance with the primary model (Supplementary Figure 6). Tier A produced nearly identical estimates, with mean absolute deviations of 0.6 percentage points (Supplementary Figure 7), indicating strong alignment between adjusted past-week and past-month data.

The Strict subset (Tier B) generally tracked the primary model within 2.0 pp; however, greater variability was observed due to reduced sample size and geographic coverage. Deviations remained stable across the lifespan for most pain sites, though larger divergences (up to ~3.5 pp) were observed in the oldest age groups (≥ 80 years) for chest pain likely due to data sparsity in the strict subset. Overall, restricting the analysis to strictly past-month studies yielded trajectories substantively similar to the full adjusted model but with reduced precision, supporting the validity of using the full global dataset to maximize representativeness.

**Supplementary Figure 6: Deviation of restricted data subsets from the primary model**

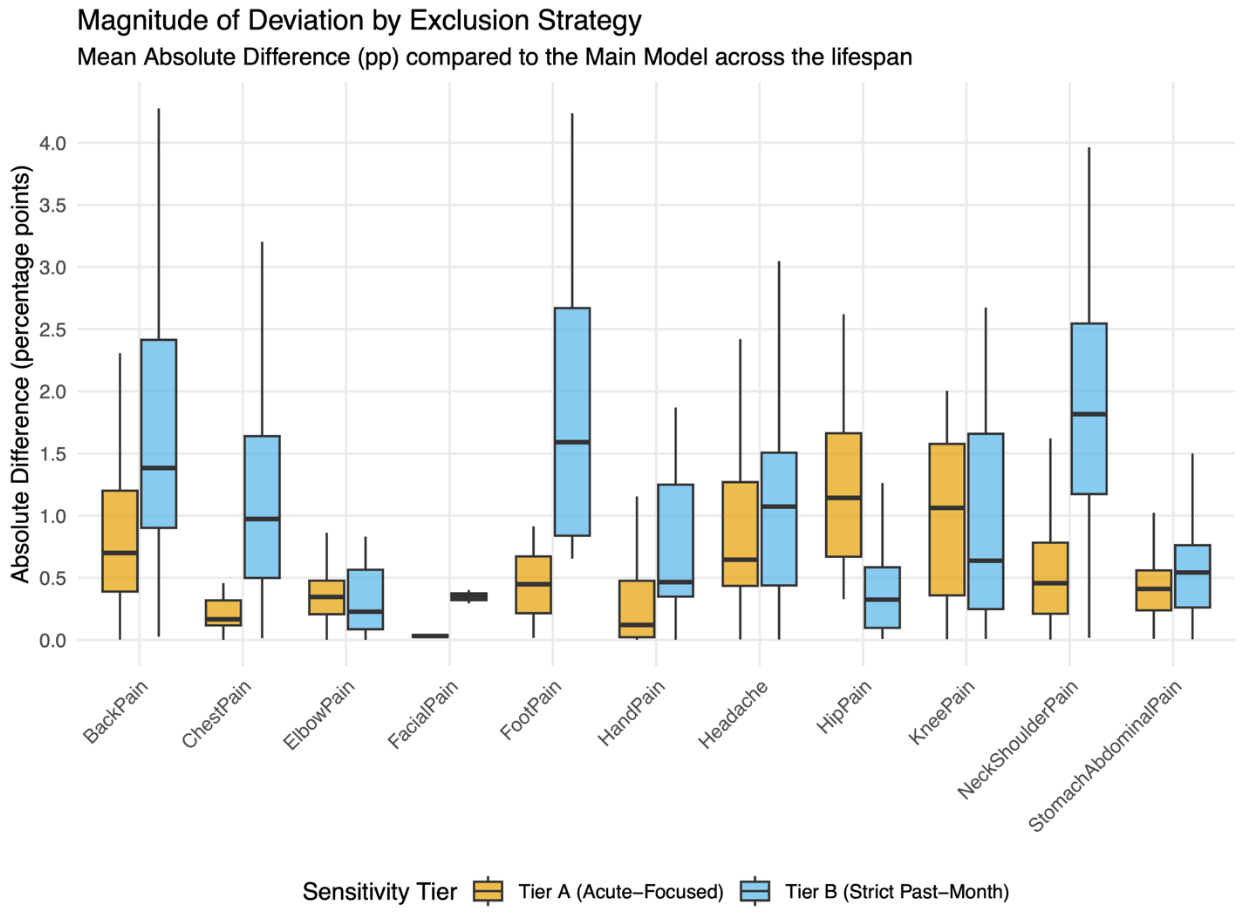

*Distribution of absolute differences in prevalence estimates for the Acute-Focused (Tier A) and Strict (Tier B) subsets relative to the primary model.*

**Supplementary Figure 7: Concordance between primary model and restricted data subsets**

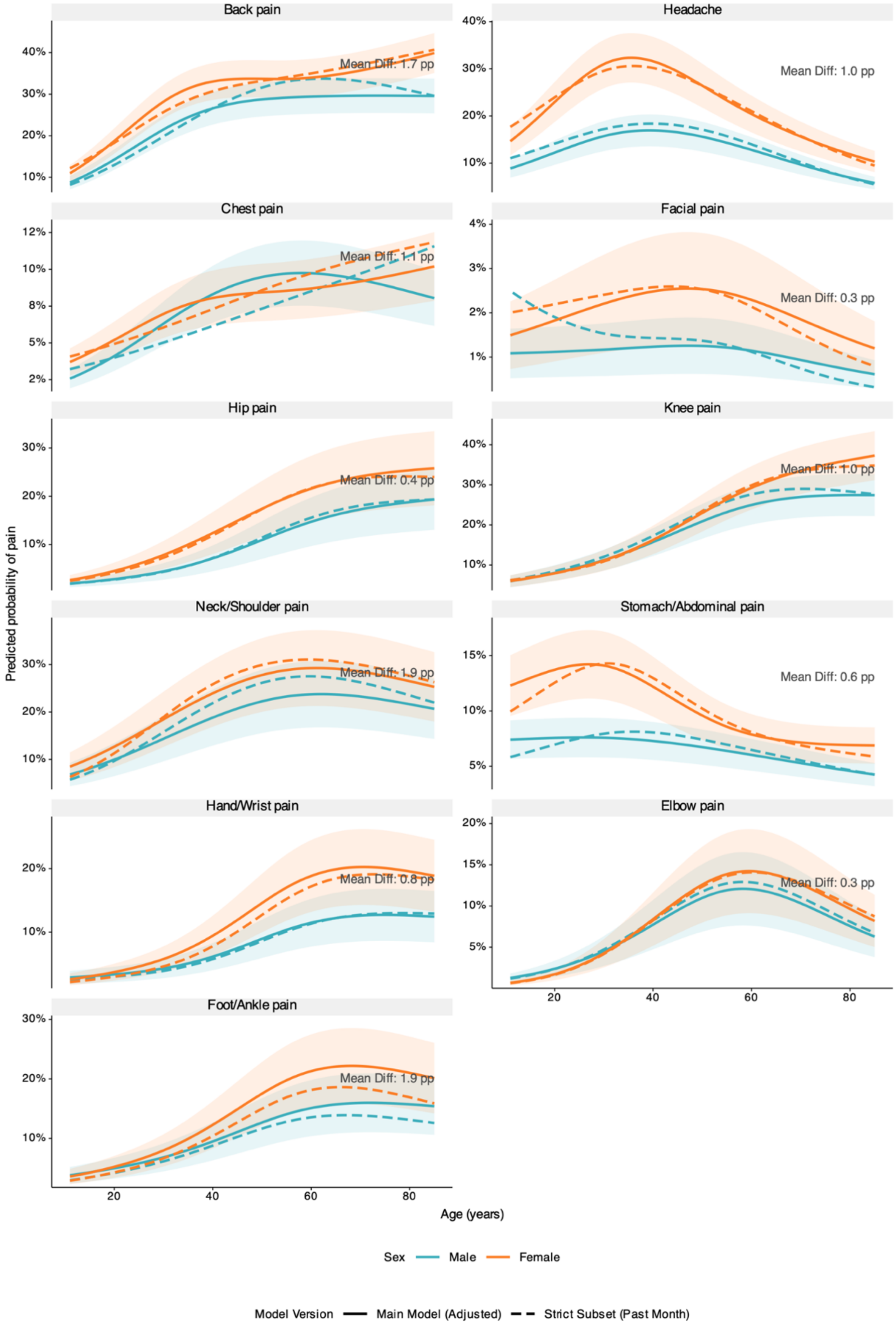

*The primary model is compared against a restricted subset containing only studies explicitly using a "past month" recall period (dashed line). Shaded regions indicate 95% confidence intervals.*

**Section 6.4. Sensitivity Analysis of Random-Effects Structure**

To evaluate the robustness of the estimated trajectories to model specification, we compared the primary model against an alternative random-effects structure allowing for study-specific random slopes for age. The primary model utilized crossed random intercepts (clustering by study and global region), assuming a common age trajectory shape while allowing prevalence levels to vary. The alternative model included random slopes to test whether the rate of aging varies systematically between study populations. Note that random slopes were only estimated for studies with sufficient within-study age variation to ensure statistical convergence.

For the majority of pain sites (9 of 11), the estimated age–sex trajectories were highly robust to changes in model structure (Supplementary Figure 8). The mean absolute difference between the primary and random-slope models for these sites was minimal (average MAD ≈ 0.9 pp; Supplementary Figure 9). However, the random-slope model produced notable deviations for two sites: headache (MAD = 3.9 pp), which exhibited a later age of decline, and back pain (MAD = 5.1 pp), which showed a steeper increase in late life compared to the primary model.

Despite these specific deviations, the primary random-intercept model was retained to ensure the most globally representative estimates. Estimating study-specific age slopes requires data spanning a wide age range; however, a substantial proportion of the included studies focus on specific life stages (e.g., exclusively adolescents or older adults) and lack the age span coverage to estimate a reliable age-slope. As such, relying on a random-slope specification would disproportionately weight results toward general population surveys with broad age spans, which predominantly come from high-income regions, while effectively discounting data from age-specific cohorts. The primary model integrates evidence from all study designs regardless of their internal age range, aiming to represent a more inclusive estimation of the global burden.

**Supplementary Figure 8: Magnitude of deviation between random-intercept and random-slope models**

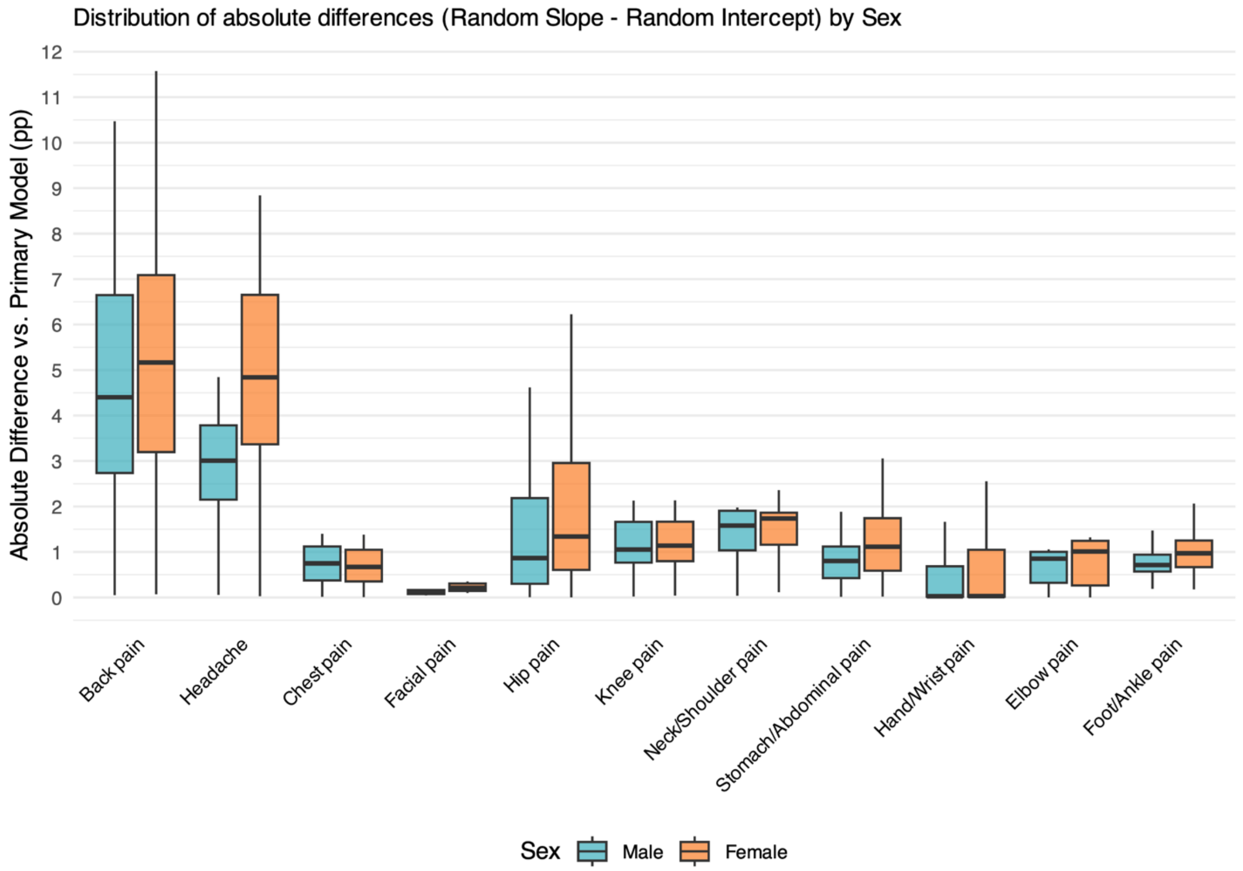

*Distribution of absolute differences in predicted prevalence across the lifespan when including study-specific random slopes.*

**Supplementary Figure 9: Sensitivity of trajectories to random-effects structure**

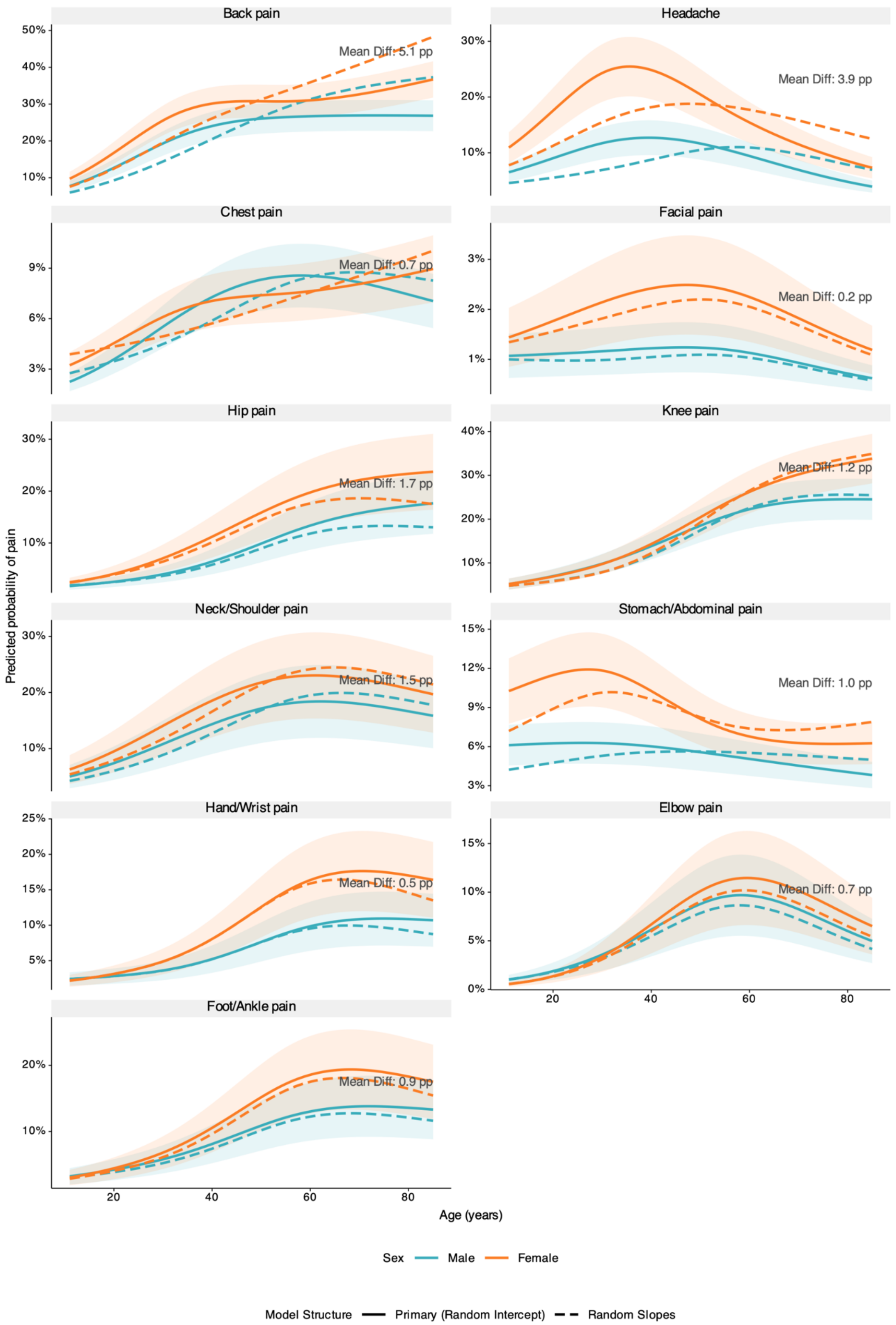

*Comparison of the primary random-intercept model (solid lines) against a model allowing for study-specific random slopes for age (dashed lines).*

**Section 6.5. Sensitivity Analysis of Functional Form (Age Spline Complexity)**

To determine the optimal functional form for the age trajectory, we performed a model selection sensitivity analysis comparing natural cubic splines with varying levels of complexity (degrees of freedom [df] = 1 to 4). We utilized a 5-fold cross-validation approach grouped by study to assess out-of-sample predictive performance, alongside Akaike Information Criterion (AIC) and Bayesian Information Criterion (BIC) on the full dataset to assess goodness-of-fit.

Performance metrics (Log Loss, AUC, and Brier score) were broadly similar across all degrees of freedom (Supplementary Table 4). For example, the AUC and Brier score for back pain remained stable at approximately 0.57 regardless of spline complexity, and Log Loss and Brier score differences were negligible (<0.01). This suggests that while age is a predictor of pain, increasing the complexity of the age curve does not significantly change the ranking of high-risk vs. low-risk individuals.

However, goodness-of-fit statistics calculated on the full dataset strongly favored non-linear specifications (Supplementary Figure 10). For all pain sites, AIC and BIC values decreased substantially moving from a linear specification (df=1) to a spline (df=3), with reductions often exceeding 10,000 points. This indicates that while a linear model can roughly rank risk, it significantly underfits the data and fails to accurately estimate the probability of pain at specific ages.

We selected df=3 (a natural spline) as the primary functional form for all sites. While df=4 provided marginal further improvements in BIC for some sites, the trajectory shapes for df=3 and df=4 were visually indistinguishable (Supplementary Figure 11), indicating that the additional complexity yielded little benefit in the epidemiological interpretation. Conversely, the linear model (df=1), despite having similar predictive power, lacks epidemiological plausibility; it fails to capture well-established non-monotonic life-course patterns, such as the early life peak followed by decline through late-life in headache. The df=3 specification provided the optimal balance, capturing the requisite biological curvature while maximizing parsimony and minimizing the risk of boundary artifacts (overfitting) in the oldest age groups where data density is lower.

**Supplementary Figure 10: Model selection statistics for functional form**

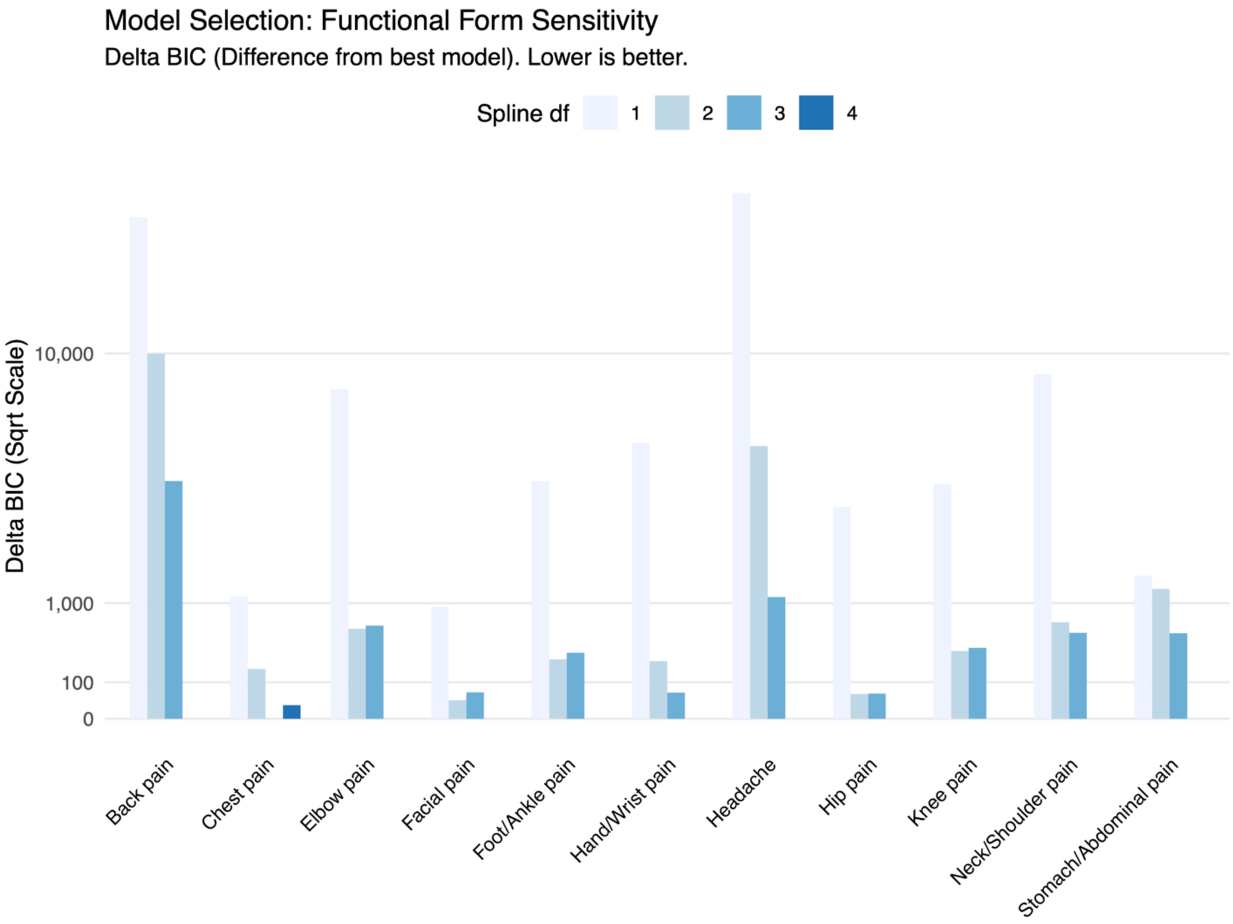

*Change in Bayesian Information Criterion (BIC) relative to the best-fitting model. Lower values indicate better fit. Note the square-root scale on the y-axis.*

**Supplementary Figure 11: Sensitivity of age trajectories to spline complexity**

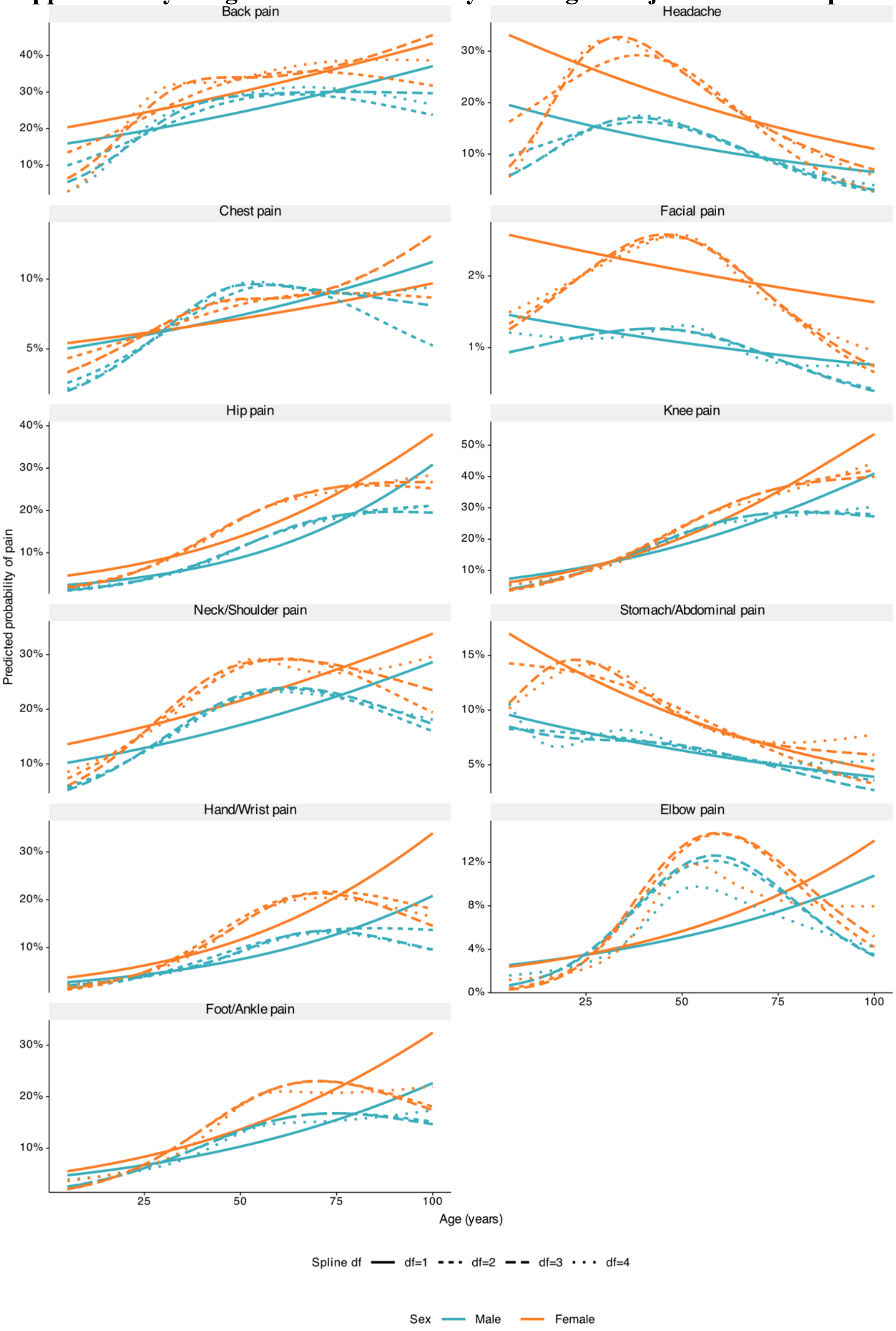

*Overlaid trajectories derived from natural cubic splines with varying degrees of freedom (df=1 to df=4).*

**Supplementary Table 4: Model selection metrics for age-trajectory functional form**

| Pain site | Spline complexity | AUC | Log loss | Brier | AIC | BIC |
|---|---|---|---|---|---|---|
| Back pain | 1 | 0.571 | 0.585 | 0.197 | 5488518.8 | 5488639.5 |
| Back pain | 2 | 0.573 | 0.584 | 0.197 | 5479608.5 | 5479755.9 |
| Back pain | 3 | 0.574 | 0.584 | 0.197 | 5473836.2 | 5474010.5 |
| Back pain | 4 | 0.574 | 0.585 | 0.197 | 5469576.6 | 5469777.7 |
| Chest pain | 1 | 0.562 | 0.309 | 0.082 | 853833.3 | 853942.3 |
| Chest pain | 2 | 0.568 | 0.307 | 0.082 | 852873.1 | 853006.3 |
| Chest pain | 3 | 0.569 | 0.307 | 0.082 | 852663.4 | 852820.8 |
| Chest pain | 4 | 0.569 | 0.307 | 0.082 | 852653.1 | 852834.7 |
| Elbow pain | 1 | 0.571 | 0.233 | 0.057 | 507045.7 | 507153.5 |
| Elbow pain | 2 | 0.618 | 0.233 | 0.058 | 499486.5 | 499618.2 |
| Elbow pain | 3 | 0.617 | 0.234 | 0.058 | 499505.2 | 499660.9 |
| Elbow pain | 4 | 0.626 | 0.229 | 0.057 | 498831.4 | 499011.1 |
| Facial pain | 1 | 0.616 | 0.149 | 0.034 | 458326.2 | 458436.8 |
| Facial pain | 2 | 0.617 | 0.15 | 0.034 | 457392.7 | 457527.9 |
| Facial pain | 3 | 0.617 | 0.15 | 0.034 | 457394.7 | 457554.5 |
| Facial pain | 4 | 0.618 | 0.15 | 0.034 | 457318 | 457502.4 |
| Foot/Ankle pain | 1 | 0.609 | 0.341 | 0.097 | 915301.6 | 915411.2 |
| Foot/Ankle pain | 2 | 0.613 | 0.342 | 0.097 | 911311.6 | 911445.6 |
| Foot/Ankle pain | 3 | 0.613 | 0.342 | 0.097 | 911349.6 | 911508 |
| Foot/Ankle pain | 4 | 0.612 | 0.341 | 0.097 | 910999.3 | 911182 |
| Hand/Wrist pain | 1 | 0.631 | 0.381 | 0.112 | 1056832.7 | 1056942.5 |
| Hand/Wrist pain | 2 | 0.634 | 0.379 | 0.112 | 1051358.3 | 1051492.5 |
| Hand/Wrist pain | 3 | 0.634 | 0.379 | 0.112 | 1051136.7 | 1051295.3 |
| Hand/Wrist pain | 4 | 0.635 | 0.379 | 0.112 | 1051060.9 | 1051244 |
| Headache | 1 | 0.609 | 0.499 | 0.161 | 3514562.9 | 3514680.8 |
| Headache | 2 | 0.618 | 0.505 | 0.163 | 3499415.9 | 3499560 |
| Headache | 3 | 0.616 | 0.511 | 0.166 | 3494926.7 | 3495097 |
| Headache | 4 | 0.616 | 0.509 | 0.165 | 3493791.1 | 3493987.6 |
| Hip pain | 1 | 0.63 | 0.327 | 0.092 | 1265875.7 | 1265988.6 |
| Hip pain | 2 | 0.633 | 0.331 | 0.093 | 1262526.6 | 1262664.6 |
| Hip pain | 3 | 0.633 | 0.331 | 0.093 | 1262502.8 | 1262665.9 |
| Hip pain | 4 | 0.634 | 0.33 | 0.093 | 1262430.4 | 1262618.6 |
| Knee pain | 1 | 0.615 | 0.489 | 0.156 | 1917582 | 1917694.6 |
| Knee pain | 2 | 0.614 | 0.491 | 0.157 | 1913775.1 | 1913912.7 |
| Knee pain | 3 | 0.614 | 0.491 | 0.157 | 1913783.3 | 1913945.9 |
| Knee pain | 4 | 0.615 | 0.49 | 0.157 | 1913380.9 | 1913568.5 |

| | | | | | | |
|---|---|---|---|---|---|---|
| Neck/Shoulder pain | 1 | 0.565 | 0.566 | 0.186 | 2375098.6 | 2375212.5 |
| Neck/Shoulder pain | 2 | 0.577 | 0.555 | 0.182 | 2366876.2 | 2367015.5 |
| Neck/Shoulder pain | 3 | 0.578 | 0.554 | 0.182 | 2366706.5 | 2366871.1 |
| Neck/Shoulder pain | 4 | 0.579 | 0.555 | 0.182 | 2366127.7 | 2366317.6 |
| Stomach/Abdominal pain | 1 | 0.584 | 0.385 | 0.114 | 2042452.7 | 2042567.4 |
| Stomach/Abdominal pain | 2 | 0.582 | 0.386 | 0.114 | 2042155.6 | 2042295.8 |
| Stomach/Abdominal pain | 3 | 0.579 | 0.387 | 0.115 | 2041413.6 | 2041579.3 |
| Stomach/Abdominal pain | 4 | 0.577 | 0.388 | 0.115 | 2040840.5 | 2041031.7 |

*Comparison of predictive performance and goodness-of-fit across natural cubic splines with varying degrees of freedom (df=1–4). AUC, Log Loss, and Brier Score represent out-of-sample performance averaged across 5 cross-validation folds grouped by study. AIC and BIC represent goodness-of-fit calculated on the full dataset. AIC=Akaike Information Criterion; AUC=Area Under the Curve; BIC=Bayesian Information Criterion; df=degrees of freedom.*

**Section 6.6. Sensitivity Analysis of Missing Risk Factor Data**

To evaluate the potential influence of selection bias arising from missing covariate data, we performed a sensitivity analysis comparing the primary complete-case approach against an imputed dataset. The primary models excluded participants with missing data on BMI, smoking status, or income. Given the scale of the pooled dataset, conventional multiple imputation (e.g., MICE) was computationally prohibitive; we therefore employed a single stochastic imputation based on a Stochastic Gradient Descent (SGD) classifier, which scales efficiently to large samples. Because single imputation does not propagate imputation uncertainty into variance estimates, this analysis was designed to assess the direction and magnitude of potential bias rather than to replace the primary complete-case framework.

The imputation model was fitted within each study separately using predictors including education level, employment status, marital status, age, sex, country, and pain site; imputed datasets were then pooled across studies before refitting the exposure models. To avoid unreliable extrapolation, studies with excessive missingness (>50%) for a given variable were excluded from the imputation process. We compared the age-adjusted Relative Risks (RR) derived from the complete-case data against those derived from the imputed data.

Results demonstrated high robustness across all exposure–outcome associations (Supplementary Figure 12). For BMI and smoking status, point estimates and 95% confidence intervals (CIs) from the imputed models substantially overlapped with those from the complete-case analysis across all pain sites. For income (Lower vs. Upper quintile), the direction and magnitude of associations were largely consistent between methods. However, for headache, hip, knee, and neck-shoulder pain the imputed models yielded slightly higher relative risk estimates than the complete-case analysis. These findings suggest that the primary complete-case results are conservative and not substantively biased by the exclusion of participants with missing covariates. Consequently, the primary complete-case analysis was retained to maintain transparency and avoid over-reliance on modeled assumptions.

**Supplementary Figure 12: Sensitivity of relative risk estimates to missing data imputation**

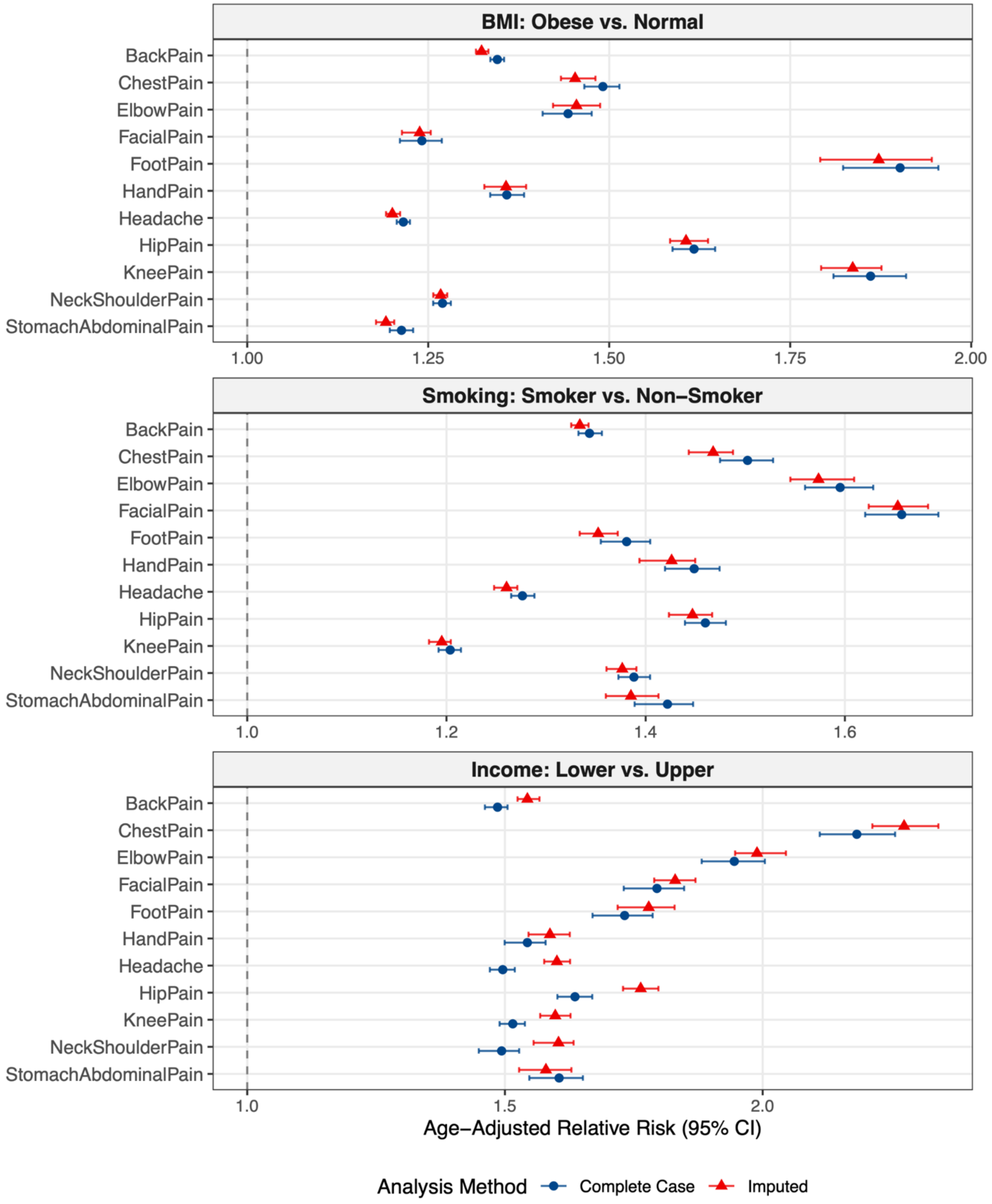

*Comparison of age-adjusted Relative Risks (RR) derived from the primary Complete Case analysis (Blue circles) versus the Imputed sensitivity analysis (Red triangles) Error bars represent 95% confidence intervals.*

### Section 7. Individual-level risk factor gradients of pain across the lifespan

Age-specific gradients in pain prevalence were estimated for sex, smoking status, body mass index category, and income quintile. For each pain site, model-predicted trajectories across age are displayed by exposure category. For smoking analyses, the analytic sample was restricted to participants aged 13 years and older, as self-reported smoking was not available younger children. Across exposures, the strongest gradients were generally observed between early and mid-adulthood (ages 18–65), with attenuation in older age groups for, particularly for smoking and income. This late-life attenuation may partly reflect selective survival, as individuals with the highest exposure burden are less likely to survive into older age, potentially underestimating the true exposure–pain association in these groups. All three exposures were associated with elevated prevalence across all pain sites. Income gradients were generally the largest in magnitude, likely reflecting its role as a proxy for multiple structural determinants of health, followed by obesity and then smoking. Obesity showed disproportionately strong associations at lower-limb sites (knee, foot, hip), consistent with cumulative mechanical load. Additionally, age-binned and age-adjusted (overall) risk ratios are provided for smoking, body mass index, and income across all pain sites, as well as sex-based risk ratios for all pain sites plus three secondary phenotypes: any bodily pain, high-intensity pain (NRS $\geq 7$), and generalized pain.

**Supplementary Table 5. Risk ratios for sex and health exposures across pain sites.**

| Pain site | Overall | 5–17 y | 18–35 y | 35–50 y | 50–65 y | 65–80 y | 80–100 y |
|---|---|---|---|---|---|---|---|
| **Sex (Female vs. Male)** | | | | | | | |
| Back | 1.28 (1.27–1.28) | 1.27 (1.26–1.28) | 1.30 (1.29–1.31) | 1.23 (1.22–1.23) | 1.17 (1.17–1.18) | 1.23 (1.22–1.23) | 1.41 (1.40–1.43) |
| Knee | 1.21 (1.20–1.22) | 0.96 (0.94–0.98) | 1.00 (0.99–1.02) | 1.07 (1.06–1.08) | 1.16 (1.15–1.17) | 1.26 (1.24–1.27) | 1.38 (1.35–1.42) |
| Headache | 1.78 (1.76–1.79) | 1.63 (1.62–1.65) | 1.92 (1.89–1.95) | 1.80 (1.77–1.82) | 1.62 (1.60–1.63) | 1.63 (1.62–1.65) | 1.94 (1.90–1.98) |
| Hip | 1.44 (1.42–1.46) | 1.55 (1.50–1.60) | 1.69 (1.66–1.73) | 1.64 (1.61–1.66) | 1.48 (1.46–1.50) | 1.37 (1.35–1.39) | 1.35 (1.31–1.40) |
| Neck/shoulder | 1.27 (1.26–1.29) | 1.31 (1.28–1.34) | 1.32 (1.30–1.33) | 1.28 (1.26–1.29) | 1.22 (1.22–1.23) | 1.23 (1.21–1.24) | 1.33 (1.29–1.36) |
| Hand | 1.52 (1.49–1.55) | 0.90 (0.86–0.94) | 1.31 (1.29–1.34) | 1.57 (1.55–1.60) | 1.61 (1.58–1.65) | 1.58 (1.54–1.62) | 1.54 (1.48–1.60) |
| Foot | 1.29 (1.27–1.32) | 0.93 (0.89–0.97) | 1.17 (1.14–1.19) | 1.33 (1.30–1.35) | 1.39 (1.36–1.41) | 1.36 (1.33–1.39) | 1.26 (1.21–1.31) |
| Stomach/abdominal | 1.62 (1.60–1.64) | 1.50 (1.48–1.53) | 1.84 (1.82–1.87) | 1.64 (1.63–1.66) | 1.40 (1.39–1.42) | 1.42 (1.39–1.44) | 1.72 (1.66–1.79) |
| Elbow | 1.16 (1.14–1.19) | 0.59 (0.55–0.63) | 0.95 (0.93–0.98) | 1.11 (1.09–1.13) | 1.17 (1.15–1.19) | 1.23 (1.20–1.26) | 1.36 (1.28–1.45) |
| Chest | 1.09 (1.07–1.12) | 1.48 (1.42–1.54) | 1.10 (1.08–1.13) | 0.94 (0.93–0.95) | 0.92 (0.91–0.93) | 1.03 (1.01–1.05) | 1.33 (1.25–1.43) |
| Facial | 1.83 (1.80–1.87) | 1.44 (1.35–1.54) | 1.75 (1.71–1.81) | 1.97 (1.93–2.02) | 2.00 (1.96–2.05) | 1.93 (1.87–1.99) | 1.83 (1.70–1.97) |
| Any bodily pain | 1.16 (1.15–1.17) | 1.26 (1.25–1.27) | 1.20 (1.19–1.21) | 1.14 (1.13–1.15) | 1.11 (1.10–1.11) | 1.12 (1.11–1.12) | 1.19 (1.17–1.20) |
| High-intensity pain | 1.64 (1.60–1.66) | 1.65 (1.53–1.74) | 1.46 (1.41–1.48) | 1.32 (1.29–1.35) | 1.29 (1.26–1.31) | 1.52 (1.47–1.54) | 2.39 (2.14–2.51) |
| Generalized pain | 1.49 (1.46–1.53) | 1.03 (0.96–1.11) | 1.22 (1.19–1.26) | 1.39 (1.36–1.42) | 1.51 (1.48–1.54) | 1.57 (1.53–1.62) | 1.60 (1.51–1.70) |
| **Income quintile (Lower vs. Upper)** | | | | | | | |
| Back | 1.49 (1.46–1.50) | 0.97 (0.95–1.00) | 1.29 (1.27–1.32) | 1.53 (1.50–1.56) | 1.56 (1.53–1.59) | 1.42 (1.39–1.44) | 1.19 (1.16–1.22) |
| Knee | 1.52 (1.49–1.54) | 1.01 (0.97–1.05) | 1.36 (1.33–1.39) | 1.59 (1.56–1.62) | 1.58 (1.54–1.61) | 1.43 (1.40–1.46) | 1.21 (1.17–1.24) |
| Headache | 1.50 (1.47–1.52) | 1.10 (1.06–1.13) | 1.32 (1.30–1.35) | 1.54 (1.50–1.58) | 1.66 (1.62–1.70) | 1.67 (1.62–1.72) | 1.53 (1.45–1.61) |
| Hip | 1.64 (1.60–1.67) | 1.20 (1.13–1.27) | 1.57 (1.52–1.63) | 1.78 (1.73–1.83) | 1.69 (1.64–1.74) | 1.45 (1.41–1.49) | 1.14 (1.09–1.18) |
| Neck/shoulder | 1.49 (1.45–1.53) | 0.93 (0.89–0.96) | 1.26 (1.23–1.28) | 1.51 (1.48–1.54) | 1.57 (1.54–1.60) | 1.48 (1.45–1.52) | 1.30 (1.25–1.35) |
| Hand | 1.54 (1.50–1.58) | 1.15 (1.07–1.23) | 1.58 (1.52–1.64) | 1.80 (1.74–1.86) | 1.65 (1.59–1.70) | 1.28 (1.25–1.32) | 0.96 (0.92–1.00) |
| Foot | 1.73 (1.67–1.79) | 0.94 (0.87–1.01) | 1.49 (1.43–1.55) | 1.90 (1.82–1.98) | 1.88 (1.79–1.97) | 1.56 (1.49–1.62) | 1.19 (1.12–1.25) |
| Stomach/abdominal | 1.61 (1.55–1.65) | 1.15 (1.09–1.21) | 1.44 (1.38–1.49) | 1.66 (1.60–1.73) | 1.67 (1.61–1.73) | 1.51 (1.44–1.58) | 1.17 (1.06–1.29) |
| Elbow | 1.94 (1.88–2.00) | 1.25 (1.13–1.37) | 1.65 (1.57–1.72) | 1.90 (1.83–1.97) | 1.98 (1.90–2.05) | 1.94 (1.84–2.05) | 1.83 (1.65–2.00) |
| Chest | 2.18 (2.11–2.26) | 1.41 (1.26–1.55) | 2.00 (1.89–2.12) | 2.42 (2.32–2.53) | 2.20 (2.12–2.29) | 1.70 (1.60–1.79) | 0.98 (0.86–1.11) |
| Facial | 1.79 (1.73–1.85) | 0.86 (0.81–0.92) | 1.41 (1.36–1.46) | 1.88 (1.82–1.94) | 2.04 (1.97–2.10) | 1.78 (1.68–1.88) | 1.35 (1.22–1.49) |

| Pain site | Overall | 5–17 y | 18–35 y | 35–50 y | 50–65 y | 65–80 y | 80–100 y |
|---|---|---|---|---|---|---|---|
| **Smoking (Smoker vs. Never-Smoker)*** | | | | | | | |
| Back | 1.34 (1.33–1.36) | 1.58 (1.55–1.60) | 1.47 (1.46–1.49) | 1.36 (1.35–1.37) | 1.25 (1.24–1.26) | 1.17 (1.16–1.17) | 1.07 (1.06–1.09) |
| Knee | 1.20 (1.19–1.21) | 1.68 (1.63–1.72) | 1.55 (1.52–1.57) | 1.36 (1.34–1.38) | 1.16 (1.15–1.17) | 0.98 (0.97–0.99) | 0.79 (0.77–0.82) |
| Headache | 1.28 (1.26–1.29) | 1.51 (1.47–1.54) | 1.39 (1.37–1.41) | 1.25 (1.24–1.27) | 1.17 (1.15–1.18) | 1.11 (1.09–1.13) | 1.05 (1.01–1.09) |
| Hip | 1.46 (1.44–1.48) | 1.61 (1.54–1.68) | 1.67 (1.63–1.71) | 1.62 (1.59–1.65) | 1.45 (1.43–1.47) | 1.25 (1.23–1.27) | 1.01 (0.98–1.05) |
| Neck/shoulder | 1.39 (1.37–1.40) | 1.55 (1.51–1.58) | 1.54 (1.52–1.57) | 1.46 (1.44–1.48) | 1.32 (1.30–1.33) | 1.14 (1.13–1.16) | 0.93 (0.91–0.96) |
| Hand | 1.45 (1.42–1.47) | 2.22 (2.12–2.31) | 1.98 (1.93–2.03) | 1.68 (1.64–1.71) | 1.37 (1.34–1.39) | 1.08 (1.06–1.11) | 0.87 (0.83–0.91) |
| Foot | 1.38 (1.35–1.40) | 1.67 (1.58–1.76) | 1.64 (1.60–1.69) | 1.54 (1.50–1.57) | 1.34 (1.31–1.37) | 1.11 (1.08–1.13) | 0.90 (0.85–0.95) |
| Stomach/abdominal | 1.42 (1.39–1.45) | 1.57 (1.51–1.63) | 1.51 (1.48–1.55) | 1.42 (1.40–1.45) | 1.33 (1.30–1.36) | 1.24 (1.20–1.27) | 1.12 (1.05–1.20) |
| Elbow | 1.60 (1.56–1.63) | 1.66 (1.55–1.77) | 1.83 (1.77–1.89) | 1.81 (1.76–1.85) | 1.53 (1.49–1.56) | 1.12 (1.08–1.17) | 0.78 (0.72–0.84) |
| Chest | 1.50 (1.47–1.53) | 1.36 (1.29–1.44) | 1.52 (1.47–1.57) | 1.62 (1.59–1.65) | 1.48 (1.45–1.50) | 1.23 (1.19–1.26) | 0.86 (0.80–0.91) |
| Facial | 1.66 (1.62–1.69) | 2.06 (1.97–2.16) | 1.93 (1.88–1.98) | 1.74 (1.70–1.77) | 1.48 (1.44–1.52) | 1.18 (1.14–1.23) | 0.91 (0.84–0.97) |
| **Obesity (Obese vs. Normal)** | | | | | | | |
| Back | 1.35 (1.34–1.36) | 1.34 (1.33–1.36) | 1.33 (1.32–1.34) | 1.32 (1.31–1.33) | 1.33 (1.32–1.34) | 1.36 (1.35–1.37) | 1.40 (1.38–1.42) |
| Knee | 1.85 (1.81–1.89) | 1.02 (0.98–1.05) | 1.52 (1.49–1.55) | 1.96 (1.92–2.01) | 2.03 (1.97–2.09) | 1.89 (1.83–1.94) | 1.63 (1.58–1.67) |
| Headache | 1.21 (1.20–1.22) | 1.31 (1.29–1.32) | 1.25 (1.24–1.26) | 1.20 (1.19–1.21) | 1.14 (1.13–1.15) | 1.07 (1.05–1.08) | 0.96 (0.94–0.98) |
| Hip | 1.61 (1.53–1.67) | 0.89 (0.82–0.96) | 1.28 (1.24–1.32) | 1.63 (1.59–1.66) | 1.71 (1.66–1.77) | 1.63 (1.57–1.69) | 1.45 (1.40–1.49) |
| Neck/shoulder | 1.27 (1.25–1.29) | 1.01 (0.98–1.04) | 1.14 (1.13–1.16) | 1.25 (1.23–1.27) | 1.30 (1.28–1.32) | 1.30 (1.28–1.33) | 1.29 (1.26–1.32) |
| Hand | 1.36 (1.33–1.38) | 1.12 (1.06–1.19) | 1.32 (1.28–1.35) | 1.43 (1.40–1.45) | 1.41 (1.38–1.44) | 1.32 (1.29–1.35) | 1.22 (1.18–1.26) |
| Foot | 1.89 (1.81–1.96) | 1.29 (1.19–1.38) | 1.76 (1.69–1.83) | 2.08 (1.98–2.17) | 2.04 (1.93–2.16) | 1.78 (1.69–1.88) | 1.49 (1.41–1.56) |
| Stomach/abdominal | 1.20 (1.19–1.22) | 1.17 (1.15–1.19) | 1.24 (1.22–1.25) | 1.24 (1.22–1.26) | 1.21 (1.19–1.22) | 1.16 (1.13–1.19) | 1.08 (1.03–1.13) |
| Elbow | 1.44 (1.41–1.48) | 1.17 (1.06–1.27) | 1.37 (1.31–1.43) | 1.47 (1.42–1.52) | 1.46 (1.42–1.51) | 1.40 (1.35–1.45) | 1.29 (1.22–1.37) |
| Chest | 1.49 (1.46–1.51) | 0.85 (0.80–0.90) | 1.20 (1.16–1.23) | 1.53 (1.50–1.56) | 1.56 (1.54–1.59) | 1.41 (1.37–1.44) | 1.09 (1.03–1.14) |
| Facial | 1.24 (1.22–1.27) | 1.03 (0.96–1.09) | 1.16 (1.12–1.19) | 1.25 (1.22–1.28) | 1.29 (1.26–1.32) | 1.26 (1.22–1.30) | 1.19 (1.11–1.27) |

** For smoking, the adolescent age band corresponds to ages 13–17 years and is shown under the 5–17 column*

*Age stratified risk ratios (RR) for prevalence of self-reported pain outcomes comparing female versus male, lower versus upper income quintile, smoker versus never-smoker, and obese versus normal weight. Values are RR (95% CI)*

**Section 8. Population attributable fraction by region and country**

Population attributable fractions (PAFs) were estimated for obesity, smoking, and low household income across 11 anatomical pain sites and stratified by world region and country. For each exposure, age- and sex-specific PAFs were computed at the country level by combining study-derived risk ratios with country-specific exposure prevalence from the Global Burden of Disease study and population denominators (Section 5.4). The combined PAF was calculated using the multiplicative independence formula to account for co-occurrence of exposures. Regional and global estimates were obtained as case-weighted averages across the 11 anatomical sites.

Globally, an estimated 18·3% of the site-specific pain burden was jointly attributable to the three modifiable exposures (Supplementary Table 6). Obesity contributed the largest share in eastern Europe (13·0%), northern America (14·6%), and north Africa and west Asia (13·3%), whereas its contribution was below 5% in sub-Saharan Africa and central and southern Asia. Smoking-attributable fractions were highest in eastern Europe (8·4%) and western Europe (7·2%) and lowest in sub-Saharan Africa (2·5%). Income-attributable fractions were more uniform across regions (6·7%–8·5%), consistent with the use of within-study relative income ranking. The combined PAF ranged from 12·6% in sub-Saharan Africa to 27·1% in eastern Europe, indicating that the composition and magnitude of modifiable pain burden differ substantially across global contexts (Extended Data Figure 5).

### Supplementary Table 6: Population attributable fractions for pain by exposure and world region

| Region | Obesity PAF (%) | Smoking PAF (%) | Income PAF (%) | Combined PAF (%) |
|---|---|---|---|---|
| **Eastern Europe** | 13 | 8.4 | 8.5 | 27.1 |
| **Northern America** | 14.6 | 5.1 | 8.3 | 25.4 |
| **North Africa & West Asia** | 13.3 | 5.4 | 7.8 | 24 |
| **Oceania** | 11.2 | 6.6 | 8 | 23.5 |
| **Western Europe** | 10.1 | 7.2 | 8.3 | 23.3 |
| **Latin America** | 11.2 | 4.6 | 8 | 21.8 |
| **East & Southeast Asia** | 3.8 | 6.9 | 8.4 | 17.9 |
| **Central & Southern Asia** | 2.7 | 4.4 | 7.6 | 13.9 |
| **Sub-Saharan Africa** | 4.2 | 2.5 | 6.7 | 12.6 |
| **Global** | 6.3 | 5.5 | 7.9 | 18.3 |

*Case-weighted population attributable fractions (PAF, %) for obesity, smoking, and low household income, averaged across 11 anatomical pain sites. The combined PAF was calculated as* $1 - (1 - PAF_{obesity})(1 - PAF_{smoking})(1 - PAF_{income})$*, accounting for co-occurrence of exposures; individual exposure-specific PAFs may therefore sum to more than the combined value. Because individuals may report pain at multiple anatomical sites, site-specific pain cases are not mutually exclusive; estimates represent the proportion of the total site-specific pain burden attributable to each exposure rather than the proportion of individuals with any pain. Obesity and smoking prevalence were obtained from the Global Burden of Disease study (obesity: GBD 2021; smoking: GBD 2015). Income was operationalised as within-study relative rank (quintiles), with a fixed prevalence of 20% per quintile across all countries.*

### Section 9. Average pain intensity across the lifespan

Model-estimated average pain intensity among individuals reporting pain (NRS 1–10) followed a non-linear trajectory across the lifespan (Extended Data Figure 6). Intensity rose steeply from childhood, with the most rapid gains occurring around age 20 in both sexes, before peaking at approximately 5.0 in women and 4.6 in men around age 48. After midlife, intensity declined gradually, falling to approximately 3.5 in men and 4.0 in women by age 100. Women reported consistently higher intensity than men across the full age range, with a mean difference of approximately 0.4–0.5 NRS points that widened from early adulthood through midlife. These reference trajectories provide the distributional basis for individual-level percentile scoring in the Global Lifespan Pain Benchmarking Tool (Section 10).

### Section 10. Global benchmarking framework and interactive application

To facilitate the application of the reference curves for exploratory public health surveillance, we developed the Global Lifespan Pain Benchmarking Tool — an open-source interactive web application enabling external users to benchmark independent cohort datasets against the global reference standards derived in this study, without requiring access to the individual participant data (IPD) or advanced programming expertise.

The framework addresses two fundamental questions for any external population: (1) Is the observed pain burden higher or lower than demographically expected? and (2) Is this deviation statistically meaningful given the known heterogeneity of global pain reporting? Given the substantial cultural and methodological variability inherent in self-reported pain, this tool is intended for exploratory insight and hypothesis generation rather than definitive clinical inference.

#### Statistical Methodology

The benchmarking tool employs a Conditional Maximum Likelihood Estimation (MLE) strategy for out-of-sample prevalence estimation. The global fixed-effects trajectories derived in the primary analysis are treated as fixed reference values. For each participant in the external dataset, an expected probability ($P_{exp}$) is assigned based on age, sex, recall window (short [past week], medium [past month], or long [past ≥3 months]), and case definition (chronic vs. general). A study-specific log-odds deviation ($u_{new}$) is then estimated via an intercept-only logistic GLM with the logit-transformed expected probabilities entered as an offset term. The resulting intercept represents the log-odds shift required to maximise the likelihood of the external data conditional on the global reference.

To determine whether an observed shift constitutes a statistical anomaly, it must be standardised against the expected between-study variance. Global pain reporting exhibits substantial heterogeneity attributable to cultural, linguistic, and methodological factors, with heavy-tailed distributions of study-level effects. To prevent extreme outliers in the reference pool from artificially widening the tolerance band for normality, between-study heterogeneity (τ) was estimated using the Median Absolute Deviation (MAD) of the study-level random effects from the primary models, scaled by the constant factor 1.4826 to provide a consistent estimator of the standard deviation under normality. The resulting standardised Z-score quantifies the deviation of the external cohort ($u_{new}$) relative to this heterogeneity estimate (τ) and the within-study estimation uncertainty ($SE_u$):

$$Z = \frac{u_{new}}{\sqrt{\tau^2 + SE_u^2}}$$

where $SE_u$ is the standard error of the estimated offset intercept, which decreases as a function of external cohort sample size. This formulation ensures that benchmarking thresholds are defined by the central consensus of global studies, while guarding against over-interpretation of random fluctuations in small external datasets.

The tool additionally allows benchmarking of pain intensity when numeric rating scale (NRS) data are available. For cohorts reporting NRS pain scores, each participant receives a population-referenced percentile relative to the global age- and sex-specific reference distribution derived from the intensity models. Because NRS scores are bounded (1–10), right-skewed, and heteroscedastic, percentiles are estimated from a Gamma distribution parameterised using the model-derived mean intensity and the within-study coefficient of variation from the fitted reference model. The resulting percentile reflects where an individual's reported pain intensity lies relative to the expected distribution for their demographic profile.

**Application Architecture and Deployment**

The framework is deployed via a cloud-hosted web application [https://evppainlab.shinyapps.io/global-pain-benchmark/] which allows users to upload cohort data (CSV format) directly within their browser for instantaneous analysis; no uploaded data are stored or transmitted beyond the active session.

**Interactive Visualisations and Output**

The application generates a suite of automated visualisations to contextualise population-level pain burden (Extended Data Figure 7). Users may toggle between sex-stratified and combined-cohort views, and define custom demographic stratification variables.

*Global Pain Scorecard:* A lollipop chart reporting the magnitude of deviation (Observed-to-Expected [O/E] Ratio) and statistical significance (Z-Score) for each of the 11 pain sites simultaneously. Each site is assigned to a colour-coded deviation zone: Within Norm ($|Z| < 1.28$) indicates prevalence consistent with standard global variation; Above/Below Norm ($1.28 \leq |Z| < 1.96$) indicates deviation exceeding the central range of global heterogeneity; Far Above/Far Below Norm ($|Z| \geq 1.96$) flags a statistical outlier falling outside the 95% global prediction interval.

*Anatomical Pain Profile:* A polar radar chart mapping the O/E ratio across all 11 bodily sites in anatomical order (head to foot), which can be used to identify the anatomical pattern of a cohort's pain burden relative to a globally aligned expectation (O/E = 1.0).

*Lifespan Trajectory Overlay:* An age-prevalence fan chart superimposing the cohort's observed LOESS trend against the global reference trajectory and its 50%, 75%, and 95% prediction intervals. This visualisation allows users to identify at which life stages their cohort diverges most substantially from global norms, and whether that divergence is consistent or age-specific.
